# Unsupervised representation learning improves genomic discovery and risk prediction for respiratory and circulatory functions and diseases

**DOI:** 10.1101/2023.04.28.23289285

**Authors:** Taedong Yun, Justin Cosentino, Babak Behsaz, Zachary R. McCaw, Davin Hill, Robert Luben, Dongbing Lai, John Bates, Howard Yang, Tae-Hwi Schwantes-An, Yuchen Zhou, Anthony P. Khawaja, Andrew Carroll, Brian D. Hobbs, Michael H. Cho, Cory Y. McLean, Farhad Hormozdiari

**Author notes:** Joint supervision (Cory Y. McLean, Farhad Hormozdiari). Insitro, South San Francisco, CA 94080, USA.

## Abstract

High-dimensional clinical data are becoming more accessible in biobank-scale datasets. However, effectively utilizing high-dimensional clinical data for genetic discovery remains challenging. Here we introduce a general deep learning-based framework, REpresentation learning for Genetic discovery on Low-dimensional Embeddings (REGLE), for discovering associations between genetic variants and high-dimensional clinical data. REGLE uses convolutional variational autoencoders to compute a *non-linear, low-dimensional, disentangled embedding* of the data with highly heritable individual components. REGLE can incorporate expert-defined or clinical features and provides a framework to create accurate disease-specific polygenic risk scores (PRS) in datasets which have minimal expert phenotyping. We apply REGLE to both respiratory and circulatory systems: spirograms which measure lung function and photoplethysmograms (PPG) which measure blood volume changes. Genome-wide association studies on REGLE embeddings identify more genome-wide significant loci than existing methods and replicate known loci for both spirograms and PPG, demonstrating the generality of the framework. Furthermore, these embeddings are associated with overall survival. Finally, we construct a set of PRSs that improve predictive performance of asthma, chronic obstructive pulmonary disease, hypertension, and systolic blood pressure in multiple biobanks. Thus, REGLE embeddings can quantify clinically relevant features that are not currently captured in a standardized or automated way.

## 1 Introduction

High-dimensional clinical data (HDCD) provide a unique opportunity to reveal the genetic architecture of diseases and complex traits when coupled with biobank-scale genetic data [1, 2, 3, 4, 5, 6]. However, we lack statistical methods to fully utilize HDCD in genome-wide association studies (GWAS), as standard GWAS require the phenotype of interest to be encoded as a single scalar. Multiple methods have been developed to use HDCD in GWAS, but each has unique limitations.

A natural procedure to use HDCD in GWAS is to perform GWAS on every single data coordinate (e.g. time points or pixels). For example, prior work performed GWAS on each recorded point of electrocardiograms to identify its genetic architecture [7]. There are multiple shortcomings of this approach: 1) running GWAS on thousands of phenotypes can be prohibitively computationally expensive, 2) HDCD often have a correlation structure in which the actual number of degrees of freedom is much lower than the number of coordinates in the data, and 3) multiple hypothesis testing correction for highly correlated coordinates reduces statistical power [8, 9]. One popular approach to address these issues is to use principal component analysis (PCA) [10] on the HDCD and then perform GWAS on a subset of the principal components (PCs) [11]. However, PCA assumes a linear relationship between the raw HDCD and the underlying biological factors of interest, and does not explicitly model temporal or spatial structure of HDCD. This incomplete representation of the data can lead to suboptimal downstream genetic analysis.

Machine learning-based (ML-based) phenotyping uses HDCD as input to a supervised machine learning model to predict trait labels, and then performs GWAS using the model predictions as the target phenotype [3, 12, 6]. While ML-based phenotyping can augment standard GWAS on manually defined trait labels, the supervised model only learns signals related to the specific target trait. Additionally, for the common case in which the supervised model uses deep learning, many labeled examples may be required to achieve good performance.

The most common method for GWAS on HDCD uses a small number of expert-defined features (EDFs) extracted from the HDCD as the target phenotypes. For example, spirograms are a graphical representation of spirometry test results, a widely-used clinical test for lung function that measures airflow and volume over time [13, 14]. Spirograms can be summarized into EDFs including forced vital capacity (FVC), forced expiratory volume in the 1st second (FEV_1_), FEV_1_/FVC, peak expiratory flow (PEF) and forced mid-expiratory flow (FEF_25-75%_) [15]. Spirogram EDFs are used in clinical settings to diagnose diseases such as COPD [16, 17]. In another example, photoplethysmograms (PPG) measure volumetric changes in peripheral blood circulation using infrared light. Previously studied EDFs of PPG include the presence (or absence) of a notch, the position of the notch, the position of the peak, the position of the shoulder, and the peak-to-peak time [18, 19, 20, 21, 22]. PPG EDFs are known to detect diseases such as coronary heart disease [18]. Spirograms and PPG EDFs are heritable, and GWAS on EDFs have helped identify the genetic architecture of lung [23, 24, 25] and circulatory function [26, 27, 28]. However, EDFs may not capture the entirety of biological factors encoded in spirograms or PPG, thus GWAS on these EDFs may not exploit the full potential of these HDCD.

To overcome these limitations we develop a principled method, REpresentation learning for Genetic discovery on Low-dimensional Embeddings (REGLE), that is computationally efficient, requires no labels, and can incorporate information from EDFs if they are available. We apply REGLE in two case studies to understand the genetic architecture of lung function from raw spirograms and circulatory function from PPG. Compared to GWAS on spirogram EDFs (e.g. FEV_1_) and PPG EDFs (e.g., absence of notch), our GWAS on the learned encodings both recovers most known genetic loci linked to lung and circulatory function and also detects additional loci. We computed polygenic risk scores (PRSs) from GWAS on the learned encodings and evaluated their ability to discriminate disease risks. In the case of REGLE on spirograms, we observed strong enrichment for lung function, genetically causal links between the encodings and COPD and asthma, and significantly improved PRSs for asthma and COPD in multiple datasets and genetic ancestries. In the case of REGLE on PPGs, we observed that PRSs from GWAS on the learned encodings of PPGs can discriminate hypertension and systolic blood pressure in multiple datasets and produce stronger heart and blood vessel function enrichments. These results indicate that REGLE successfully extracts a meaningful low dimensional representation of lung function from spirograms and of circulatory function from PPG, which in turn improves genetic discovery for both organ systems.

## 2 Results

### 2.1 Overview of REGLE

REGLE consists of three main steps: 1) learning a non-linear, low-dimensional, disentangled representation (i.e. an encoding) of the HDCD, 2) performing GWAS on each encoding coordinate, and 3) using PRSs from the encoding coordinates as genetic scores of general biological functions, and potentially combining them to create a PRS for a disease or trait of interest (Figure 1).

**Figure 1:**
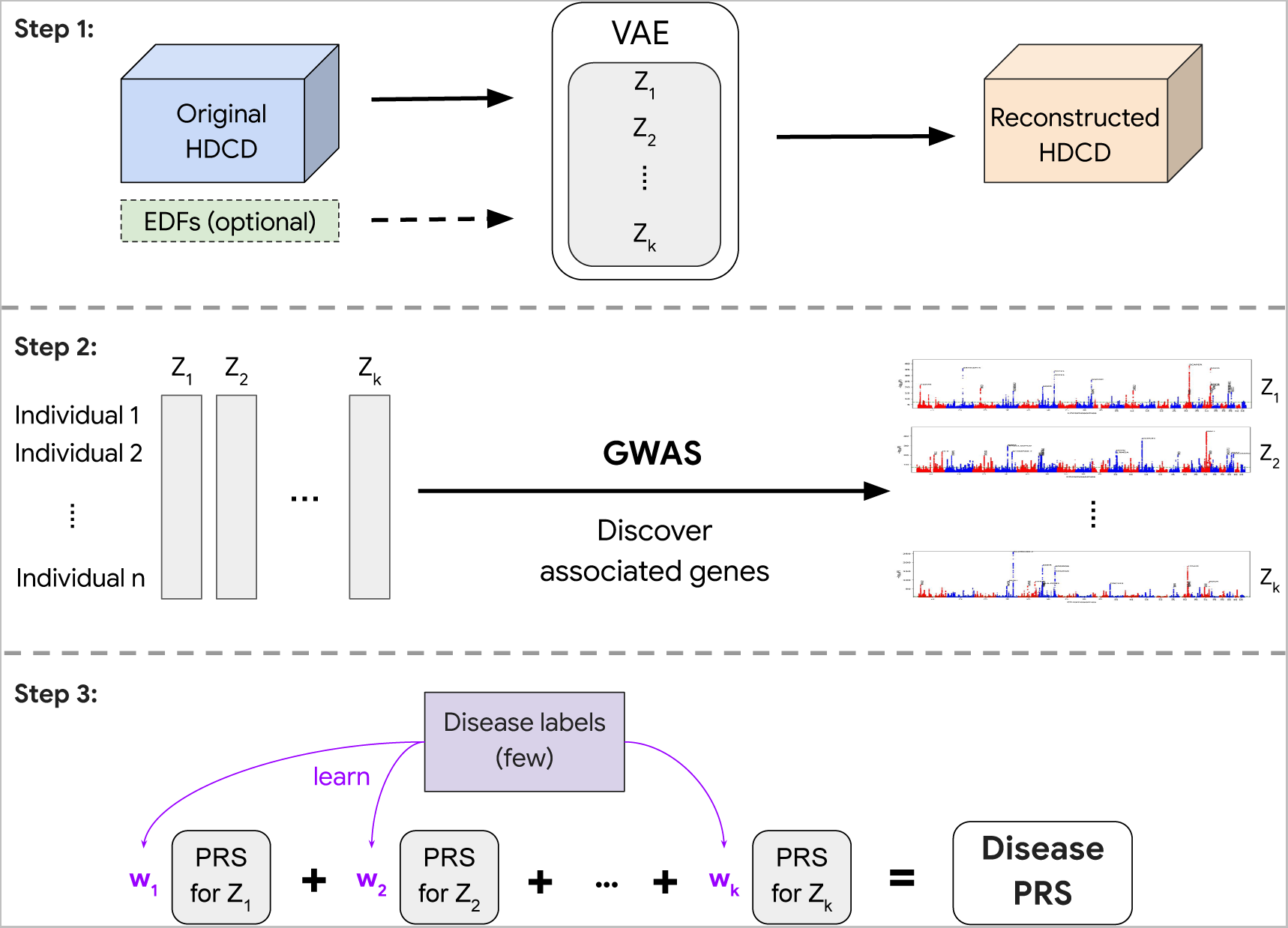
Overview of representation learning for genetic discovery on low-dimensional embeddings (REGLE). In Step 1, we learn a low-dimensional embedding using a VAE where we optionally condition the decoder on EDFs. In Step 2, we perform GWAS on all learned variables (and EDFs if they are used). Finally in Step 3, we train a small linear model to learn weights for each latent variable PRS to obtain the final disease-specific PRS.

In the first step, we train a variational autoencoder (VAE) [29] to compress and reconstruct HDCD (Figure 1 and Methods). Autoencoders consist of a pair of function approximators, typically called an encoder and a decoder, connected by a low-dimensional “bottleneck” layer. The encoder summarizes the input data efficiently into a small set of numbers represented at the bottleneck layer, and the decoder reconstructs the input data from the low-dimensional summary [30]. VAEs [29] are a special type of autoencoders that introduce stochasticity in the encoder. The VAE implicitly forces the learned encodings to be relatively disentangled [31], i.e. the encodings have relatively uncorrelated coordinates and separable biological factors can be better captured in each coordinate.

In addition, REGLE enables relevant EDFs to be optionally included in the input to the decoder of the model, so that the encoder is encouraged to learn only the residual signals not represented by the EDFs (Figure 1). This ability to incorporate prior knowledge of important data features (from users or clinicians) is one of the key advantages of REGLE.

In the second step, we perform GWAS independently on each learned encoding coordinate for all individuals (Methods). In the final step, we compute coordinate-specific PRSs that represent intermediate genetic scores of biological function, and linearly combine these coordinate-specific PRSs into a single disease-specific PRS by training on a small number of individuals with disease labels (Figure 1).

### 2.2 Overview of REGLE on spirograms

Spirograms are a graphical representation of clinical pulmonary function tests, typically represented by volume-time, flow-time, and flow-volume curves. Spirograms are used to diagnose respiratory diseases such as COPD and understand lung function [32, 24, 6]. To understand the genetic architecture of human lung function, we applied REGLE to obtain low-dimensional representations of spirogram curves, which we call spirogram encodings (SPINCs) (Figure 2). To construct SPINCs, we trained a convolutional VAE [29] to reconstruct spirograms (volume-time and flow-time; Figure 2a and Methods). In addition, we constructed another set of encodings we call *residual* spirogram encodings (RSPINCs) by injecting five EDFs (FEV_1_, FVC, FEV_1_/FVC, PEF, and FEF_25-75%_) into the input to the decoder of the VAE to reconstruct flow-volume (Figure 2a). We generated SPINCs and RSPINCs for all individuals (*n*=351,120) in UK Biobank [33] using their first visit spirogram, excluding individuals whose spirogram failed our quality control (QC) measures (Methods). We used 80% of the European-ancestry individuals (*n*=259,692) to train the (R)SPINCs models and 20% (*n*=65,266) to evaluate the reconstruction performance and choose hyperparameters (Supplementary Figure 1, Supplementary Table 1, and Methods). Using just 5 SPINCs (the same as the number of common spirogram EDFs), we observed highly accurate reconstruction of the input spirograms (Figure 2b). SPINCs consistently outperformed an equivalent number of PCs in terms of reconstruction accuracy at small latent dimensions (Figure 2c, Supplementary Table 2, Supplementary Notes). We observed similarly accurate reconstructions using EDFs+RSPINCs and confirmed that the addition of RSPINCs improves the reconstruction quality significantly, compared to using a decoder-only model to reconstruct curves using EDFs only (Supplementary Figure 2); we used 2 RSPINCs to balance the number of additional coordinates and the reconstruction accuracy. Importantly, the learned representations are highly consistent when trained with multiple different initializations (Supplementary Figure 3, Supplementary Notes).

**Figure 2:**
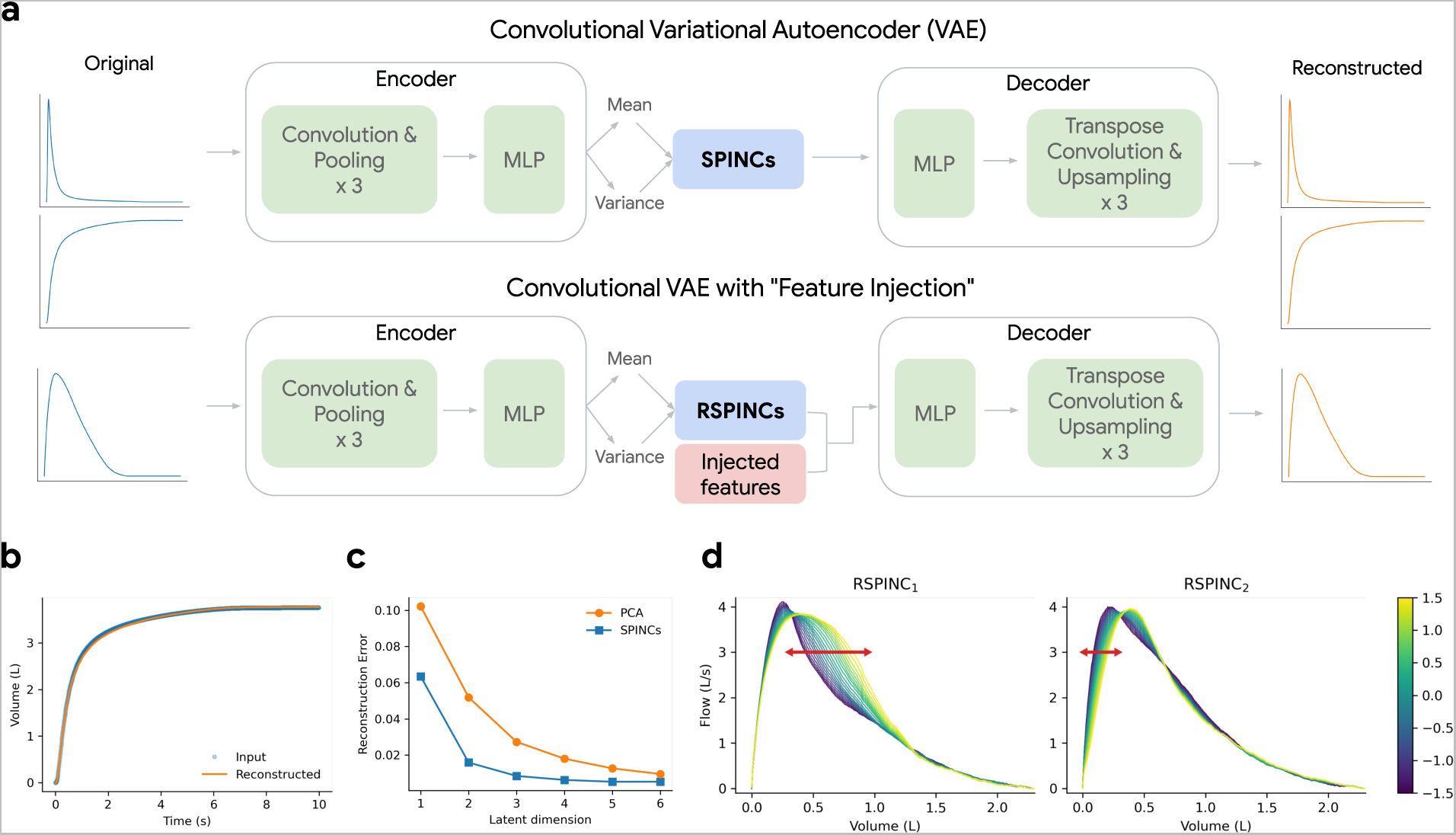
Overview of REGLE on spirograms. a) Learning spirogram encodings (SPINCs) using a convolutional variational autoencoder (VAE) and *residual* spirogram encodings (RSPINCs) using a convolutional VAE with “feature injection”, using expert-defined features (EDFs) for example. b) Reconstructing a spirogram (volume-time curve) from SPINCs (dim=5). c) Reconstruction errors (mean squared error across time points) for reconstructed spirograms using the SPINCs model and PCA with a varying latent dimension. Both the SPINCs model and PCA are trained (or “fitted”) on a training set and the reconstruction error was evaluated in a separate validation set. d) Spirograms created by RSPINCs (dim=2) decoder using a fixed set of injected features (i.e. EDFs) and varying one RSPINC coordinate while fixing the other one to be zero. Line color indicates the varying RSPINC coordinate value.

### 2.3 Overview of REGLE on PPGs

Photoplethysmograms (PPG) measure volumetric changes in blood in peripheral circulation, typically using infrared light. They are represented as pulse waveforms and used as an inexpensive and noninvasive way to measure arterial stiffness. PPGs can also be used to classify elevated blood pressure or cardiovascular disease (CVD), and to interrogate circulatory function [34, 35, 36]. To understand the genetic architecture of human circulatory function from these highly available data, we applied REGLE to obtain low-dimensional representations of PPG curves computed from a median single heartbeat (median is taken over multiple heartbeats as computed by the pulse waveform device), which we call photoplethysmogram encodings (PLENCs) (Figure 3). To construct PLENCs, we trained a convolutional VAE [29] to reconstruct PPGs (Figure 3a and Methods). We generated PLENCs for all individuals (*n*=170,714) in UK Biobank [33] using their first-visit PPG, excluding individuals whose PPG failed our QC measures (Methods). We used 80% of the European-ancestry individuals (*n*=136,239) to train the PLENCs models and 20% (*n*=34,475) to evaluate the reconstruction performance and choose hyperparameters (Supplementary Figure 4, Supplementary Table 1, and Methods). With just 5 PLENCs (the same as the number of PPG EDFs), we observed highly accurate reconstruction of the input PPG (Figure 3b and Supplementary Table 3). PLENCs consistently outperformed an equivalent number of PCs in terms of reconstruction accuracy at small latent dimensions (Figure 3c, Supplementary Notes).

**Figure 3:**
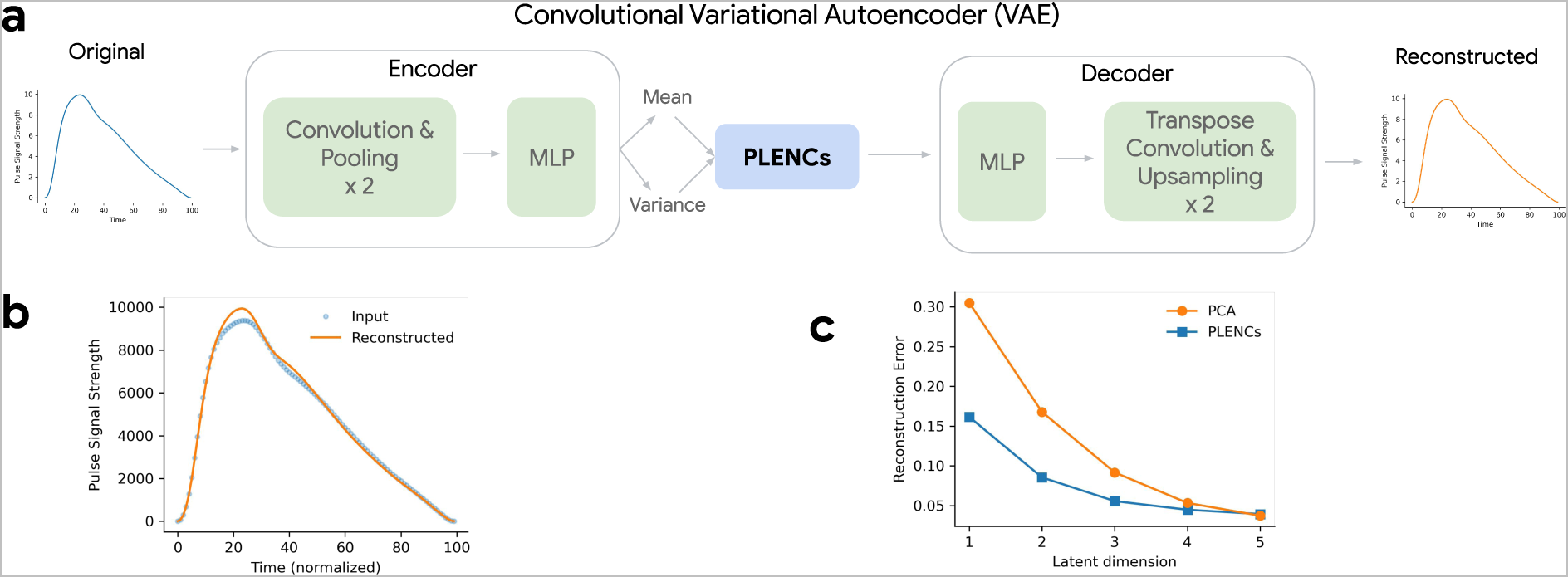
Overview of REGLE on PPG. a) Learning PPG encodings (PLENCs) using a convolutional variational autoencoder (VAE). b) Reconstructing PPG from PLENCs (dim=5). c) Reconstruction errors (mean squared error across time points) for reconstructed PPGs using the PLENCs model and PCA with a varying latent dimension. Both the PLENCs model and PCA are trained (or “fitted”) on a training set and the reconstruction error was evaluated in a separate validation set.

### 2.4 (R)SPINCs are partially interpretable

To interpret the influence of RSPINC coordinates on spirogram shape, we fixed the values of EDFs (obtained from a randomly selected individual in the validation set) and varied one RSPINC coordinate while keeping the other one fixed at zero, and generated the corresponding flow-volume spirograms using only the decoder portion of the RSPINCs model (Figure 2d). A typical flow-volume spirogram consists of two distinct parts: the first part, a relatively brief part to reach peak flow where the flow increases monotonically as the volume increases, and the second part, the main part of the spirogram where the flow decreases monotonically. In Figure 2d, we clearly observed that varying the first coordinate of RSPINCs amounts to widening or narrowing of the second part (negative slope) while keeping the first part relatively fixed. Similarly, varying the second coordinate of RSPINCs widens or narrows the first part (positive slope) while keeping the second part relatively fixed. Notably, when varying either coordinate of RSPINCs, the maximum flow value (PEF) and the final volume value (FVC) stay roughly the same, as expected since all EDFs were fixed.

### 2.5 (R)SPINCs and PLENCs encode information beyond EDFs

Some coordinates of SPINCs are highly correlated with known EDFs (Supplementary Figure 5). For example, the 3rd coordinate of SPINCs is 96% correlated with FVC and 94% correlated with FEV_1_, while the 2nd coordinate is 73% correlated with FEV_1_/FVC (after flipping the signs) (Supplementary Figure 5). Both RSPINCs coordinates have low correlation (|*R*| *<* 0.3) with all EDFs, which is expected since they were encouraged to learn only residual signals not captured by the EDFs (Supplementary Figure 5). (R)SPINCs are also correlated with other predictors of lung function (“covariates”), such as age, sex, height, body mass index, and smoking status (Supplementary Figure 5). To investigate if (R)SPINCs include information beyond EDFs and covariates, we residualized both the EDFs and the covariates from (R)SPINCs and computed correlation with tabular UK Biobank features. Multiple groups of fields strongly and significantly correlated with the (R)SPINCS even after residualizing the EDFs and the covariates, including asthma, breathing issues, cognitive function, and allergies (Supplementary Tables 4 to 6).

We observed similar qualitative results for PLENCs, which also show strong correlation with known PPG EDFs (Supplementary Figure 6). For example, the 3rd coordinate of PLENCs is 75% correlated with the position of the shoulder and 74% correlated with the position of the peak, while the first coordinate is 58% correlated with the pulse rate (after sign flip). PLENCs also show correlation with other predictors of circulatory function (Supplementary Figure 6). After residualizing EDFs and covariates from PLENCs, we still observed multiple groups of fields strongly and significantly correlated with the residualized PLENCs coordinates, including pulse rate, systolic and diastolic blood pressure, and position of shoulder (Supplementary Tables 7 and 8).

Finally, using the Cox proportional hazards model, we observed that both (R)SPINCs and PLENCs are associated with overall survival. For example, the third coordinate of SPINCs had a hazard ratio of 0.68 (95% CI, 0.65 to 0.71; P=1.6E-83) implying the hazard of death decreased by 32% per one standard deviation increase in the coordinate (Supplementary Notes, Supplementary Figures 7 to 9, Supplementary Table 9, Methods).

### 2.6 REGLE detects novel loci for lung and circulatory functions

We generated SPINCs (dim=5), RSPINCs (dim=2, in addition to 5 EDFs) for all individuals with valid first-visit spirograms in UK Biobank (Supplementary Figures 1, 10 and 11; Methods) and PLENCs (dim=5) for all individuals with valid first-visit PPGs in UK Biobank (Supplementary Figure 4; Methods). We then performed GWAS on all European-ancestry individuals across all encoding coordinates and 5 EDFs using BOLT-LMM [37, 38], adjusting for covariates (Supplementary Notes; Methods; Supplementary Figures 12 to 16, Supplementary Figures 17 and 18, and Supplementary Figures 19 to 23). S-LDSC analysis demonstrates that each of the SPINCs, RSPINCs, and PLENCs have significant SNP heritability. For example, the SNP heritability of SPINCs ranged from 4.3% for SPINC_5_ to 24.8% for SPINC_3_. Importantly, the RSPINCs, which are relatively uncorrelated with the EDFs, were also significantly heritable (4.5-16.2%), indicating the existence of additional heritable signal within the spirograms (Supplementary Table 10).

GWAS on 5 SPINCs detected 575 independent genome-wide significant (GWS) loci (*R*^2^ *≤* 0.1 and *P ≤* 5 *×* 10*^−^*^8^) after merging hits within 250kb together (Table 1, Methods). To compare our results to known lung function loci from previous literature, we combined the largest published GWAS on lung function (using FEV_1_, FVC, PEF, and FEV_1_/FVC) from Shrine et al. [25] (580,869 individuals, compared to our 324,702 individuals in UK Biobank) with all lung function-related loci in the NHGRI-EBI GWAS Catalog [39] (Methods). This resulted in 1104 independent loci after merging loci by distance (250kb), hereafter referred to as “previously known loci”. Most GWS loci from SPINCs and EDFs+RSPINCs recover previously known loci (89% for SPINCs, 90% for EDFs+RSPINCs). Out of 575 GWS SPINCs loci, 65 (11%) were not previously known, compared to 32 from EDFs and 15 from PCA. Of 659 EDFs+RSPINCs GWS loci, 63 (10%) were not previously known (Table 1). Functional enrichment analysis with GARFIELD [40] shows that these loci are enriched for lung tissue DNase I hypersensitive sites (Supplementary Figures 24 to 28; Supplementary Figures 29 and 30; Supplementary Notes) and the EDFs+RSPINCs loci show stronger ontology term enrichments than EDFs loci alone (Supplementary Figure 31) using GREAT [41]. Notably, we found a strong enrichment for RSPINC_2_ in blood (Supplementary Figure 30). Finally, we performed multiple analyses to ensure these novel loci were not detected by EDFs or previous works (Supplementary Notes; Supplementary Tables 11 and 12).

GWAS on 5 PLENCs detected 90 independent GWS loci (Table 1, Methods). To compare our results to known cardiovascular function loci from previous literature, we utilize all cardiovascular function-related loci in the NHGRI-EBI GWAS Catalog [39] (Methods; 520 independent known loci after merging by distance). For cardiovascular function and PPG, EDFs were the absence of notch, the position of notch, the position of peak, the position of shoulder, and the peak-to-peak time. Out of 90 GWS PLENCs loci, 50 (56%) were not previously known (Table 1 and Supplementary Table 13). Functional enrichment analysis with GARFIELD [40] shows that PLENCs GWS loci are enriched for fetal heart, heart, and blood vessel tissue DNase I hypersensitive sites (Supplementary Figures 32 to 36; Supplementary Notes).

**Table 1:**
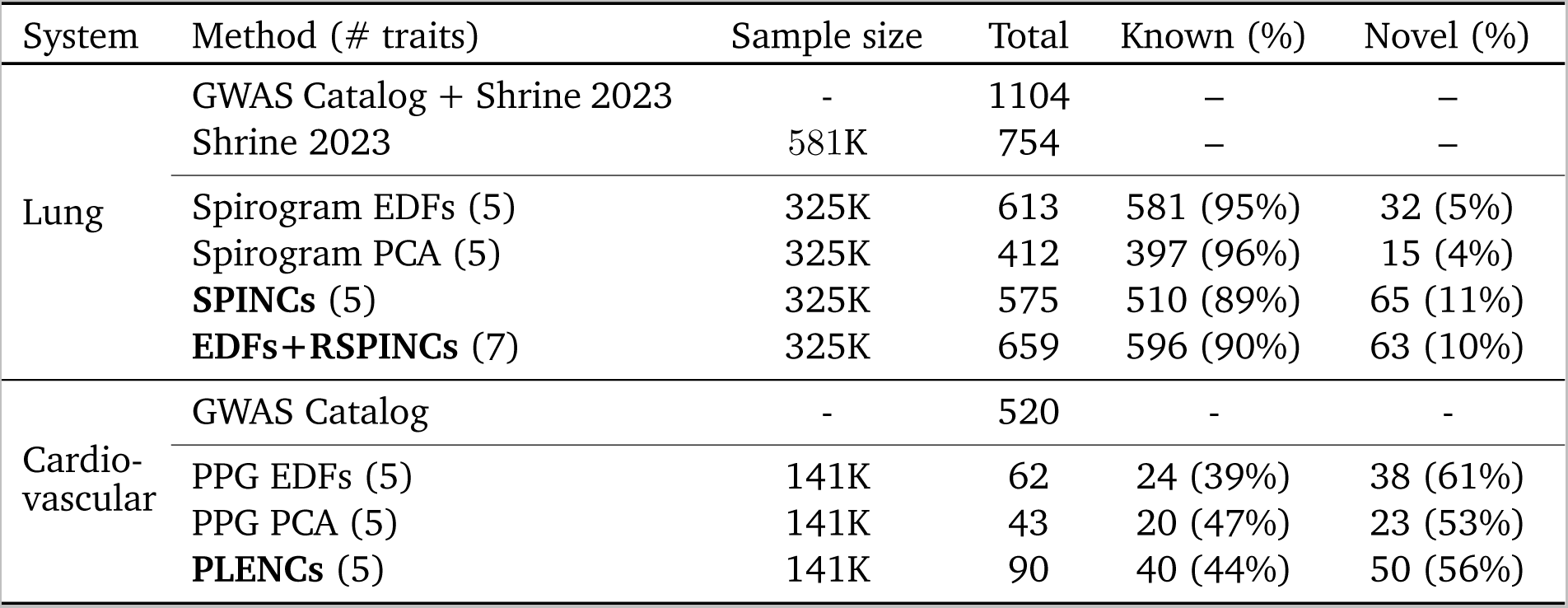
Comparison of GWAS significant loci. For lung function and spirograms, expert-defined features (EDFs) are FEV_1_, FVC, FEV_1_/FVC, PEF, and FEF_25-75%_, and “known” and “novel” is in reference to lung function loci in GWAS Catalog and Shrine et al. [25]. For cardiovascular function and PPG, EDFs are the absence of notch, the position of notch, the position of peak, the position of shoulder, and the peak-to-peak time, and “known” and “novel” is in reference to cardiovascular disease loci in GWAS Catalog.

### 2.7 (R)SPINCs improves asthma and COPD PRS over EDFs in UK Biobank

We computed PRSs using BOLT-LMM [37, 38] effect sizes for 5 SPINCs and 2 RSPINCs coordinates, in addition to 5 spirogram EDFs. We treat these sets of PRSs as intermediate genetic scores for lung function. Given a specific trait, a set of such intermediate PRSs, and a (small) set of individuals for whom the trait status is available, one can combine the intermediate PRSs into a single trait-specific PRS via a weighted linear sum of the intermediate PRSs. The weights of the linear score can be learned in any dataset composed of individuals with available genetics and trait labels (Figure 1). We created disease-specific PRSs for asthma and COPD from three sets of intermediate PRSs: 1) 5 EDFs, 2) 5 SPINCs, and 3) 5 EDFs plus 2 RSPINCs. We learned the disease-specific PRS weights within the modeling set (*n*=324,958) of European ancestry individuals in UK Biobank using medical-record-based asthma and COPD statuses. To evaluate the performance of each disease-specific PRS, we computed the accuracy of the PRS in a completely separate set of individuals of European ancestry (*n*=110,722) not previously used for model training or GWAS.

We observed that the top-decile high-risk individuals based on the SPINCs asthma PRS have significantly higher asthma prevalence than individuals in the top-decile of the EDF PRS (Figure 4, Supplementary Table 14). The SPINCs asthma PRS more effectively stratifies the risk groups than EDFs on both ends of the risk spectrum. In addition, we observed statistically significant improvements in AUC-ROC, AUC-PR, and Pearson correlation by using the SPINCs asthma PRS (Supplementary Table 14). We observed the same trend for COPD (Figure 4, Supplementary Table 15). Furthermore, we observed that the EDFs+RSPINCs PRS outperforms the EDFs PRS on all metrics for both asthma and COPD; The differences in AUC-ROC, AUC-PR, and Pearson correlation are statistically significant for both diseases, and the difference in the top decile prevalence is significant for asthma (Figure 4, Supplementary Tables 14 and 15). We observed that the SPINCs COPD PRS outperforms the FEV_1_/FVC PRS (Supplementary Table 15) for predicting medical-record-based COPD, despite FEV_1_/FVC having previously been shown to be one of the best phenotypes for generating a COPD PRS, even outperforming the binary COPD PRS [6]. These results provide further evidence that SPINCs capture more genetic determinants of lung function related to asthma and COPD than the same number of EDFs, and that RSPINCs capture additional genetic factors not captured by the EDFs.

**Figure 4:**
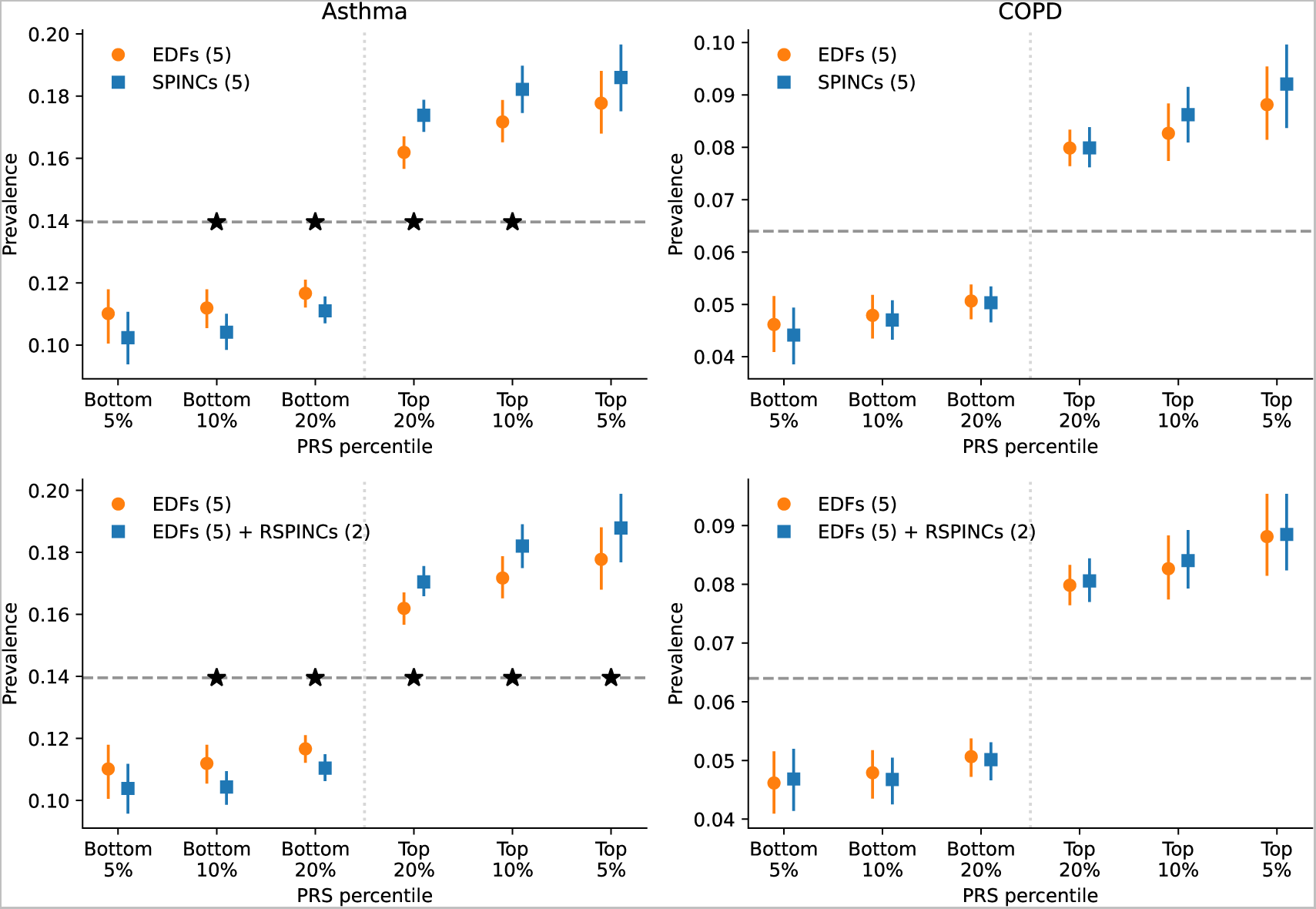
PRS using SPINCs and RSPINCs in UK Biobank. Combined PRS for medical-record-based asthma and COPD using three sets of intermediate PRS: 5 EDFs, 5 SPINCs, and 5 EDFs + 2 RSPINCs. Each set of PRS is combined by a linear model trained using the target phenotype labels and the prevalence of the phenotypes in the top and bottom 5%, 10%, and 20% PRS individuals is evaluated in a separate evaluation set. Vertical line segments indicate 95% confidence interval generated by bootstrapping (300 repetitions). The horizontal dashed line shows the total prevalence. Star (*) sign indicates a statistically significant difference between the two methods using *paired* bootstrapping (300 repetitions) with 95% confidence. Lower is better for the bottom percentiles; higher is better for the top percentiles.

**Figure 5:**
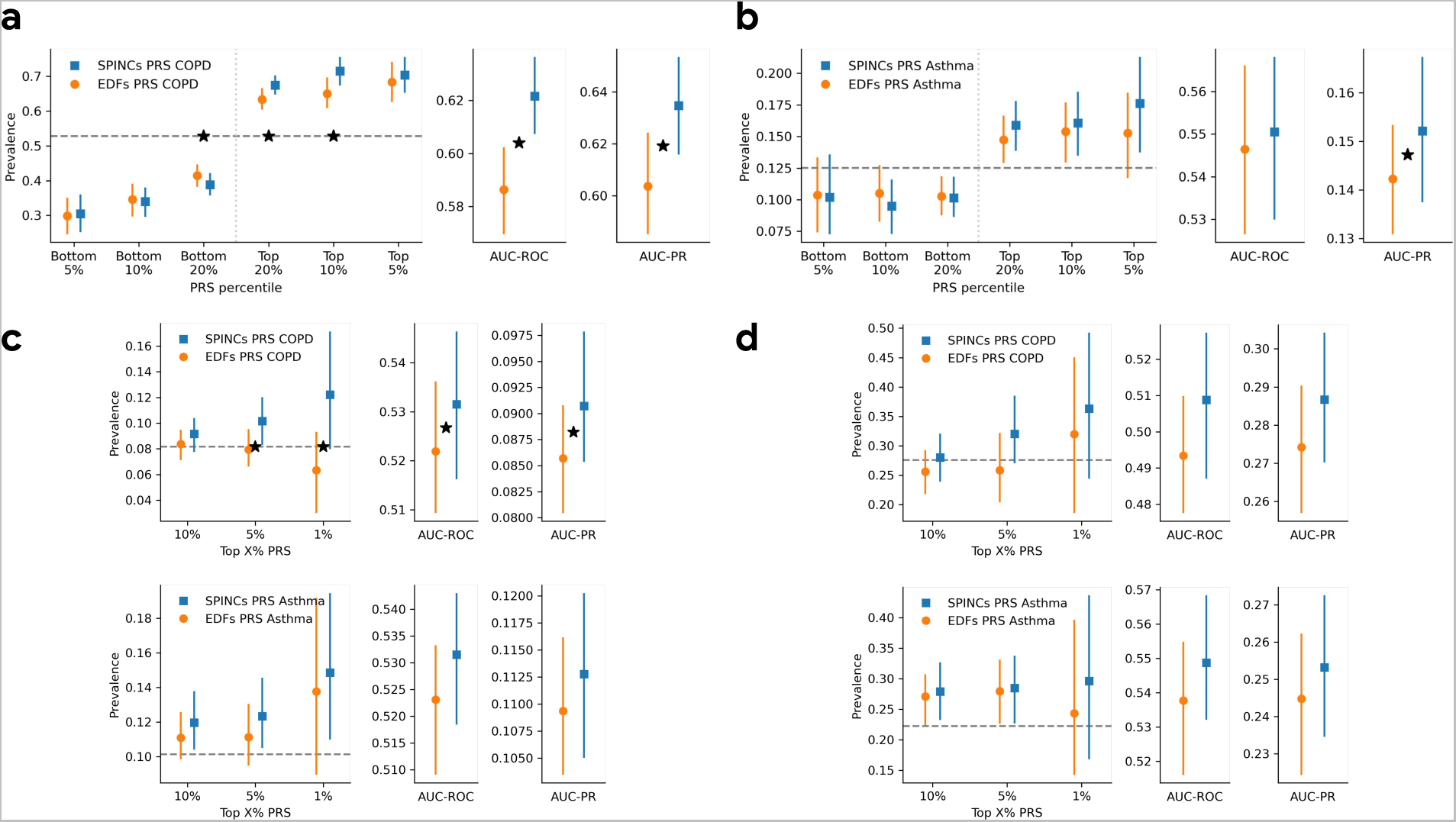
SPINCs PRS transferred to multiple independent datasets. SPINCs PRSs for COPD and asthma generated on UK Biobank are transferred to four independent datasets: COPDGene, eMERGE III, EPIC Norfolk, and Indiana Biobank. a) PRS evaluation in COPDGene dataset on COPD. b) PRS evaluation in eMERGE III dataset on asthma. c) PRS evaluation in EPIC Norfolk study on COPD and asthma. d) PRS evaluation in Indiana Biobank on COPD and asthma. In all figures, solid vertical intervals represent 95% confidence interval generated by bootstrapping (300 repetitions). The horizontal dashed lines show the total prevalence in the evaluation set. Star symbols indicate a statistically significant difference between the two methods using paired bootstrapping (300 repetitions) with 95% confidence.

Finally, since each disease-specific PRS is constructed from intermediate (R)SPINCs coordinate PRSs using just 5–7 learned weights, we explored whether these weights could be learned from a subset of the training data. For both asthma and COPD, (R)SPINCs-based PRS fit with as few as 100 disease cases performed indistinguishably from those trained on the full training data of 45,609 and 12,468 cases, respectively (Figure 1 “Step 3”, Supplementary Figure 37). We also evaluated PRSs generated by GWAS with a cohort-level phenotype adjustment using inverse-normal transformation. We observed fewer significant differences with this adjustment, though SPINCs and EDFs+RSPINCs still maintain some statistically significant differences for asthma PRS (Supplementary Figure 38).

### 2.8 (R)SPINCs PRS transferred to independent datasets and ancestry groups

The performance of PRS computed in one dataset can degrade significantly when transferring the variant weights directly to another dataset or a different ancestry group [42]. In addition, the quality of COPD and asthma labels in different datasets can vary widely (e.g. using physician diagnoses, self report, or medical records). To test the generalizability of our (R)SPINCs PRSs to individuals outside of UK Biobank and non-European ancestry individuals, we transferred the variant weights from the asthma and COPD PRSs in UK Biobank directly to the Genetic Epidemiology of COPD (COPDGene) dataset [43], the eMERGE III dataset, the EPIC-Norfolk dataset [44], and Indiana Biobank dataset [45] (Supplementary Table 16).

For the COPDGene dataset, we computed PRS of all individuals using the same variant effect sizes obtained by BOLT-LMM in UK Biobank and the same linear weights to combine the EDFs, EDFs+RSPINCs, and SPINCs PRS as before after matching variants. We used the “race” field in COPDGene as a proxy for genetic ancestry and computed PRS performance in the two available subsets separately: “Non-Hispanic White” and “African American”. We observed that for COPD, the SPINCs COPD PRS outperforms the EDFs COPD PRS for both subset of individuals in COPDGene for all four evaluation metrics (AUC-ROC, AUC-PR, top-decile prevalence, and Pearson correlation). In the “Non-Hispanic White” subset (*n*=6,576), which matches the UK Biobank ancestry group on which the PRS was trained, all four metrics are statistically significant (paired bootstrapping; Figure 5a, Supplementary Table 17). In the “African American” subset (*n*=3,140), differences were statistically significant for AUC-ROC and Pearson correlation (Supplementary Table 17). The EDFs+RSPINCs COPD PRS significantly outperformed the EDFs COPD PRS in “Non-Hispanic White” in AUC-ROC and Pearson correlation, but did not in the “African American” subset (Supplementary Table 17).

We also transferred the UK Biobank PRSs to eMERGE III (“White” subset, *n*=8,288), EPIC Norfolk (self-reported “White”, *n*=21,010), and the Indiana Biobank (mostly European, see Methods, *n*=5,254), to evaluate asthma, asthma and COPD, and asthma and COPD, respectively. We observed consistent improvement from using SPINCs PRSs over EDFs PRSs for both COPD and asthma phenotypes for top-percentile prevalences, AUC-ROC, and AUC-PR. The improvement was statistically significant for AUC-PR and the top 1% and 5% prevalence in eMERGE III, and for AUC-ROC and AUC-PR in EPIC Norfolk (Figure 5b-d).

### 2.9 PLENCs improves hypertension and systolic blood pressure PRS over EDFs

We computed PRSs using BOLT-LMM for 5 PLENC coordinates in addition to 5 PPG EDFs, and utilized these sets of PRSs as intermediate genetic scores for cardiovascular function. Since PPG is known to carry signals about blood pressure [46], we created trait-specific PRSs for hypertension (HTN; a binary trait) and systolic blood pressure (SBP; a continuous trait) using the REGLE framework (Figure 1, Supplementary Table 18). We evaluated HTN and SBP PRS generated by PLENCs and PPG EDFs in independent datasets (COPDGene, eMERGE III, and EPIC-Norfolk) in addition to the held-out test set in UK Biobank. For HTN we measured the top and bottom PRS group prevalences, AUC-ROC, and AUC-PR, and for SBP we measured the Pearson correlation and the Spearman rank correlation. Note that we did not perform HTN and SBP PRS evaluation in Indiana Biobank due to unusually high prevalence of hypertension (more than 80%) and the usage of blood pressure medication by a majority of its population.

We observed consistent trend of improvement from using PLENCs PRSs over EDFs PRSs for both HTN and SBP, with the exception of AUC-ROC of HTN in EPIC Norfolk (Figure 6). Notably, PLENCs PRS for SBP outperformed EDFs PRS for all datasets for both correlation metrics, e.g. 2*×* higher Pearson correlation (6% vs. 3%) in the UK Biobank test set, and the differences were statistically significant in three out of four datasets.

**Figure 6:**
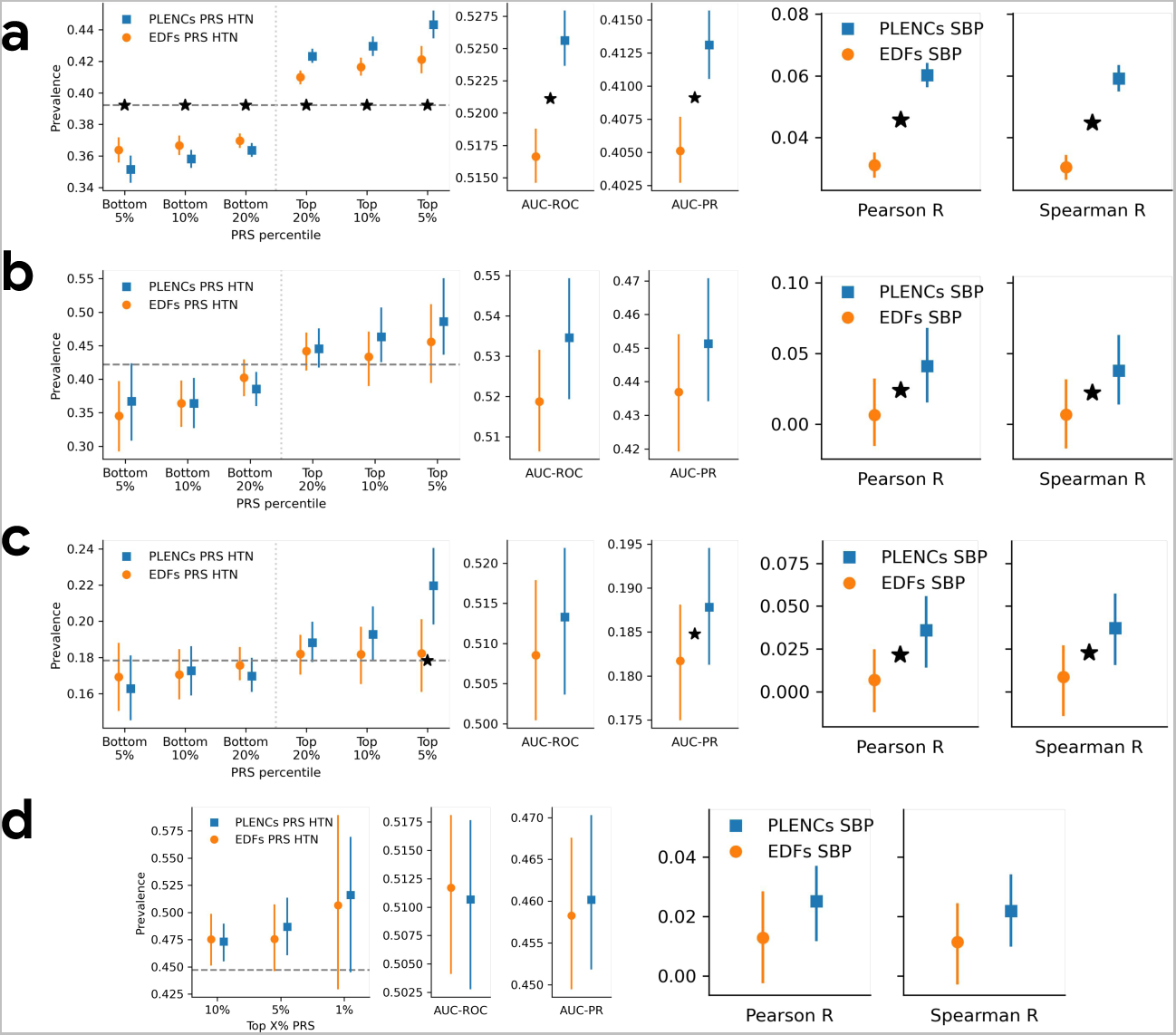
PLENCs PRS generated in UK Biobank evaluated in multiple independent datasets. PLENCs PRS for hypertension (HTN) and systolic blood pressure (SBP) generated on UK Biobank are evaluated in four independent datasets: UK Biobank, COPDGene, eMERGE III, and EPIC Norfolk. a) PRS evaluation in UK Biobank, evaluated in a separate test set not used for GWAS. b) PRS evaluation in COPDGene dataset. c) PRS evaluation in eMERGE III. d) PRS evaluation in EPIC Norfolk. In all figures, solid vertical intervals represent 95% confidence interval generated by bootstrapping (300 repetitions). The horizontal dashed lines show the total prevalence in the evaluation set. Star symbols indicate a statistically significant difference between the two methods using paired bootstrapping (300 repetitions) with 95% confidence.

### 2.10 High association between REGLE encodings and UKB phenotypes PRSs

To assess the influence of (R)SPINCs and PLENCs on traits and health outcomes, we performed phenome-wide association studies (PheWAS). We compared pruning+thresholding PRSs of all (R)SPINCs and and PLENCs coordinates to PRSs of 7,145 phenotypes computed by the Pan-UK Biobank consortium (Methods, URLs). For SPINCs, significant associations with strong correlation magnitude are driven by the 2nd and 3rd coordinates, which as mentioned above are strongly correlated with EDFs, and include expected phenotypes such as FEV_1_, FEV_1_/FVC, and PEF (Supplementary Tables 19 and 20). These coordinates also show strong correlation to other diseases and traits associated with alterations in lung function, identifying relationships with systemic lupus erythematosus [47, 48], thyroid dysfunction [49], and gluten-free diet [50]. For RSPINCs, significant correlations with strong magnitude are nearly all driven by RSPINC_1_ and also include the same diseases (Supplementary Tables 21 and 22). For PLENCs, we observed all coordinates show significant correlation with different traits including blood traits (red blood cell, eosinophil count, age high blood pressure diagnosed, and haemoglobin concentration), PPG traits (pulse rate and pulse wave reflection index), ECG traits (QRS duration and P duration), blood pressure, and vascular/heart problems (Supplementary Tables 23 and 24). The strongest correlation was obtained from 1st PLENCs coordinate with pulse rate (R=-0.67; P *≤* 1.00E-300) and ECG heart rate (R=-0.52; P *≤* 1.00E-300).

Finally, we performed latent causal variable (LCV) analysis [51] to identify potentially causal relationships between (R)SPINCs and EDFs with COPD, asthma, and other lung-related phenotypes (sarcoidosis, systemic lupus erythematosus, thyroid dysfunction, and gluten-free diet) where we observed significant correlation between their PRSs in PheWAS (Supplementary Tables 25 and 26). We observed a significantly positive genetic causality proportion (GCP) from the 2nd SPINCs coordinate to COPD, suggesting a genetic causal link from SPINCs to COPD. We also observed a significant negative GCP from the 5th SPINCs coordinate to asthma, suggesting a genetic causal link from asthma to SPINCs (Supplementary Tables 25 and 26; Supplementary Notes). Interestingly, all significant GCPs were negative from SPINCs to other lung-related phenotypes we considered, implying that the genetic causal directions are from those phenotypes to SPINCs. In the case of PLENCs, we observed that 1st PLENC coordinate has a significant negative GCP with hypertension (Supplementary Tables 25 and 26), implying a genetic causal link from hypertension to PLENCs.

## 3 Discussion

Large biobanks provide unique opportunities to identify the genetic factors underlying complex traits and diseases, but accurately phenotyping [52] the cohorts remains a core challenge. We proposed a general unsupervised deep representation learning method, REGLE (representation learning for genetic discovery on low-dimensional embeddings), to improve discovery of genetic components of high-dimensional clinical data (HDCD). We showcased the effectiveness of REGLE in spirograms and photoplethysmograms, where it produced latent variables (“encodings”) that are both partially interpretable and effective for identifying genetic variants associated with lung and circulatory functions. Multiple lines of evidence show the relevance of the model representations for quantifying general lung and circulatory functions.

Unsupervised representation learning of HDCD for genomic discovery is attractive owing to the difficulty of acquiring (or defining) manual phenotypes at scale. Prior work has explored applying transfer learning [53] and contrastive learning [54] to retinal fundus images, or multimodal autoencoders to cardiac data modalities [55]. A key strength of our method is the use of a VAE to generate the non-linear low-dimensional representations of HDCD. First, by construction, the coordinates of the latent representation are minimally correlated, which strengthens the combined power of the downstream GWASs. As a result, the PRSs of the learned encodings are also minimally correlated and contain relatively orthogonal genetic signals, which may contribute to the superior accuracy of the disease/trait specific PRS created by the REGLE framework. Second, the learned representations are stable up to changes in sign or order as we observed empirically (Section 2.4), potentially due to a grounding effect of a VAE prior in the probabilistic model. As changes in sign or order do not affect genetic discovery–though they may affect the sign of the effect size–the results of the REGLE framework are stable and replicable. Regular autoencoders without a prior do not have this stability property as they can learn any invertible linear transformation of a specific learned representation.

The architecture modification we introduce in REGLE to support expert-defined features (EDFs) enables a principled use of expert human knowledge and encourages the remaining latent coordinates to encode biological function explicitly not captured by the EDFs. This provides the opportunity to build upon and improve existing clinical practices with the extra information provided from the residual features. For example, clinical review of pulmonary function tests includes visual inspection of the curves for variation in shape. Coving of the flow volume loop is an indicator of obstruction. The ability of (R)SPINCs to identify these differences while holding EDFs constant suggest that (R)SPINCs are quantifying clinically relevant parameters that are not currently captured in a standardized or automated way. While we demonstrated the value of the method on spirograms to model lung function, it can be generalized to other HDCD modalities such as images. The improved performance of SPINCs, EDFs+RSPINCs, and PLENCs PRS over EDFs PRS provides evidence for the existence of such residual genetic information. Importantly, the (R)SPINCs PRS for respiratory diseases (asthma and COPD) and PLENCs PRS for cardiovascular functions (hypertension and SBP) combine each latent coordinate GWAS into a PRS based solely on the effect size estimates from the GWAS on the learned coordinates, and the disease-specific PRS is simply a learned weighted sum of the handful (i.e., five or seven) coordinate PRSs. This has important implications for disease risk prediction: it demonstrates that given a dataset with widespread unlabeled quantification of lung system function (i.e. spirograms) and/or circulatory system function (i.e. photoplethysmograms), genetic predictors for specific lung or cardiovascular diseases can be accurately learned with very few disease labels (enough to learn 5-7 features). We hypothesize that unsupervised quantification of other organ systems may be similarly beneficial for improving polygenic prediction across a wealth of diseases. Finally, we note that the PRS performance reported here represents a lower bound achievable by the method; jointly estimating disease-specific variant effect size estimates on the set of variants identified by the (R)SPINCs and PLENCs GWAS may further improve prediction performance.

There are several limitations to this work. First, while unsupervised representation learning of HDCD for biological function is likely beneficial across data modalities, the generalizability of the specific VAE method introduced here to even higher dimensional modalities like imaging and video has not been assessed. In particular, VAEs tend to produce blurry image reconstructions [56, 57]; whether and how that affects the ability of a VAE to encode representations meaningful for genomic discovery is important future work. Second, we did not directly optimize multiple GWASs for novel genomic discovery, but used a straightforward (conservative) method to define and merge independent associated loci. A possible extension would be to combine the signals from multiple (R)SPINC and PLENC coordinate GWAS [11] or apply methods that maximize heritability (e.g. [58]). Third, we did not fully explore model architecture and training strategies specifically for genomic discovery. Some ideas which may warrant further investigation include: 1) using previously proposed modifications to the VAE loss function and the training procedure to maximize the degree of disentanglement of coordinates while balancing the reconstruction error [59, 60, 61], 2) incorporating an additional loss term to explicitly discourage correlation between RSPINCs and EDFs, and 3) introducing (semi-) supervision in model training to overcome the limitations of purely unsupervised training [62]. Fourth, in the case spirograms, none of the spirograms in UK Biobank were obtained after bronchodilation, and the asthma and COPD labels defined using clinical records are known to be noisy [6]. Fifth, we generated individual-level spirogram representations from the first measurement, despite some individuals having up to three acceptable blows in UK Biobank. Integrating all acceptable blows from an individual could produce a more comprehensive representation of their lung function [63]. Sixth, model training was performed exclusively in individuals of European ancestry. While PRS evaluation was performed in multiple datasets and ancestries, the impact of ancestry-specific model training was not explored.

Despite these limitations, REGLE provides a mechanism for identifying genetic influences on organ function in the absence of labeled data, and naturally admits incorporating expert features in the model. It also provides a method to create disease/trait specific PRS with very few labels (i.e. in the order of hundreds). As biobanks with rich imaging, activity monitoring, medical records, and paired genetic data continue to grow, we anticipate that this or similar methods will be increasingly used to further elucidate the genetic underpinnings of human traits and diseases.

## 4 Methods

### 4.1 UK Biobank data preparation for spirograms

Spirograms from UK Biobank were sourced from the data field 3066, which contains the volume in milliliters of exhalation at 10 millisecond intervals (volume-time curve) and were preprocessed closely following the procedures in Cosentino et al. [6]. To generate flow-time curves, we approximated the first derivative of volume with respect to time by taking a finite difference in the volume-time curves. We normalized the volume-time and flow-time curves to 1000 time points, by either truncating longer curves or by right-padding shorter curves with zero (for flow-time curves) or the final value (for volume-time curves), and removed FEV_1_, FVC, PEF values in the extreme tail (top/bottom 0.5%) of the observed values and all blows that fail the acceptability provided by UK Biobank data field 3061. We used the first acceptable blow of an individual when there is more than one. In addition, we dropped all flow curves whose values don’t fall in [-10, 20], all volume curves whose values are not in [-5, 10], and all flow curves in which the proportion of nonzero values are less than 20%. Finally, we generated flow-volume curves from volume-time and flow-time curves by interpolating 1000 evenly spaced volume values between 0 and 6.58 liters (the maximum observed volume in the dataset).

We then subdivide all European ancestry individuals processed this way into a 80% training set and a 20% validation set. After additionally removing related individuals, there are 259,692 individuals in the training set and 65,266 individuals in the validation set (Supplementary Figure 1).

Asthma and COPD status were determined by medical records using self report, ICD9, and ICD10 codes as defined in Cosentino et al. [6].

### 4.2 UK Biobank data preparation for photoplethysmograms

Photoplethysmograms (PPG) from UK Biobank were sourced from the data field 4205, which contains the arterial stiffness pressure curve. Each waveform is actually a single pulse with 100 points. Then we computed the minimum, maximum, mean, and median distribution value of PPG. We keep the PPG when all four statistics fall in 0.1 and 99.9 percentiles of the related statistics values of all PPGs. We then subdivide all European ancestry individuals processed this way into a 80% training set and a 20% validation set. After additionally removing related individuals, there are 112,730 individuals in the training set and 28,545 individuals in the validation set (Supplementary Figure 4).

Hypertension status was determined by medical records using self report, ICD9 codes (401.* and 405.*), and ICD10 codes (I10 and I15.*). Systolic blood pressure (SBP) was determined by automated reading and Data-Field 4080 was used in UK Biobank.

### 4.3 Convolutional VAE model architecture and training

To generate SPINCs, we encode the flow-time and volume-time curves. In our VAE, we use one-dimensional convolutional layers to utilize the temporal context of this time series, encoding the two curves in two channels. In the encoder, we first apply three 1D convolutional layers, each followed by max pooling, and use three fully-connected layers to generate the mean and variance of the bottleneck layer. We use 5 latent dimensions, identical to the number of EDFs, and each latent coordinate is sampled from the Gaussian distribution with the learned means and variances. The decoder architecture is a mirror image of the encoder. We start with three fully-connected layers followed by transpose convolutions layers, each prepended by an upsampling layer. See “SPINCs model architecture” in Supplementary Notes for full details.

For RSPINCs, we encode the flow-volume curve alone, and we apply the same sequences of convolutional and fully-connected layers as we did for SPINCs, while using only 2 latent dimensions in this case. Importantly, we use a novel VAE architecture to concatenate the 5 EDFs directly to the sampled output of the bottleneck layer (the layer right before the decoder) to learn only the residual signals not represented by the EDFs (Figure 2a). As a result, the encoder output dimension is 2 while the decoder input has dimension 5 + 2 = 7. See “RSPINCs model architecture” in Supplementary Notes for full details.

For PLENCs, we encode the PPG curves. In our VAE, we use one-dimensional convolutional layers to utilize the temporal context of this time series. In the encoder, similar to SPINCs, we first apply three 1D convolutional layers, each followed by max pooling, and use three fully-connected layers to generate the mean and variance of the bottleneck layer. We use 5 latent dimensions, identical to the number of EDFs, and each latent coordinate is sampled from the Gaussian distribution with the learned means and variances. The decoder architecture is a mirror image of the encoder. We start with three fully-connected layers followed by transpose convolutions layers, each prepended by an upsampling layer. See “PLENCs model architecture” in Supplementary Notes for full details.

All models are trained using the standard VAE loss function consisting of the reconstruction loss and the (rescaled) Kullback–Leibler (KL) divergence loss. For RSPINCs the KL divergence loss is only applied to the learned encodings, not to the injected EDFs. For optimization, the Adam optimizer [64] is used with varying learning rates and batch sizes. The final learning rate and batch size values (“hyperparameters”) for (R)SPINCs and PLENCs were chosen to minimize the VAE loss in the validation set (Supplementary Notes, Supplementary Table 1).

After training SPINCs, RSPINCs, and PLENCs models, we use the encoders of the trained models to generate the encodings for each individuals, using the mean value of the learned Gaussian distribution of the encodings.

All models were implemented in TensorFlow V2 [65] and XManager (URLs) was used to manage multiple machine learning experiments.

### 4.4 Phenotypic Correlation Analysis

To residualize EDFs and/or covariates from (R)SPINCs and PLENCs, we used ordinary least squares linear regression. To compute the correlation of the EDFs-and-covariates-residualized (R)SPINCs and PLENCs with the tabular fields in UK Biobank, we first preprocessed the tabular fields to remove special codes, normalize, impute and aggregate the values, and finally transformed the categorical fields into one-hot encodings. For each individual correlation analysis between a feature and one of the (R)SPINCs and PLENCs, we computed the Pearson correlation and the *P* -value with two-sided alternative hypothesis.

### 4.5 Survival analysis

We performed analysis of overall survival for European individuals in the spirometry (*n*=65,266) and PPG (*n*=28,545) validation sets using the time from first assessment (field 53) to death from any cause (field 40000). Subjects who were not known to have died were right-censored at the date of UKB data ingestion (Dec 18, 2020). We quantified the association between overall survival and each SPINC, RSPINC, PLENC, and EDF per standard deviation using the hazard ratio, which was estimated from a Cox proportional hazards model adjusting for age and sex. The proportional hazards assumption, with respect to each SPINC, RSPINC, PLENC, and EDF, was assessed using the Schoenfeld residual test. After stratifying patients into quartiles using each SPINC, RSPINC, PLENC, or EDF coordinate, the overall survival curves in Supplementary Figures 7 to 9 were constructed using the standard Kaplan-Meier estimator with bootstrapped 95% confidence intervals.

### 4.6 Genome-wide association studies and polygenic risk scores

GWAS on all spirograms EDFs, SPINCs, and RSPINCs were performed using BOLT-LMM v2.3.6 [37, 38], adjusting for age, sex, age^2^, age *×* sex, height, height^2^, body mass index, smoking status, the number of packs of cigarettes smoked per year, the type of genotyping array, and the top 15 genetic principal components as covariates. GWAS on all PPG EDFs and PLENCs were performed using BOLT-LMM v2.3.6 [37, 38], adjusting for age, sex, age^2^, age *×* sex, height, height^2^, body mass index, the type of genotyping array, and the top 15 genetic principal components as covariates.

All GWAS were restricted to European ancestry individuals to minimize confounding. For quality control we kept variants with minor allele frequency *≥* 0.001, imputation INFO score *≥* 0.8, missing call fraction *≤* 0.05, and Hardy-Weinberg equilibrium *P* -value *≥* 10*^−^*^10^, among all genotyped and imputed variants provided by UK Biobank. After GWAS, we performed S-LDSC [66] to estimate SNP-heritability and detect potential confounding. Genome-wide significant “hits” were defined as the most significant variants with *p ≤* 5 *×* 10*^−^*^8^ and independent at *R*^2^ *<* 0.1 using the PLINK –clump command. A reference panel for linkage disequilibrium (LD) calculation contained 10,000 unrelated European samples from the UKB. Significant “loci” were created based on the span of reference panel SNPs in LD (*R*^2^ *≥* 0.1) with the hits. Loci separated by fewer than 250 kb were subsequently merged.

While performing GWAS, PRSs for all traits were computed using the –predBetasFile option of BOLT-LMM, which generates PRS coefficients using a Bayesian linear mixed model. While GWAS was performed on individuals with valid spirometry measurements, we evaluated the performance of the PRS in a separate set of individuals not used for GWAS.

To determine the known lung function loci from previous literature, we extracted all significant loci from Shrine et al. [25] and searched for lung function-related keywords in the NHGRI-EBI GWAS Catalog (version v1.0.2-associations_e106_r2022-07-09) [39]. We used the following keywords for the Catalog search (case insensitive): “asthma”, “chronic obstructive pulmonary disease”, “copd”, “expiratory flow”, “fev1”, “forced expiratory”, “forced vital capacity”, and “lung function”. To determine the known cardiovascular function loci from previous literature, we used the following keywords for the NHGRI-EBI GWAS Catalog search (case insensitive): “arrhythmia”, “afib”, “atrial fibrillation”, “coronary artery disease”, “stroke”, “heart attack”, “myocardial infarction”, “heart failure”, “mace”, and “rheumatic heart disease”.

### 4.7 Summary statistics conditional and joint analysis

We applied conditional and joint analysis (COJO) on set of previously known loci using GCTA (genome-wide complex trait analysis) software (version `1.93.3beta`) and we set `–cojo-cond` to the set of known loci. We provided 10,000 unrelated European samples randomly chosen from UKB as the reference samples, which is the same reference used to perform LD clumping to define hits, as part of GCTA-COJO input parameters.

### 4.8 PRS evaluation on respiratory diseases and cardiovascular traits on multiple datasets

#### COPDGene dataset

COPDGene is a study of 10,300 current and former smokers with and without COPD, self-reported non-Hispanic White and African-American, without known lung diseases other than COPD and asthma (dbGaP accession phs000179.v6.p2). Additional study details, the study protocol and details of genotyping have been previously published [43, 67], and additionally detailed at copdgene.org. We used the provided variant calls in VCF files and imputed the variants to the Haplotype Reference Consortium (HRC) reference panel using Michigan Imputation Server [68], resulting in 39,127,678 total variants. COPD cases were determined using the GOLD (Global Initiative for Chronic Obstructive Lung Disease) criteria, where GOLD stage 2 or higher was considered as cases. Among 6,576 non-Hispanic White individuals, we had access to 1,131 (17%) asthma cases and 2,781 (42%) COPD cases, and the rest were used as controls. Meanwhile, among 3,140 African-American individuals, 760 (24%) were asthma cases and 802 (26%) were COPD cases. We used the blood pressure measurements and the “high blood pressure” variable included in the dataset to define systolic blood pressure and hypertension traits.

#### EPIC-Norfolk dataset

The European Prospective Investigation into Cancer in Norfolk (EPIC-Norfolk) is a general population-based cohort study of men and women aged 40–79 years living in Norfolk, UK and recruited from general practices between 1993 to 1997. EPIC-Norfolk cohort participants were linked annually to nationally held hospital records and death certificates from 1999 to 2019 using UK National Health Service numbers. COPD was defined as any hospital admission or cause of death coded 490–492, 494–496 (ICD-9) or J40–J44, J47 (ICD-10). Asthma was similarly defined using codes 493 (ICD-9) or J45, J46 (ICD-10). Hypertension was defined using hospital records and death certificates for ICD codes 401.*, 405.* (ICD-9) and I10, I15.* (ICD-10). The systolic blood pressure was determined using the continuous systolic blood pressure from EPIC-Norfolk health examination at baseline, which is the time-point with the highest number of individuals. In a small set of participants who do not have a baseline blood pressure measurement, we used blood pressure measured at the earliest subsequent health examination. Blood pressure was measured at two time points during the examination, with the mean used for analysis.

#### eMERGE III dataset

We utilize five consent groups that does not require IRB approval: General Research Use (GRU), Health/Medical/Biomedical-Genetic Studies, (HBM-GSO), Health/Medical/Biomedical (HMB), Health/Medical/Biomedical (MDS) HMB-MDS, and Health/Medical/Biomedical (PUB, GSO) (HMB-PUB-GSO) (dbGaP accession phs001584.v2.p2). In eMERGE III, we have access to 1,038 asthma cases and 7,250 controls for European ancestry while in the case of African ancestry we have access to 649 asthma cases and 1,367 controls. We used the 39,131,578 variants that are imputed to the HRC reference provided by dbGaP [69]. Asthma and systolic blood pressure traits were defined using the corresponding variables in the dataset. Hypertension was defined using two variables, “CASE_CONTROL_CKD_T2D_HTN” and “CASE_CONTROL_CKD_T2D”, where the individuals in the former group but not in the latter group were defined as the hypertension cases. Note that this limited our analysis to hypertensive individuals without chronic kidney disease or type 2 diabetes.

#### Indiana Biobank dataset

The Indiana Biobank is a state-wide collaboration that provides centralized processing and storage of specimens that are linked to participants’ electronic medical information via Regenstrief Institute at Indiana University. COPD was diagnosed by using ICD9: 491, 492, and 496, and ICD10: J41, J42, J43, and J44. Asthma was diagnosed by using ICD9: 493, and ICD10: J45 and J46. Cases were defined as having at least one in-patient diagnosis or two out-patient diagnoses. Those participants not having any diagnosis were defined as controls. Thus, we have 1,445 COPD cases and 3,808 controls while we have 1,171 asthma cases and 4,083 controls. Among 5,253 individuals for COPD evaluation, 3797 were of European ancestry, 1,371 of African ancestry, and 85 Hispanic ancestry. Among 5,254 individuals for asthma evaluation, 3805 of European ancestry, 1,362 of African ancestry, and 87 of Hispanic ancestry. Indiana Biobank samples used in this study were genotyped using Illumina Infinium Global Screening Array (GSA, Illumina, San Diego, CA) by Regeneron (Tarrytown, NY). SNPs with missing rate *>* 5%, MAF *≤* 1%, HWE *P* -value *<* 1 *×* 10*^−^*^10^ among cases and *<* 1 *×* 10*^−^*^6^ in controls were excluded as reported previously [45]. Genotyping data were imputed to 1000 Genomes using the Michigan Imputation Server [68]. Imputed variants with *R*^2^ *<* 0.30 and MAF*<* 1% were excluded. PLINK [70, 71] was used to calculate PRS by using imputation dosages.

### 4.9 Functional significance of discovered loci

We ran GREAT v4.0.4 [41] on the human GRCh37 assembly to perform functional enrichment analysis of SPINCs, RSPINCs, PLENCs, and EDFs loci. We used the default “basal+extension” region-gene association rule with 5 kb upstream, 1 kb downstream, 1000 kb extension, and curated regulatory domains included. Furthermore, we ran GARFIELD [40] with default parameters to perform tissue-specific analysis where we utilized 424 DNase I hypersensitive site hotspot annotations provided by the GARFIELD authors [40].

### 4.10 Latent causal variable analysis

We applied LCV [51] (URLs) on genome-wide summary statistics for each pair of phenotypes. To create the right input for LCV, we used the munge script provided by S-LDSC software (URLs) to restrict the variants to HapMap3 SNPs with MAF *>* 0.05 and outside the MHC region. We utilized the baseline LD scores for HapMap3 SNPs. A two-sided test was used for the estimated GCP and to compute the significant level.

### 4.11 Genetic phenome-wide association study

To compute PheWAS, we downloaded GWAS summary statistics for 7,221 phenotypes from the Pan-UKB consortium 20200615 release (URLs). After restricting to phenotypes that contained European statistics and did not persistently fail in LD clumping, we were left with 7,145 pruning+thresholding PRSs generated by PLINK (URLs) using the –clump command with an index variant significance threshold of 5 *×* 10*^−^*^8^ and LD threshold of 0.1, with LD computed from a random subset of 10,000 European individuals in UK Biobank.

SPINCs, RSPINCs, PLENCs pruning+thresholding PRSs were computed analogously to the Pan-UKB PRSs. We computed the Pearson correlations between the PRSs derived from latent dimensions with the PRSs derived from Pan-UKB phenotypes and the *P* -values with two-sided alternative hypothesis.

## URLs

Baseline and BaselineLD annotations: https://data.broadinstitute.org/alkesgroup/ldscore

BOLT-LMM software: https://data.broadinstitute.org/alkesgroup/bolt-lmm

COPDGene study: https://www.ncbi.nlm.nih.gov/projects/gap/cgi-bin/study.cgi?study_id=phs000179.v6.p2

GCTA software: https://github.com/jianyangqt/gcta

GREAT software: http://great.stanford.edu

GWAS Catalog: https://www.ebi.ac.uk/gwas/

Indiana Biobank study: https://indianabiobank.org/

Pan-UK Biobank GWAS: https://pan.ukbb.broadinstitute.org

PLINK software: https://www.cog-genomics.org/plink1.9

TensorFlow: https://www.tensorflow.org

UCSC LiftOver: https://genome.ucsc.edu/cgi-bin/hgLiftOver

UK Biobank study: https://www.ukbiobank.ac.uk

XManager: https://github.com/deepmind/xmanager

Michigan Imputation Server https://imputationserver.sph.umich.edu/index.html#!pages/home

### Data & code availability

Open-source code and trained model weights are available at https://github.com/Google-Health/ genomics-research under the regle directory. SPINCs, RSPINCs, and PLENCs values of UK Biobank individuals will be returned to UK Biobank and will be made available by UK Biobank. GWAS summary statistics for SPINCs, RSPINCs, PLENCs, and EDFs are freely available on Google Cloud Storage at https://console.cloud.google.com/storage/browser/brain-genomics-public/ research/regle.

## Supporting information

Large Supplementary Tables

## Data Availability

Datasets used in this study (UK Biobank, COPDGene, eMERGE III, EPIC Norfolk, and Indiana Biobank) are available to qualified researchers via applying access to each dataset maintainers.
All results derived from UK Biobank by this study will be returned to UK Biobank and will be made available by UK Biobank maintainers.
Open-source code used for this study is available on GitHub at https://github.com/Google-Health/genomics-research under "regle" directory.
GWAS summary statistics are freely available on Google Cloud Storage at https://console.cloud.google.com/storage/browser/brain-genomics-public/research/regle

https://console.cloud.google.com/storage/browser/brain-genomics-public/research/regle

## Acknowledgments

We thank all participants, dataset creators, and maintainers of UK Biobank, COPDGene, eMERGE III, EPIC Norfolk, and Indiana Biobank. Full acknowledgement for each dataset can be found in “Dataset acknowledgment” in Supplementary Notes. We also thank Nick Furlotte for helpful discussions.

M.H.C. was supported by NHLBI R01HL153248, R01HL149861, and R01HL147148. The content is solely the responsibility of the authors and does not necessarily represent the official views of the NIH. The funding body has no role in the design of the study and collection, analysis, and interpretation of data and in writing the manuscript.

