## Supplementary Materials for "Unsupervised representation learning improves genomic discovery and risk prediction for respiratory and circulatory functions and diseases"

[*] Joint supervision.
[1] Google Research, Cambridge, MA 02142, USA.
[2] Google Research, Palo Alto, CA 94304, USA.
[3] Department of Electrical and Computer Engineering, Northeastern University, Boston, MA 94304, USA.
[4] Channing Division of Network Medicine, Brigham and Women's Hospital, Boston, MA 02115, USA.
[5] NIHR Biomedical Research Centre at Moorfields Eye Hospital & UCL Institute of Ophthalmology, London EC1V 9EL, UK.
[6] MRC Epidemiology Unit, University of Cambridge, Cambridge CB2 0SL, UK.
[7] Department of Medical and Molecular Genetics, Indiana University School of Medicine, Indianapolis, IN 46202, USA.
[8] Verily Life Sciences, South San Francisco, CA 94080, USA.
[9] Division of Cardiology, Department of Medicine, Indiana University School of Medicine, Indianapolis, IN 46202, USA.
[10] Division of Pulmonary and Critical Care Medicine, Brigham and Women's Hospital, Boston, MA 02115, USA.
[11] Harvard Medical School, Boston, MA 02115, USA.
[◇] Present address: Insitro, South San Francisco, CA 94080, USA.

### Supplementary Notes

#### (R)SPINCs reconstruct spirograms with few latent dimensions

The quality of the reconstruction from our models improves as we increase the latent dimension (i.e. the number of coordinates in SPINCs or RSPINCs) (Figure 2b) and we observed highly accurate reconstruction of the input spirograms with just five SPINCs (Figure 2c), identical to the number of EDFs we consider. In comparison, spirogram reconstructions from PCA with five PCs resulted in a $2\times$ higher reconstruction error, which indicates SPINCs and RSPINCs encoded more information about spirograms than PCs with the same number of coordinates (Figure 2b). To have a fair comparison, SPINCs model and PCA were trained and evaluated on the same set of datasets. We observed a similar pattern of reconstruction errors using RSPINCs (Supplementary Figure 2), though note that they are not directly comparable due to the inclusion of EDFs.

#### (R)SPINCs are consistent across random weight initializations

The objective function used to train deep learning models is typically not convex, so training runs with different initialization of weights can converge to substantially different local minima. To assess the stability of our encodings to unimportant changes in training, we generated five sets of SPINCs (dim=5) using different random seeds for initialization of weights, and computed Pearson correlation of the coordinates of each set with the coordinates of all other sets. Up to a change of signs and a permutation of the coordinates, which have no significance in our model or its downstream applications, we observed that the learned encodings are highly consistent (Supplementary Figure 3).

#### (R)SPINCs GWAS

We generated SPINCs (dim=5) and RSPINCs (dim=2, in addition to 5 EDFs) for all individuals with valid first-visit spirograms in UK Biobank (Supplementary Figures 1, 10 and 11; Methods). Then, we performed GWAS on all European-ancestry individuals ($n$=324,702) on all encoding coordinates and 5 EDFs using BOLT-LMM [1, 2], adjusting for age, sex, age$^2$, age $\times$ sex, height, height$^2$, body mass index, smoking status, pack-years of smoking, the type of genotyping array, and the top 15 genetic principal components (Methods). The Manhattan plots of 5 SPINCs and 2 RSPINCs GWAS are illustrated in Supplementary Figures 12 to 16 and Supplementary Figures 17 and 18, respectively. The intercept term from the stratified linkage disequilibrium score regression (S-LDSC) [3] was close to 1 (Supplementary Table 10) for the GWAS of SPINCs and RSPINCs, indicating minimal confounding bias. The SNP-heritability estimated from S-LDSC for SPINCs and RSPINCs showed strong genetic components (Supplementary Table 10). For comparison, we also performed GWAS on the first 5 PCs of the raw spirograms following the same steps.

#### PLENCs GWAS

We generated PLENCs (dim=5) for all individuals with valid first-visit PPG in UK Biobank (Methods). Then, we performed GWAS on all European-ancestry individuals ($n$=141,275) on all encoding coordinates and 5 EDFs using BOLT-LMM [1, 2], adjusting for age, sex, age$^2$, age $\times$ sex, height, height$^2$, body mass index, the type of genotyping array, and the top 15 genetic principal components (Methods). The Manhattan plots of 5 PLENCs GWAS are illustrated in Supplementary Figures 19 to 23. The intercept term from the stratified linkage disequilibrium score regression (S-LDSC) [3] was close to 1 (Supplementary Table 10) for the GWAS of PLENCs, indicating minimal confounding bias. The SNP-heritability estimated from S-LDSC for PLENCs showed strong genetic components (Supplementary Table 10). For comparison, we also performed GWAS on the first 5 PCs of the raw PPG following the same steps.

#### Functional significance of discovered loci

We ran GREAT and GARFIELD for functional enrichment. For GREAT, we combined GWS loci from SPINCs GWAS by merging those within 250 kb of each other and analyzed the resulting set of independent regions (n=575), and performed the analogous analysis for EDFs (n=613) and EDFs+RSPINCs (n=659). The strongest consistent enrichments were Gene Ontology terms related to development and morphogenesis (Supplementary Table 27). Notably, significant enrichments for EDFs were largely found to be even more significantly enriched in the EDFs+RSPINCs, consistent with the RSPINCs identifying additional genes influencing the same biological pathways as EDFs (Supplementary Figure 31, $P = 2.4 \times 10^{-25}$, two-sided paired $t$-test).

In addition, using GARFIELD to test the enrichment of SPINCs and RSPINCs GWAS with DNase I hypersensitive hotspots, we observed a strong enrichment of SPINCs and RSPINCs in fetal lung (Supplementary Figures 24 to 28 and Supplementary Figures 29 and 30). Notably, we found a strong enrichment for RSPINC$_2$ in blood (Supplementary Figure 30).

### (R)SPINCs are associated with overall survival

We performed survival analysis for European individuals in the validation set ($n = 65,266$) across EDFs, SPINCs, and RSPINCs, fitting a Cox proportional hazards regression model to UKB death registry data while controlling for age and sex as covariates (Methods and Supplementary Table 9). The EDF and $SPINC_3$ hazard ratios (HR) for all cause mortality (i.e., overall survival) were 0.640 (95% CI $[0.615, 0.666]$), 0.679 ($[0.653, 0.77]$), 0.685 ($[0.656, 0.715]$), 0.689 ($[0.663, 0.716]$), 0.752 ($[0.728, 0.777]$), and 0.806 ($[0.786, 0.826]$) per one standard deviation increase in $FEV_1$, $SPINC_3$, FVC, $FEF_{25\text{-}75\%}$, PEF, and $FEV_1$/FVC, respectively, suggesting that these features are strongly to moderately associated with improved survival or longer time-to-death. Conversely, the $RPINC_2$, $SPINC_1$, $SPINC_2$, and $SPINC_4$ HRs of 1.064 (95% CI $[1.032, 1.097]$), 1.078 ($[1.045, 1.111]$), 1.086 ($[1.052, 1.121]$), and 1.135 ($[1.101, 1.169]$) per standard deviation increase suggesting that these features are moderately associated with shorter time-to-death. Note that the $SPINC_1$ model fails the proportional-hazards (PH) assumption ($p = 0.013$). Thus, the estimated hazard ratio is time-varying, and the reported value should be interpreted as giving the net direction of association. Neither $SPINC_5$ nor $RSPINC_1$ are significantly associated with survival. Kaplan-Meier curves for overall survival (OS) stratified by feature indicate that OS declines more rapidly for patients with higher $SPINC_1$ and lower $SPINC_3$ scores (Supplementary Figures 7 and 8).

### (R)SPINCs are genetically causally associated with asthma and COPD

To identify potentially causal relationships between (R)SPINCs, EDFs, and lung diseases, we performed latent causal variable (LCV) analysis on the traits. LCV assumes that a latent causal variable mediates the genetic correlation between two traits, and a trait A is said to be "partially genetically causal" for trait B if trait A is strongly genetically correlated with the latent causal variable for the two traits. The genetic causality proportion (GCP) (of trait A on trait B) is defined to quantify this partial causality, where GCP $= 0$ implies no partial genetic causality and GCP $= 1$ implies "full" genetic causality (i.e. the entire genetic component of trait A is causal for trait B).

We observed that the GCP of SPINCs on COPD and its significance (the highest GCP is $0.82 \pm 0.14$ from the second coordinate of SPINCs with $P = 10^{-6.7}$) is comparable to the GCP of EDFs on COPD (the highest GCP is $0.84 \pm 0.13$ from $FEV_1$/FVC with $P = 10^{-6.7}$) (Supplementary Table 26). We note that a high GCP of $FEV_1$/FVC on COPD is expected since it is the main metric to define COPD, and SPINCs seem to capture the equivalent amount of GCP for COPD.

For asthma, we found that the direction of partial genetic causality was the opposite, implying that asthma was partially genetically causal for both SPINCs and EDFs under the LCV model (i.e. the latent causal variable was more correlated with asthma than it was with SPINCs or EDFs). We observed an extremely significantly high GCP of asthma on the fifth coordinate of SPINCs (GCP = $0.71 \pm 0.12$, $P = 10^{-42.1}$), while the most significant GCP of asthma on the EDFs was much lower (the highest GCP is $0.43 \pm 0.10$ from $\text{FEV}_1$, $P = 10^{-7.2}$) (Supplementary Table 26). These findings may be consistent with many subjects with asthma having normal lung function, as defined by EDFs.

Finally, we applied LCV to a set of phenotypes (sarcoidosis, systemic lupus erythematosus, thyroid dysfunction, and diets consistent with celiac disease) where we observed significant correlation between their PRSs obtained from PRS PheWAS (Supplementary Table 26). We observed significantly high GCP for sarcoidosis with the third coordinate of SPINCs, lupus with second coordinate of SPINCs and first coordinate of RSPINCs, thyrotoxicosis with third SPINC coordinate, and gluten free diet with third SPINCs coordinate (Supplementary Table 25). Notably, for all these phenotypes, we observed a direction of effect from these phenotypes to SPINCs and RSPINCs.

### SPINCs model architecture

```
Encoder:
=================================================================
 Layer (type)                Output Shape              Param #
=================================================================
 vae_encoder_input (InputLayer) [(None, 1000, 2)]     0
 conv1d (Conv1D)             (None, 1000, 8)           168
 max_pooling1d (MaxPooling1D)  (None, 500, 8)          0
 conv1d_1 (Conv1D)           (None, 500, 16)           1296
 max_pooling1d_1 (MaxPooling1D) (None, 250, 16)        0
 conv1d_2 (Conv1D)           (None, 250, 32)           5152
 max_pooling1d_2 (MaxPooling1D) (None, 125, 32)        0
 flatten (Flatten)           (None, 4000)              0
 dense (Dense)               (None, 64)                256064
 dense_1 (Dense)             (None, 64)                4160
 dense_2 (Dense)             (None, 64)                4160
```

```
 z_mean (Dense)              (None, 5)           325

 z_log_var (Dense)           (None, 5)           325

 gaussian_sampling           (None, 5)           0
 (GaussianSampling)
==============================================================
Total params: 271,650
Trainable params: 271,650

Decoder:
=================================================================
 Layer (type)                Output Shape              Param #
=================================================================
 vae_decoder_input           [(None, 5)]               0
 (InputLayer)

 dense_3 (Dense)             (None, 64)                384

 dense_4 (Dense)             (None, 64)                4160

 dense_5 (Dense)             (None, 64)                4160

 dense_6 (Dense)             (None, 4000)              260000

 reshape (Reshape)           (None, 125, 32)           0

 up_sampling1d (UpSampling1D) (None, 250, 32)          0

 conv1d_transpose            (None, 250, 16)           5136
 (Conv1DTranspose)

 up_sampling1d_1             (None, 500, 16)           0
 (UpSampling1D)

 conv1d_transpose_1          (None, 500, 8)            1288
 (Conv1DTranspose)

 up_sampling1d_2             (None, 1000, 8)           0
 (UpSampling1D)

 conv1d_transpose_2          (None, 1000, 2)           162
 (Conv1DTranspose)

=================================================================
Total params: 275,290
```

```
Trainable params: 275,290
```

#### RSPINCs model architecture

```
Encoder:
=================================================================
 Layer (type)                Output Shape              Param #
=================================================================
 vae_encoder_input (InputLayer) [(None, 1000, 1)]     0
 conv1d (Conv1D)             (None, 1000, 8)           88
 max_pooling1d (MaxPooling1D)   (None, 500, 8)         0
 conv1d_1 (Conv1D)           (None, 500, 16)           1296
 max_pooling1d_1 (MaxPooling1D) (None, 250, 16)        0
 conv1d_2 (Conv1D)           (None, 250, 32)           5152
 max_pooling1d_2 (MaxPooling1D) (None, 125, 32)        0
 flatten (Flatten)           (None, 4000)              0
 dense (Dense)               (None, 64)                256064
 dense_1 (Dense)             (None, 64)                4160
 dense_2 (Dense)             (None, 64)                4160
 z_mean (Dense)              (None, 2)                 130
 z_log_var (Dense)           (None, 2)                 130
 gaussian_sampling           (None, 2)                 0
 (GaussianSampling)
=================================================================
Total params: 271,180
Trainable params: 271,180

Concatenate (inject 5 traditional measurements into encoder output):
=================================================================
 Layer (type)                Output Shape              Param #
=================================================================
 concatenate (Concatenate)   (None, 7)                 0
```

```
Decoder:
=================================================================
 Layer (type)                Output Shape              Param #
=================================================================
 vae_decoder_input           [(None, 7)]              0
 (InputLayer)
 dense_3 (Dense)             (None, 64)                512
 dense_4 (Dense)             (None, 64)                4160
 dense_5 (Dense)             (None, 64)                4160
 dense_6 (Dense)             (None, 4000)              260000
 reshape (Reshape)           (None, 125, 32)           0
 up_sampling1d               (None, 250, 32)           0
 (UpSampling1D)
 conv1d_transpose            (None, 250, 16)           5136
 (Conv1DTranspose)
 up_sampling1d_1             (None, 500, 16)           0
 (UpSampling1D)
 conv1d_transpose_1          (None, 500, 8)            1288
 (Conv1DTranspose)
 up_sampling1d_2             (None, 1000, 8)           0
 (UpSampling1D)
 conv1d_transpose_2          (None, 1000, 1)           81
 (Conv1DTranspose)
=================================================================
Total params: 275,337
Trainable params: 275,337
```

#### PLENCs model architecture

```
Encoder:
=================================================================
 vae_encoder_input (InputLayer)  [(None, 100, 1)]       0
 conv1d (Conv1D)           (None, 100, 8)            88
```

```
 max_pooling1d (MaxPooling1D)  (None, 50, 8)          0

 conv1d_1 (Conv1D)         (None, 50, 16)           1296

 max_pooling1d_1 (MaxPooling1D)  (None, 25, 16)         0

 flatten (Flatten)         (None, 400)              0

 dense (Dense)             (None, 64)               25664

 dense_1 (Dense)           (None, 64)               4160

 dense_2 (Dense)           (None, 64)               4160

 z_mean (Dense)            (None, 5)                325

 z_log_var (Dense)         (None, 5)                325

 gaussian_sampling (Gaussian Sampling)  (None, 5)              0

=================================================================


Decoder:

=================================================================

 vae_decoder_input (InputLayer)  [(None, 5)]            0

 dense_3 (Dense)           (None, 64)               384

 dense_4 (Dense)           (None, 64)               4160

 dense_5 (Dense)           (None, 64)               4160

 dense_6 (Dense)           (None, 400)              26000

 reshape (Reshape)         (None, 25, 16)           0

 up_sampling1d (UpSampling1D)  (None, 50, 16)         0

 conv1d_transpose (Conv1DTranspose)  (None, 50, 8)          1288

 up_sampling1d_1 (UpSampling1D)  (None, 100, 8)         0

 conv1d_transpose_1 (Conv1DTranspose)  (None, 100, 1)         81

=================================================================


Total params: 72,091

Trainable params: 72,091
```

### Dataset acknowledgment

#### UK Biobank dataset acknowledgment

This research has been conducted using the UK Biobank Resource under Application Number 65275.

#### COPDGene dataset acknowledgment

This research used data generated by the COPDGene study, which was supported by NIH grants U01 HL089856 and U01 HL089897. The COPDGene project is also supported by the COPD Foundation through contributions made by an Industry Advisory Board comprised of Pfizer, AstraZeneca, Boehringer Ingelheim, Novartis, and Sunovion.

#### EPIC Norfolk dataset acknowledgment

The EPIC-Norfolk study (DOI 10.22025/2019.10.105.00004) has received funding from the Medical Research Council (MR/N003284/1 MC-UU_12015/1 and MC_UU_00006/1) and Cancer Research UK (C864/A14136). The genetics work in the EPIC-Norfolk study was funded by the Medical Research Council (MC_PC_13048). We are grateful to all the participants who have been part of the project and to the many members of the study teams at the University of Cambridge who have enabled this research.

#### Indiana Biobank dataset acknowledgment

This study was made possible, in part, with support from the Indiana Clinical and Translational Sciences Institute funded, in part by Award Number UL1TR002529 from the National Institutes of Health, National Center for Advancing Translational Sciences, Clinical and Translational Sciences Award, and the National Center for Research Resources, Construction grant number RR020128 and the Lilly Endowment. The content is solely the responsibility of the authors and does not necessarily represent the official views of the National Institutes of Health. The authors acknowledge the Indiana University Pervasive Technology Institute for providing [HPC (Big Red II, Karst, Carbonate), visualization, database, storage, or consulting] resources that have contributed to the research results reported within this paper.

**eMERGE III dataset acknowledgment**

This research used data generated by the eMERGE III study which was obtained from dbGaP under accession phs001584.v2.p2. See Supplementary Note for full acknowledgement for eMERGE III.

Cincinnati Children's Hospital Medical Center (CCHMC) – Acknowledgement Text: CCHMC is a participating pediatric institution for Phase III of the eMERGE network, a national consortium selected to expand best practices and knowledge in effective implementation of genomic medicine to pursue a broad-based program sufficiently large to define health outcomes associated with rare variants in ~100 clinically relevant genes. CCHMC Principal Investigators (PIs) have contributed sequencing data generated from the following cohorts: (1) Better Outcomes for Children (BOfC). Since January, 2011, the Cincinnati Biobank has managed the sample repository developed through the BOfC protocol (PI: John Harley), an institution-wide broad based consent project to utilize remnant clinical samples for biomedical research from participants consented at registration. This project is made possible by institutional resources. To date, over 261,000 participants have consented to BOfC and DNA samples are stored from more than 84,000 unique patients. Through an IRB approved protocol led by Dr. Bahram Namjou (2015-7778), 2,800 samples were selected for sequencing on the eMERGE sequencing panel representing >15 primary phenotypes including Arrhythmia, Asthma, Cardiomyopathy, Chronic kidney disease, Ehlers-Danlos Syndrome, Hyperlipidemia, Autistic behavior, and Tuberous Sclerosis 1. This project is made possible by the support of U01HG008666 (PI: John Harley). (2) Return of eMERGE III Genomic Results. Through an IRB approved protocol led by Dr. Melanie Myers (2016-3361), 200 adolescent patients and their parents were consented to examine (1) their choices about results to be returned on the eMERGE sequencing panel, (2) their responses to learning negative genetic test results, and (3) the parents' responses after learning their children's positive results. All 200 participants provided blood samples. Extracted DNA samples were sequenced on the eMERGE sequencing panel. Results are to be returned to participants. This project is made possible by the support of U01HG008666 (PI: John Harley). Patients of interest were identified using anthropometric measurements, clinical data and ICD codes extracted from the EPIC electronic medical record (EMR). The extraction of data from the EMR into the de-identified data warehouse, i2b2, was made possible by institutional resources and UL1RR026314/UL1TR001425, the Cincinnati Center for Clinical and Translational Sciences and Training Grant (PI: James Heubi). Children's Hospital of Philadelphia (CHOP) Center for Applied Genomics, The Children's Hospital of Philadelphia Samples and associated genomic and phenotype data used in this study were provided by the Center for Applied Genomics at the Children's Hospital of Philadelphia (CHOP). Support for genotyping was provided by an Institutional Development Award from CHOP. Support for sequencing was provided by the National Institutes of Health through an award from the National Human Genome Research Institute's Electronic Medical Records and Genomics (eMERGE) program (U01HG008684). Columbia University Samples and data used in this study were provided by the Center for Glomerular Diseases at Columbia University, the Columbia Transplant Programs, the DataBase Shared Resource at the Herbert Irving Comprehensive Cancer Center, and the Institute for Genomic Medicine at Columbia University. Funding support for the Columbia eMERGE III research study was provided by a U01 grant from the National Human Genome Research Institute (U01HG008680; PIs – Chunhua Weng, PhD; George Hripcsak, MD; Ali Gharavi, MD). Geisinger Funding for the MyCode® sample and data collection was provided by grants from Commonwealth of Pennsylvania, the Clinic Research Fund of Geisinger Clinic, and the Regeneron Genetics Center. Partners Health Care (Harvard University) Samples and data used in this study were provided by the Partners Health Care Biobank (https://biobank.partners.org/). Funding support for the Partners Biobank was provided by Partners Health Care and Partners Personalized Medicine. Assistance with phenotype harmonization was provided by the eMERGE Coordinating Center (Grant number U01HG04603). Additional support was provided by the NIH, NHGRI eMERGE Network (U01HG 5U01HG008685-03). Funding support for genotyping, which was performed at the Translational Genomics Core, Partners Personalized Medicine and funded by Partners Personalized Medicine. Assistance with phenotype harmonization and genotype data cleaning was provided by the eMERGE Administrative Coordinating Center (U01HG004603) and the National Center for Biotechnology Information (NCBI). The datasets used for the analyses described in this manuscript were obtained from dbGaP at http://www.ncbi.nlm.nih.gov/gap through dbGaP accession number; phs000944.v1.p1. Kaiser Washington/University of Washington Funding support for Alzheimer's Disease Patient Registry (ADPR) and Adult Changes in Thought (ACT) study was provided by a U01 from the National Institute on Aging (Eric B. Larson, PI, U01AG006781). A gift from the 3M Corporation was used to expand the ACT cohort. DNA aliquots sufficient for GWAS from ADPR Probable AD cases, who had been enrolled in Genetic Differences in Alzheimer's Cases and Controls (Walter Kukull, PI, R01 AG007584) and obtained under that grant, were made available to eMERGE without charge. Funding support for genotyping, which was performed at Johns Hopkins University, was provided by the NIH (U01HG004438). Genome-wide association analyses were supported through a Cooperative Agreement from the National Human Genome Research Institute, U01HG004610 (Eric B. Larson, PI). Assistance with phenotype harmonization and genotype data cleaning was provided by the eMERGE Administrative Coordinating Center (U01HG004603) and the National Center for Biotechnology Information (NCBI). The datasets used for the analyses described in this manuscript were obtained from dbGaP at http://www.ncbi.nlm.nih.gov/gap through dbGaP accession number phs000234.v1.p1. Mayo Clinic Samples and associated genotype and phenotype data used in this study were provided by the Mayo Clinic. Funding support for the Mayo Clinic was provided through a cooperative agreement with the National Human Genome Research Institute (NHGRI), Grant #: UOIHG004599, UOIHG006379; and the Mayo Center for Individualized Medicine. Funding support for sequencing, which was performed at The Baylor Human Genomics Sequencing Center, was provided by the NIH. Assistance with phenotype harmonization and genotype data cleaning was provided by the eMERGE Administrative Coordinating Center and the National Center for Biotechnology Information (NCBI). Northwestern University Samples and data used in this study were obtained from patients of Northwestern Medicine, Chicago, IL, who were recruited for the eMERGE II Pharmacogenomics Study and the eMERGE III Your Genes and Your Health Study. The Pharmacogenomics Study, a supplement to the Northwestern eMERGE II Project (U01HG006388) and the Your Genes and Your Health Study (U01HG008673) were funded through the NIH, NHGRI eMERGE Network. Vanderbilt University Funding support for the Vanderbilt Genome-Electronic Records (VGER) project was provided through a cooperative agreement (U01HG008672) with the National Human Genome Research Institute (NHGRI) with additional funding from the National Institute of General Medical Sciences (NIGMS). The dataset(s) used for the analyses described were obtained from Vanderbilt University Medical Center. Assistance with phenotype harmonization and genotype data cleaning was provided by the eMERGE Administrative Coordinating Center (U01HG004603) and the National Center for Biotechnology Information (NCBI). The datasets used for the analyses described in this manuscript were obtained from dbGaP at http://www.ncbi.nlm.nih.gov/gap through dbGaP accession number phs000188.v1.p1.

### Supplementary Figures

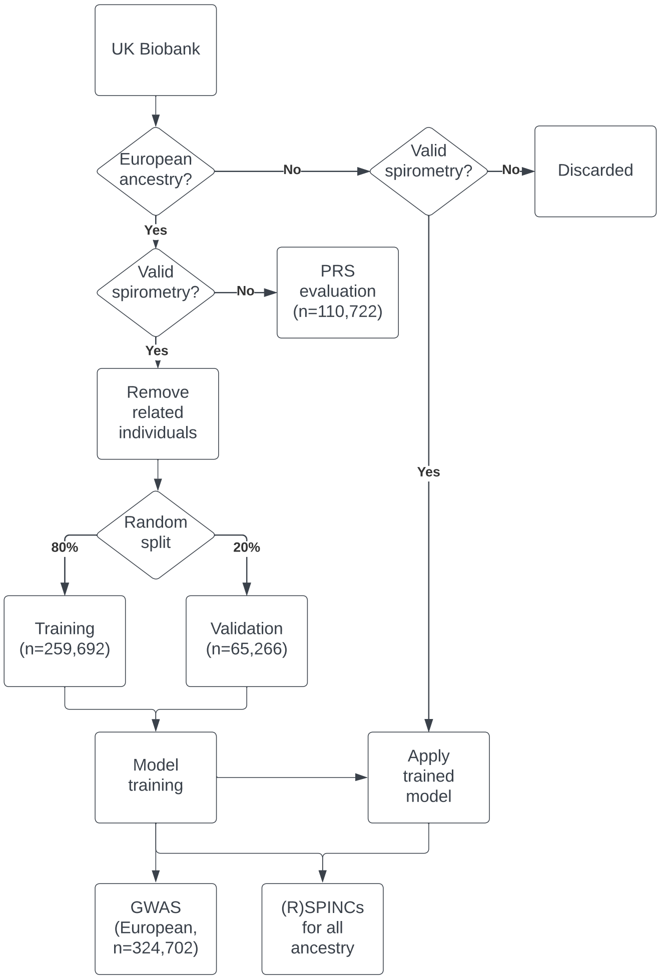


Supplementary Figure 1: **An overview of UK Biobank spirograms used in this study.** Our initial dataset consists of all European-ancestry in UK Biobank ($n$=435,766). We considered all individuals with valid spirograms as modeling dataset ($n$=325,027) and individuals with invalid spirograms are used as PRS holdout set. The PRS holdout set is from the European individuals who are not used in the ML modeling and in the GWASs ($n$=110,739). We split the ML modeling set to training (80%) and validation (20%) sets. We use all individuals in modeling set for GWAS analysis and generated (R)SPINCs for individuals with valid spirometry in all ancestry.

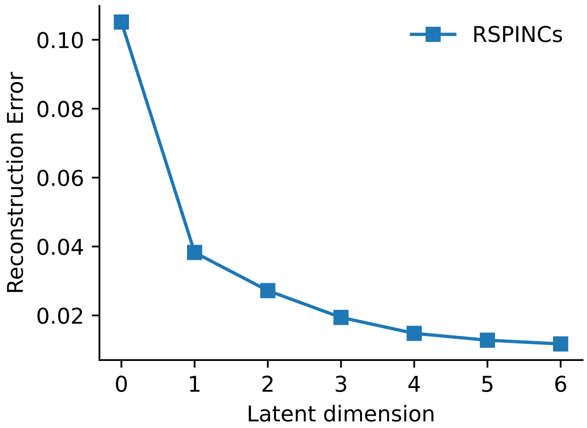


Supplementary Figure 2: **Reconstruction error using RSPINCs with varying latent dimension.** Note that all RSPINCs models include spirogram EDFs (dim = 5), so the total number of inputs used for reconstructing curves is 5 plus the latent dimension. The latent dimension of zero in the plot implies bypassing the encoder and the sampling layer of the VAE and solely using EDFs to reconstruct spirograms.

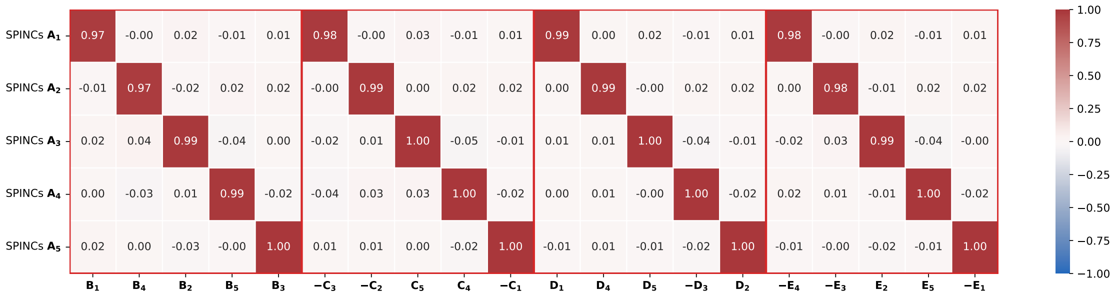


Supplementary Figure 3: **SPINCs trained with different random seeds**. Five SPINCs are trained from an identical model using different random seeds to initialize the training: model **A**, **B**, **C**, **D**, and **E**. The Pearson correlations between the coordinates of the model **A** and the coordinates of the models **B**, **C**, **D**, and **E** are displayed as a heatmap. Note the order and signs of the coordinates of models **B**, **C**, **D**, and **E** are permuted and flipped as indicated in their $x$-axis labels to maximize correlation with coordinates of model **A**.

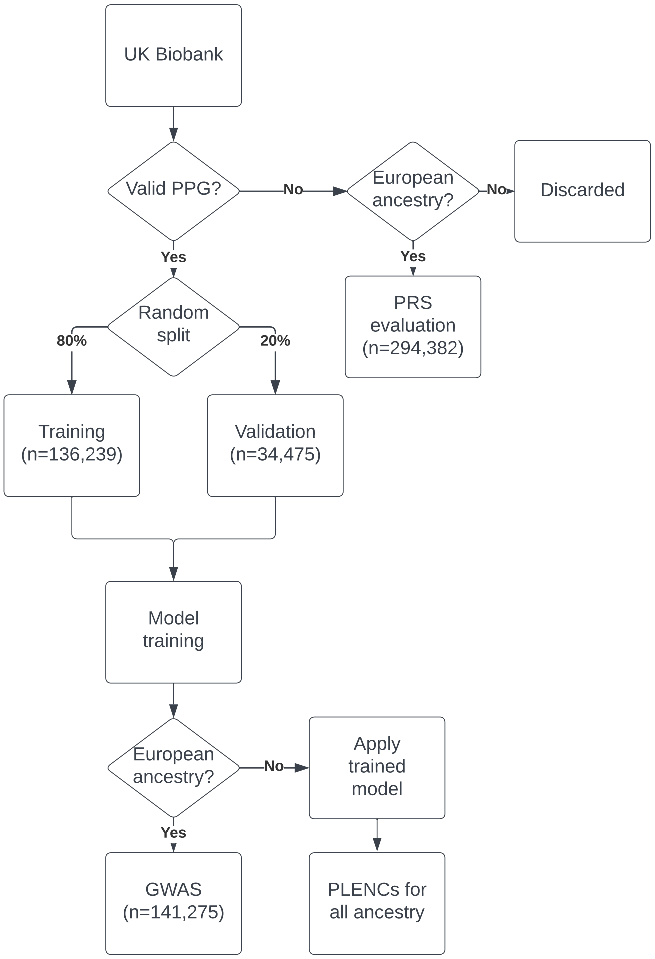


Supplementary Figure 4: **An overview of UK Biobank PPGs used in this study.** We considered all individuals with PPGs as modeling dataset ($n$=170,714) and individuals with invalid spirograms are used as PRS holdout set. The PRS holdout set is from the European individuals who are not used in the ML modeling. We split the ML modeling set to training (80%) and validation (20%) sets. We use all European ancestry individuals in modeling set for GWAS analysis and generated PLENCs for individuals with valid PPGs in all ancestry.

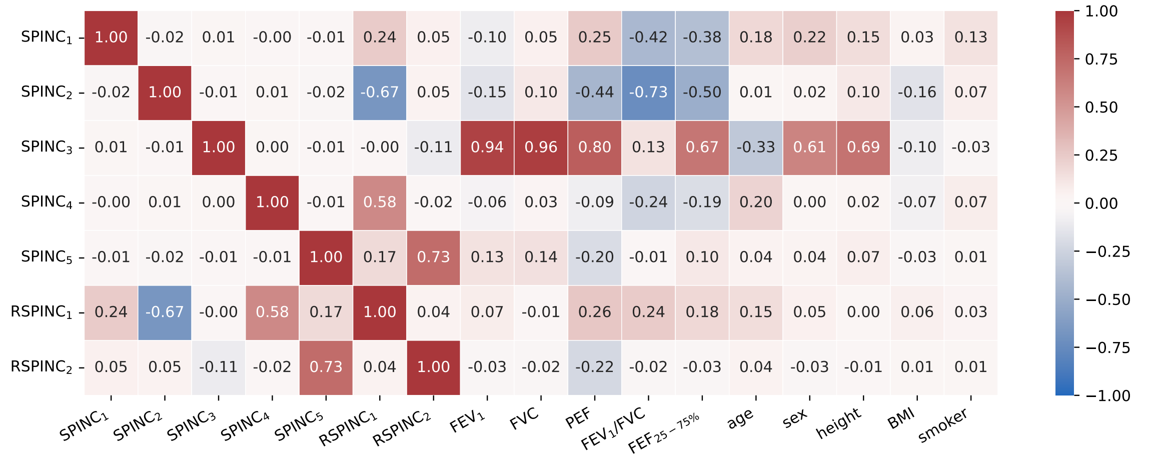


Supplementary Figure 5: **Correlation between SPINCs & RSPINCs coordinates and manual metrics and covariates**. Pearson correlation between the coordinates of SPINCs (dim=5), RSPINCs (dim=2), and the manual spirometry metrics (e.g. $FEV_1$) and other covariates (e.g. age, sex).

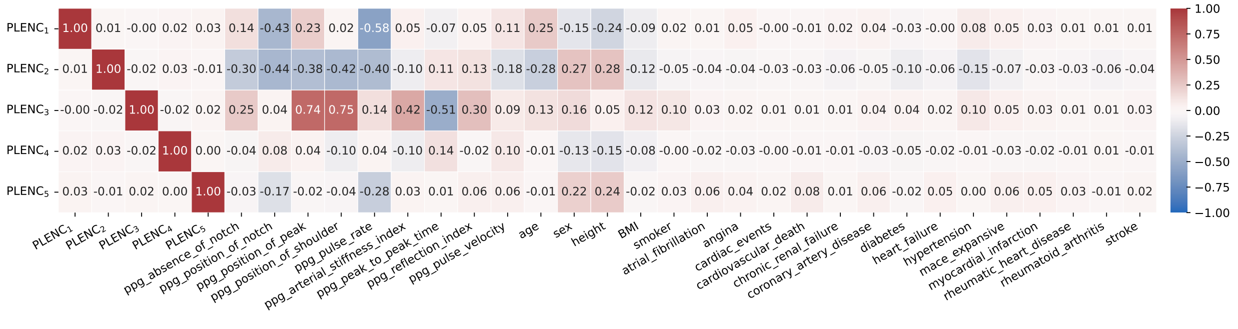


Supplementary Figure 6: **Correlation between PLENCs coordinates and manual metrics, covariates, and cardiovascular diseases**. Pearson correlation between the coordinates of PLENCs (dim=5) and the manual PPG metrics (e.g. notch position) and other covariates (e.g. age, sex).

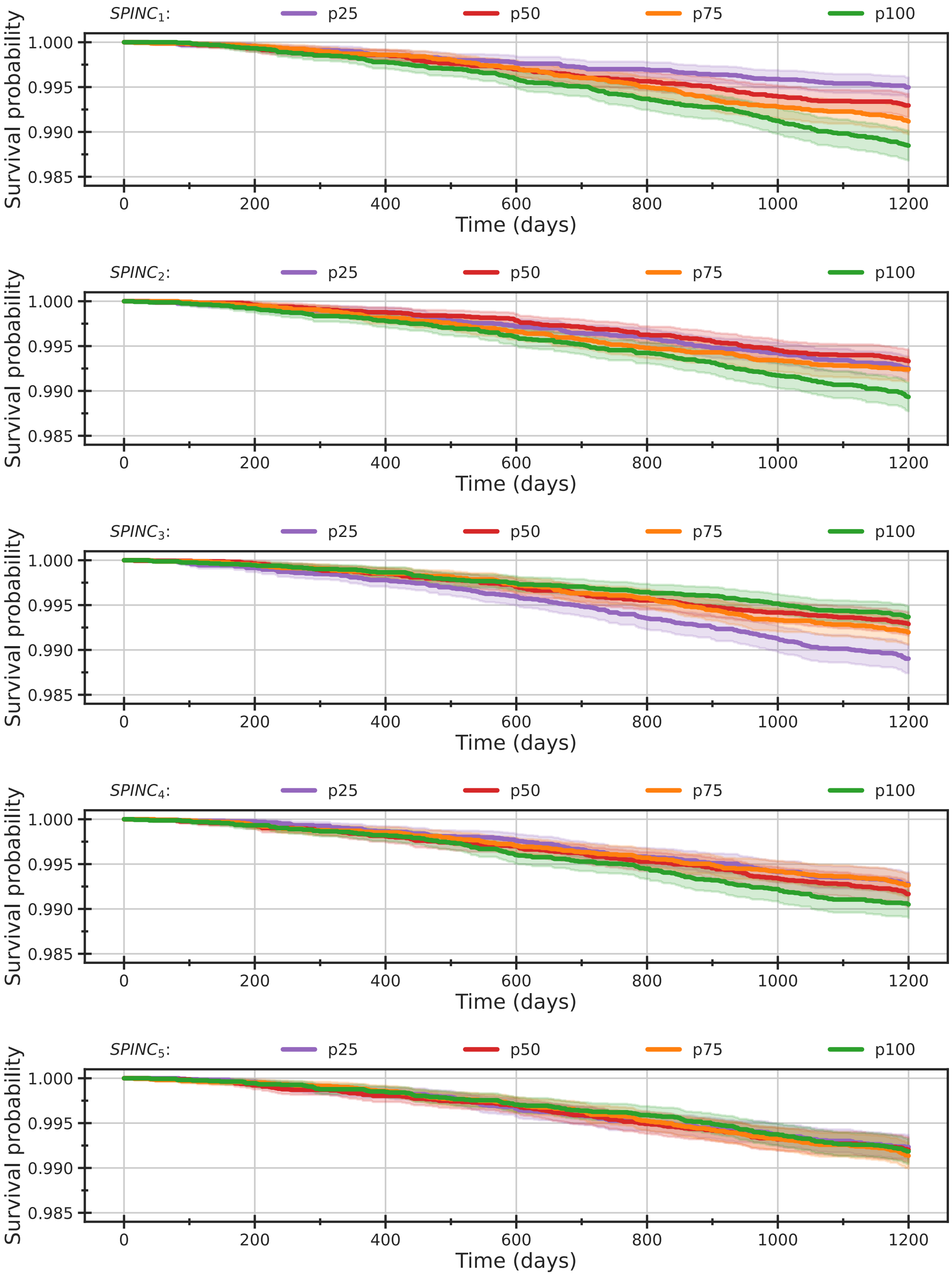


Supplementary Figure 7: **SPINCs Kaplan-Meier curves**. Kaplan-Meier curves estimating the overall survival (OS) function for European individuals in the validation dataset ($n$=65,266). Individuals were stratified into quartiles using each SPINC coordinate (e.g., "p25" denotes the bottom quartile) and OS curves were constructed using the standard Kaplan-Meier estimator with bootstrapped 95% confidence intervals. See Supplementary Table 9 for the corresponding hazard ratios per standard deviation.

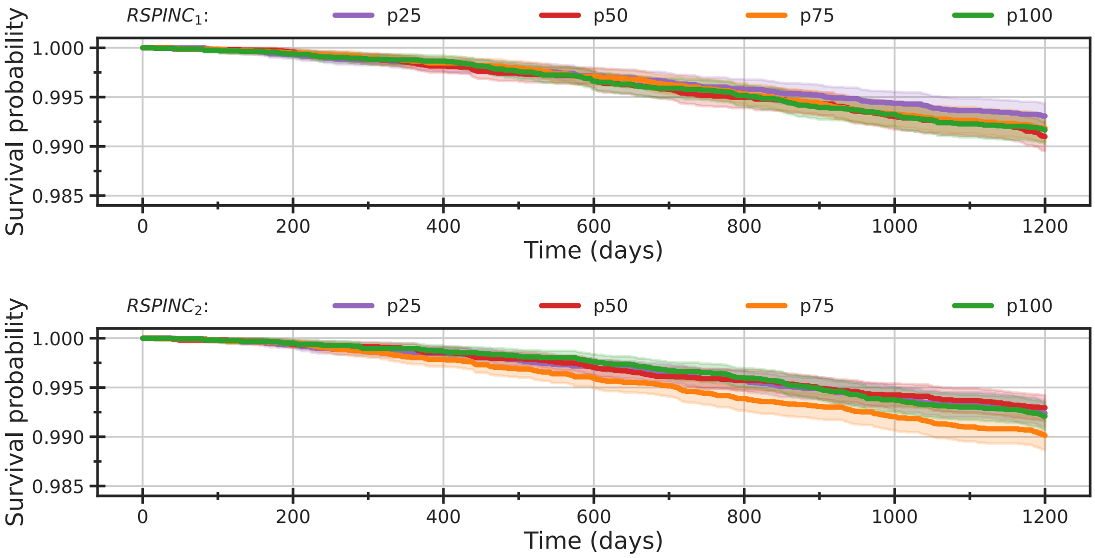


Supplementary Figure 8: **RSPINCs Kaplan-Meier curves**. Kaplan-Meier curves estimating the overall survival (OS) function for European individuals in the validation dataset ($n$=65,266). Individuals were stratified into quartiles using each RSPINC coordinate (e.g., "p25" denotes the bottom quartile) and OS curves were constructed using the standard Kaplan-Meier estimator with bootstrapped 95% confidence intervals. See Supplementary Table 9 for the corresponding hazard ratios per standard deviation.

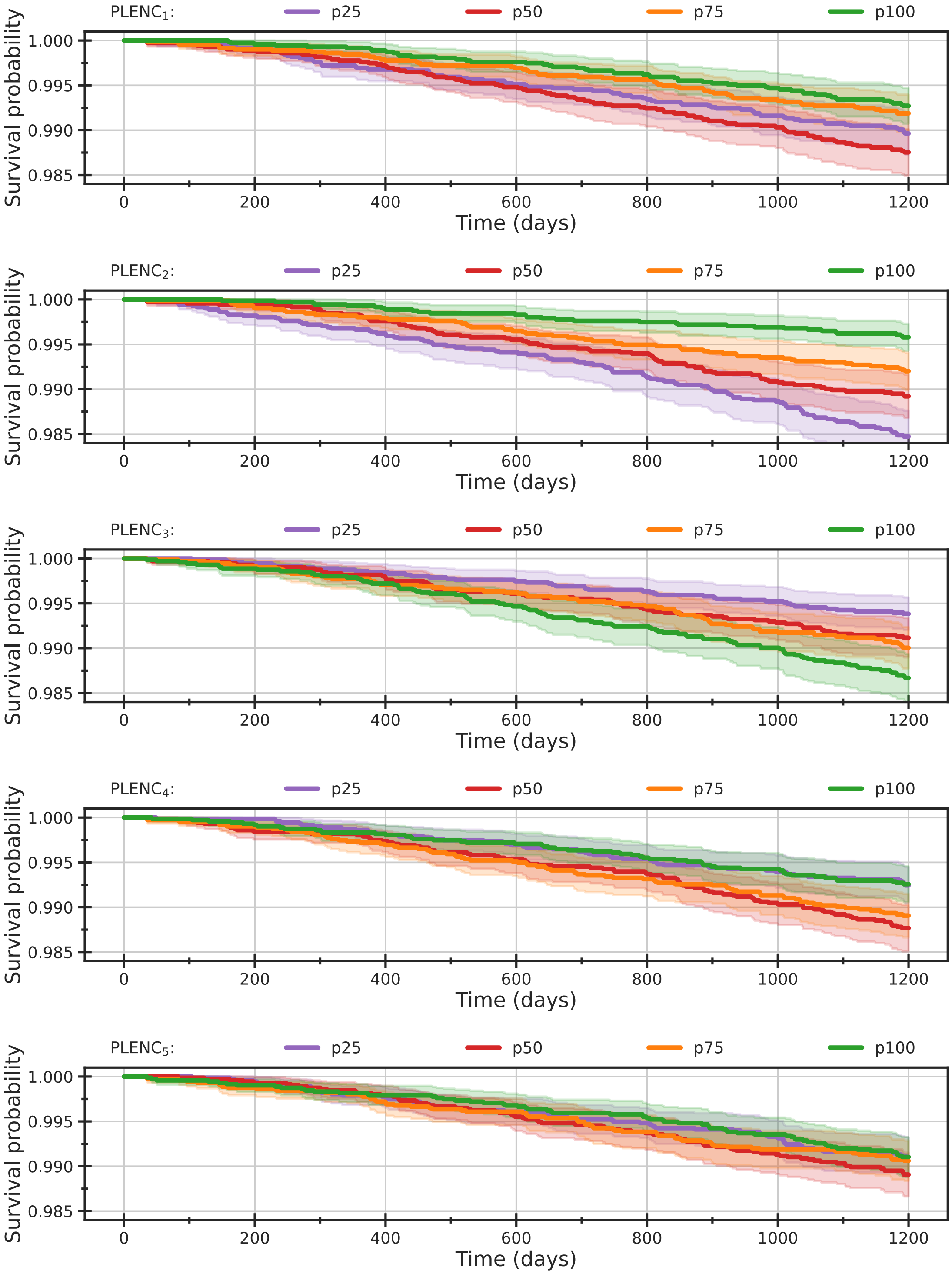


Supplementary Figure 9: **PLENCs Kaplan-Meier curves**. Kaplan-Meier curves estimating the overall survival (OS) function for European individuals in the validation dataset ($n$=28,545). Individuals were stratified into quartiles using each PLENC coordinate (e.g., “p25” denotes the bottom quartile) and OS curves were constructed using the standard Kaplan-Meier estimator with bootstrapped 95% confidence intervals. See Supplementary Table 9 for the corresponding hazard ratios per standard deviation.

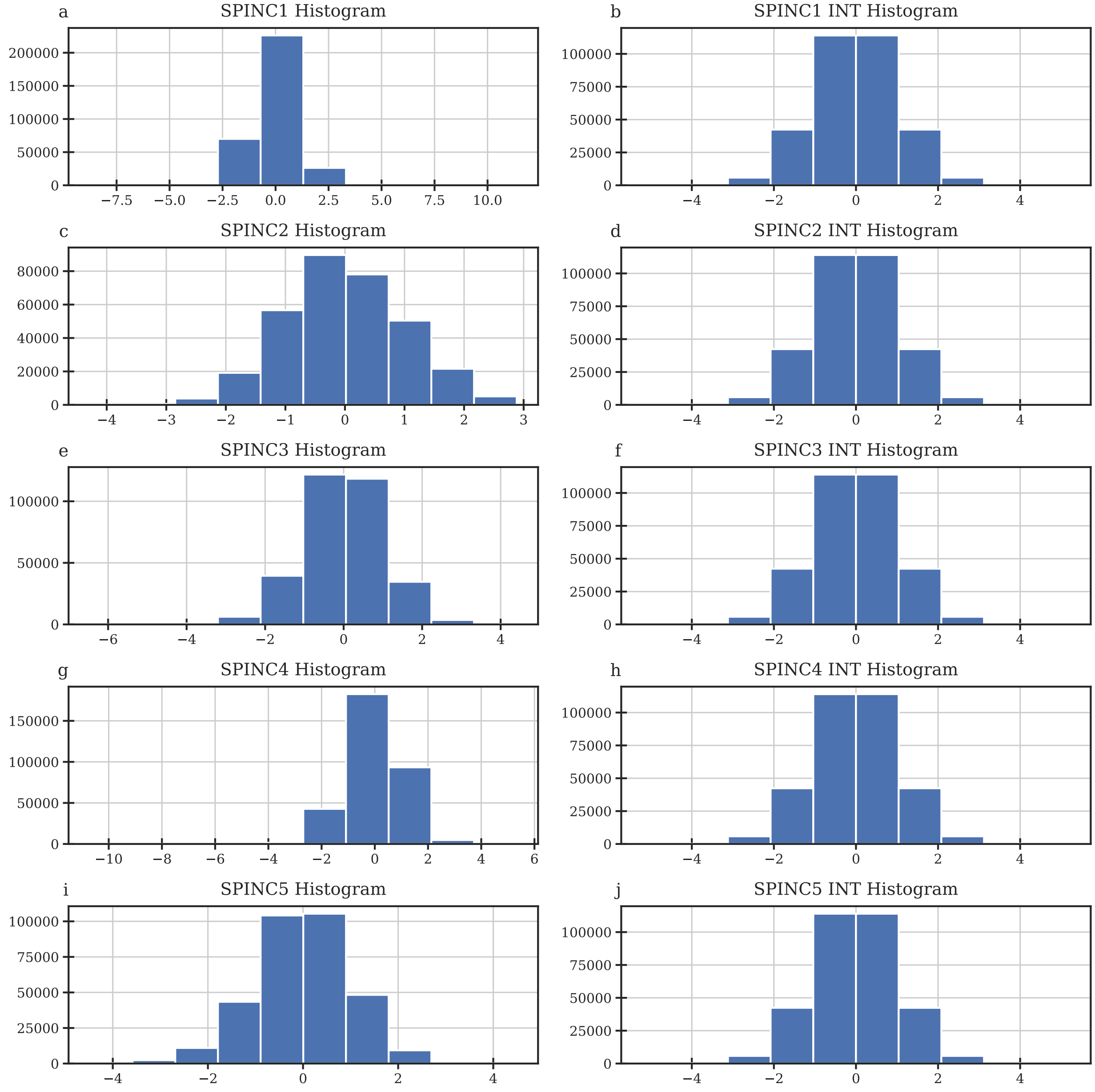


Supplementary Figure 10: **Distribution of SPINCs and inverse-normal transformed SPINCs coordinates of UK Biobank individuals.** Histogram of distribution for a) $\text{SPINC}_1$, b) $\text{SPINC}_1$ INT, c) $\text{SPINC}_2$, d) $\text{SPINC}_2$ INT, e) $\text{SPINC}_3$, f) $\text{SPINC}_3$ INT, g) $\text{SPINC}_4$, h) $\text{SPINC}_4$ INT, i) $\text{SPINC}_5$, and j) $\text{SPINC}_5$ INT.

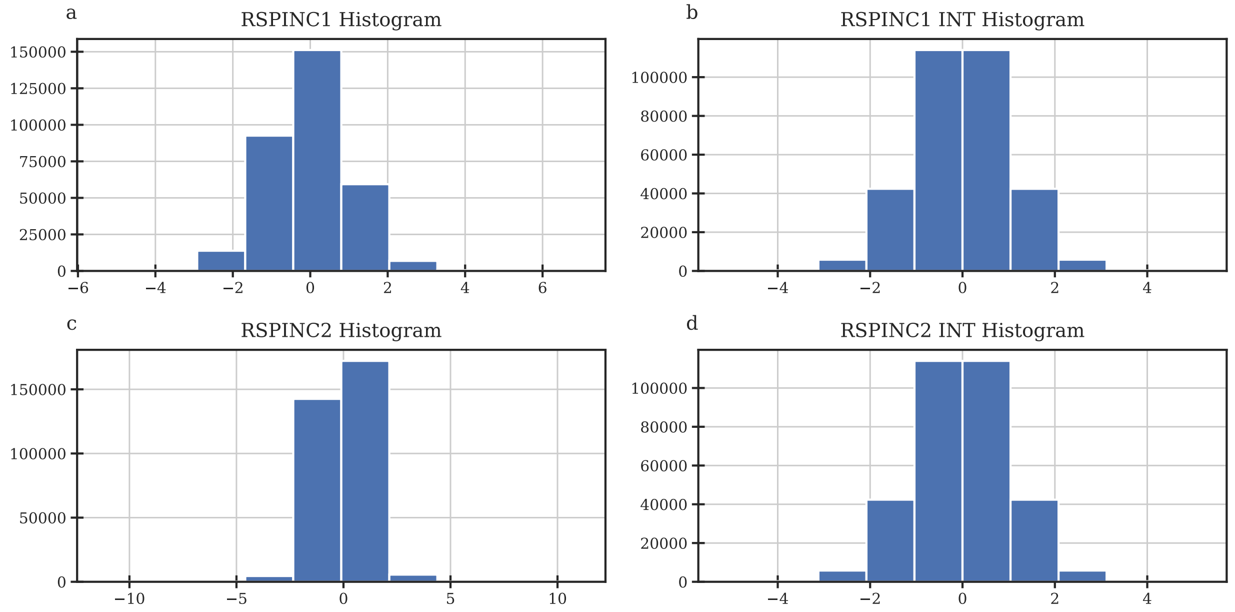


Supplementary Figure 11: **Distribution of RSPINCs and inverse-normal transformed RSPINCs coordinates of UK Biobank individuals.** Histogram of distribution for a) $\text{RSPINC}_1$, b) $\text{RSPINC}_1$ INT, c) $\text{RSPINC}_2$, and d) $\text{RSPINC}_2$ INT.

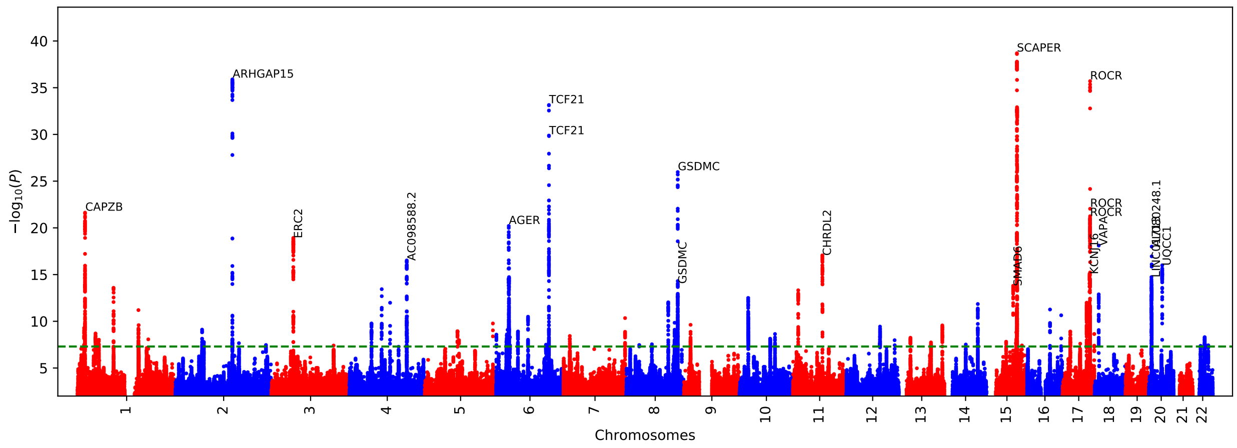


Supplementary Figure 12: **$\text{SPINC}_1$ GWAS Manhattan plot.**

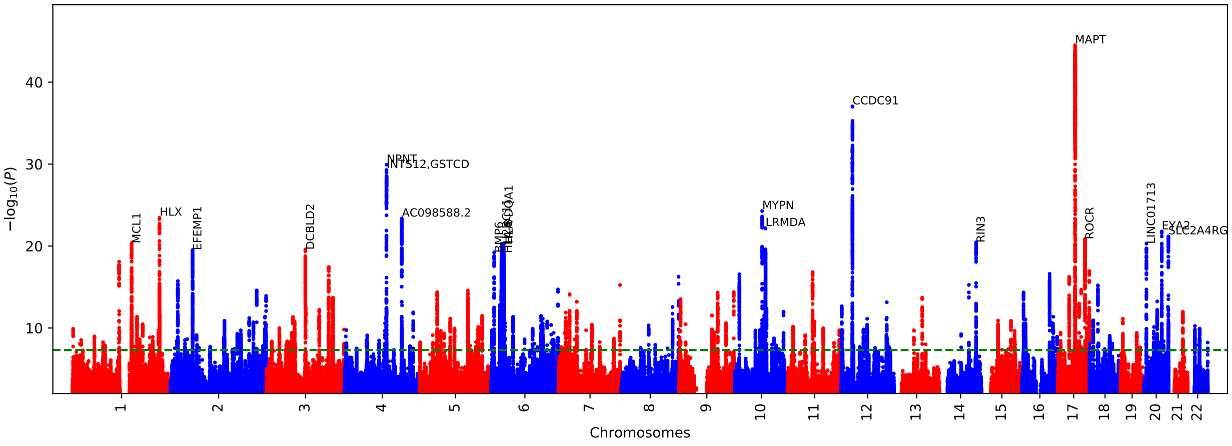


Supplementary Figure 13: **SPINC$_2$ GWAS Manhattan plot.**

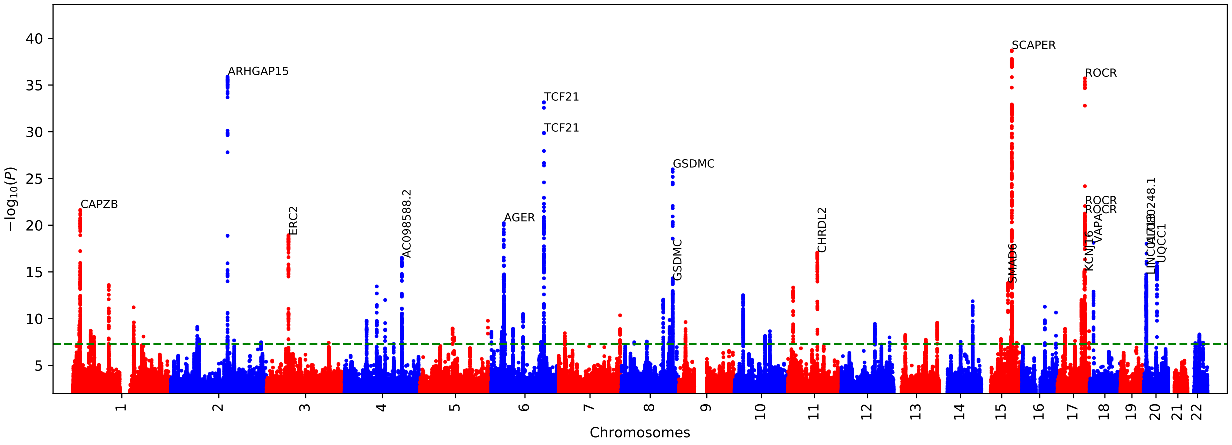


Supplementary Figure 14: **SPINC$_3$ GWAS Manhattan plot.**

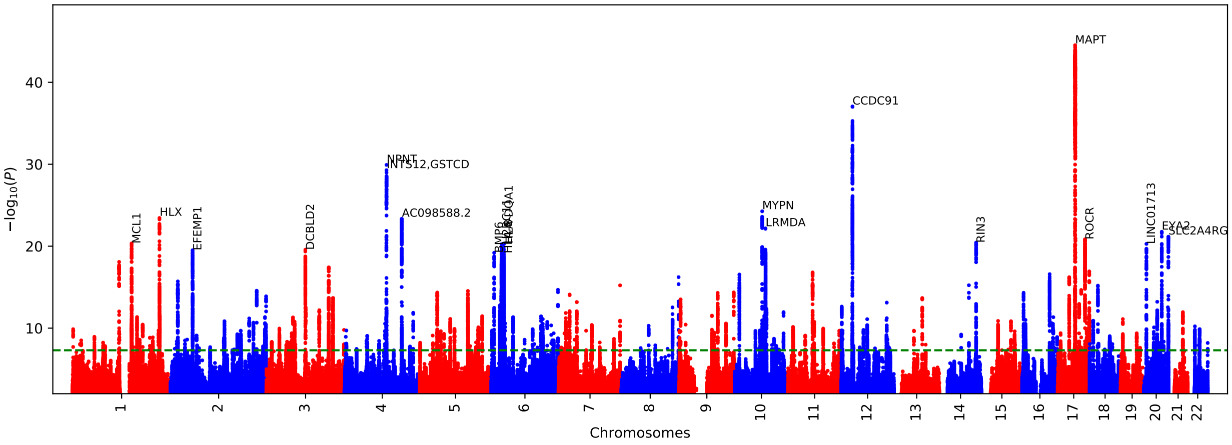


Supplementary Figure 15: **SPINC$_4$ GWAS Manhattan plot.**

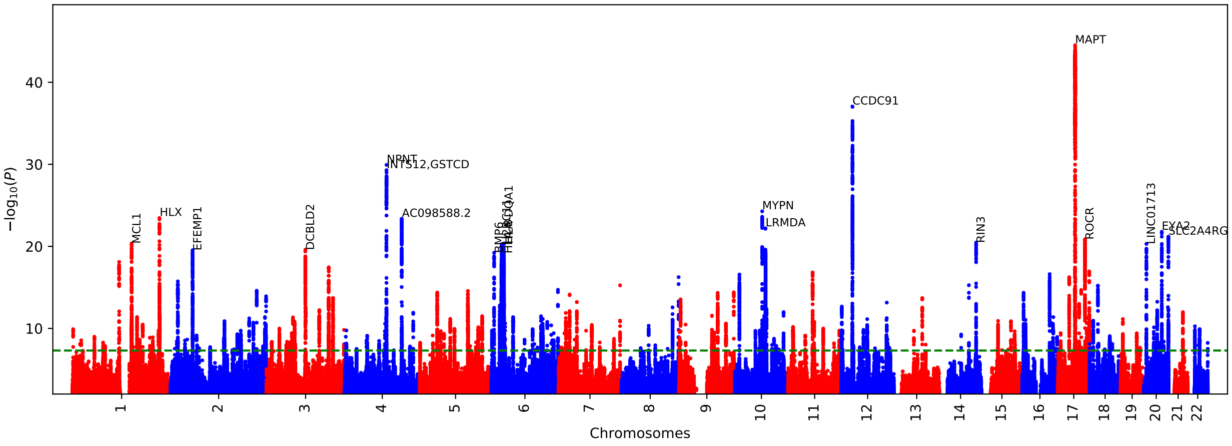


Supplementary Figure 16: **SPINC$_5$ GWAS Manhattan plot.**

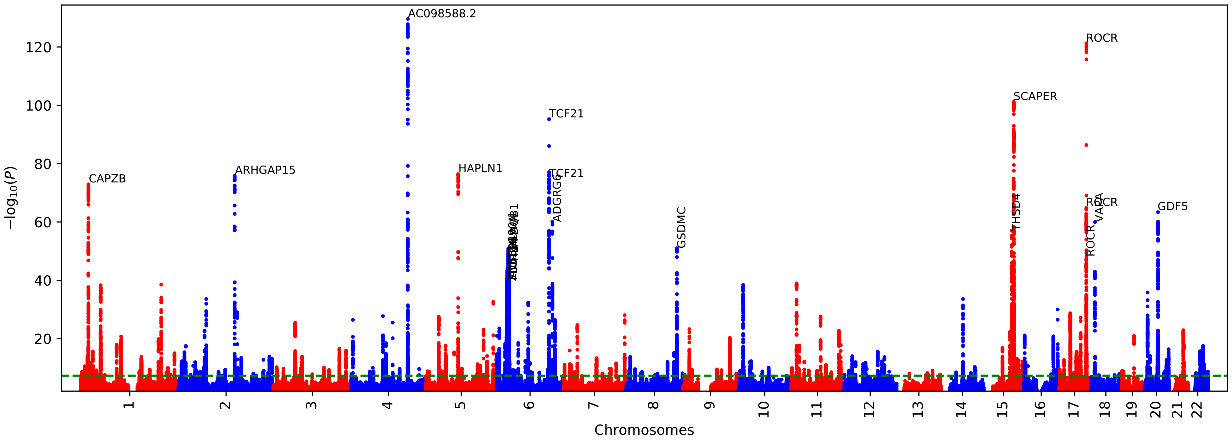


Supplementary Figure 17: **RSPINC$_1$ GWAS Manhattan plot.**

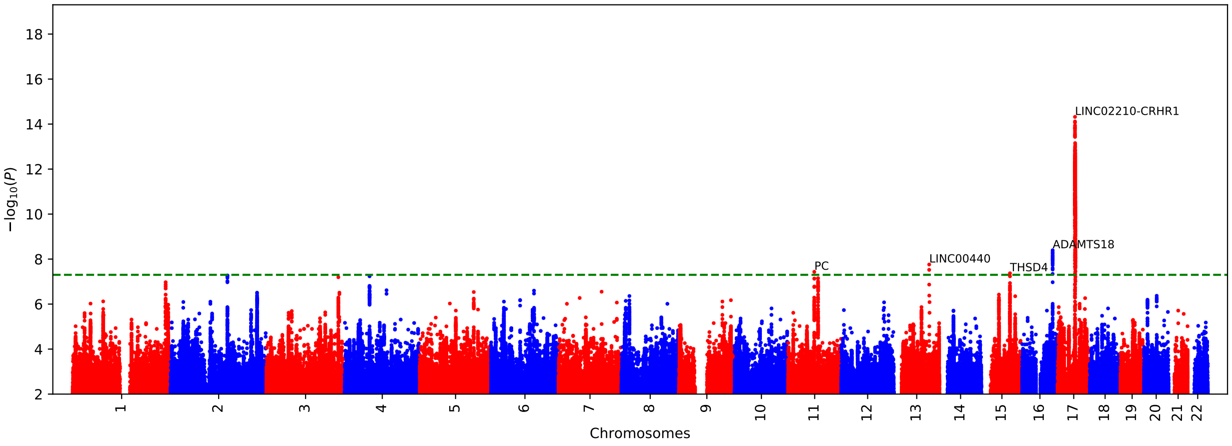


Supplementary Figure 18: **RSPINC$_2$ GWAS Manhattan plot.**

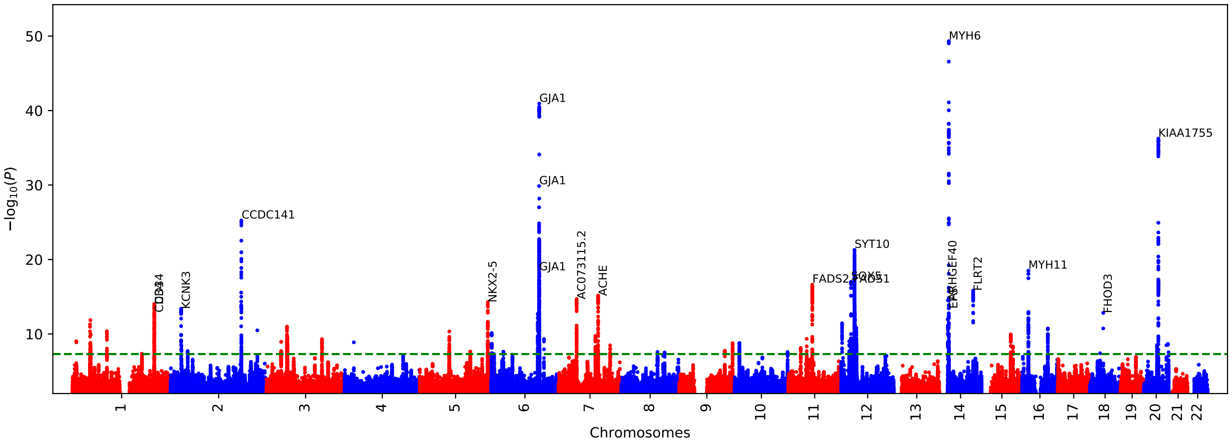


Supplementary Figure 19: **PLENC$_1$ GWAS Manhattan plot.**

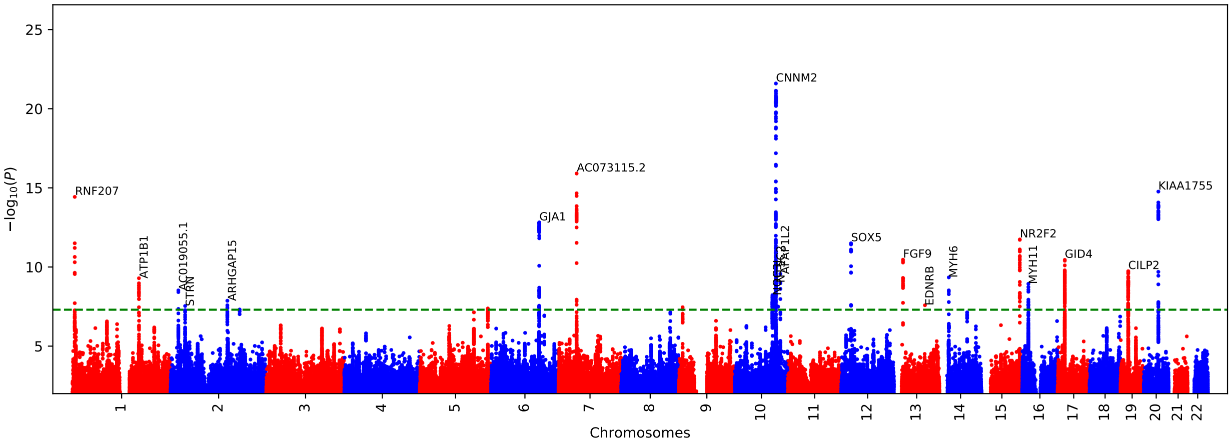


Supplementary Figure 20: **PLENC$_2$ GWAS Manhattan plot.**

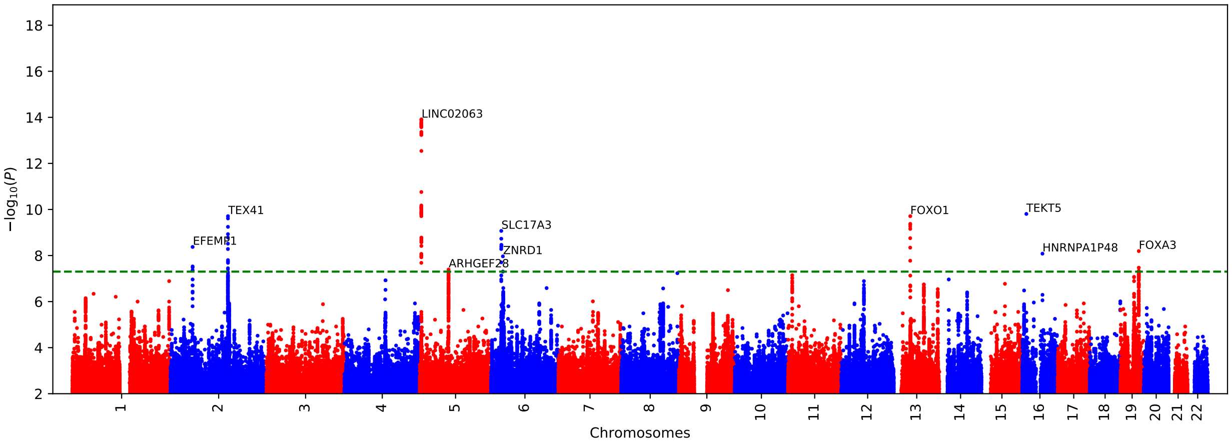


Supplementary Figure 21: **PLENC$_3$ GWAS Manhattan plot.**

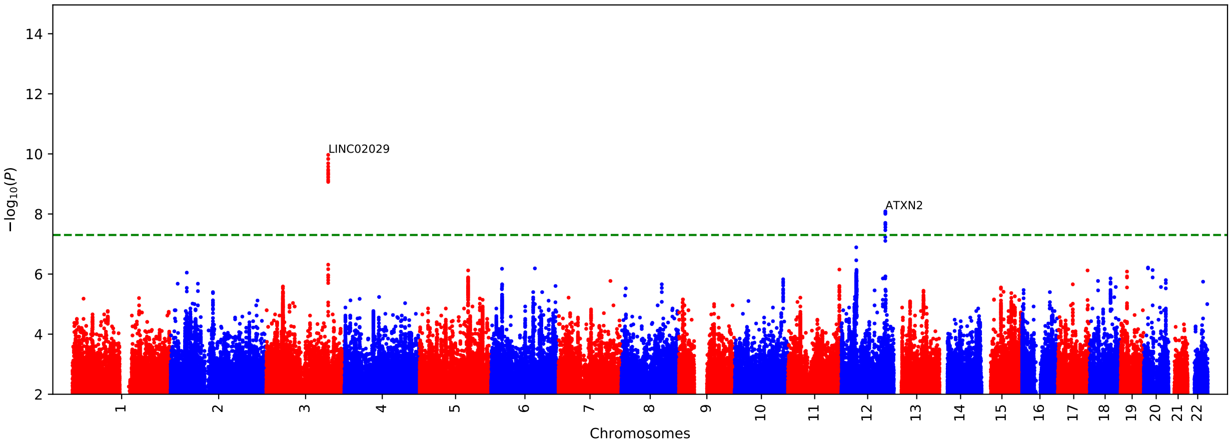


Supplementary Figure 22: **$PLENC_4$ GWAS Manhattan plot.**

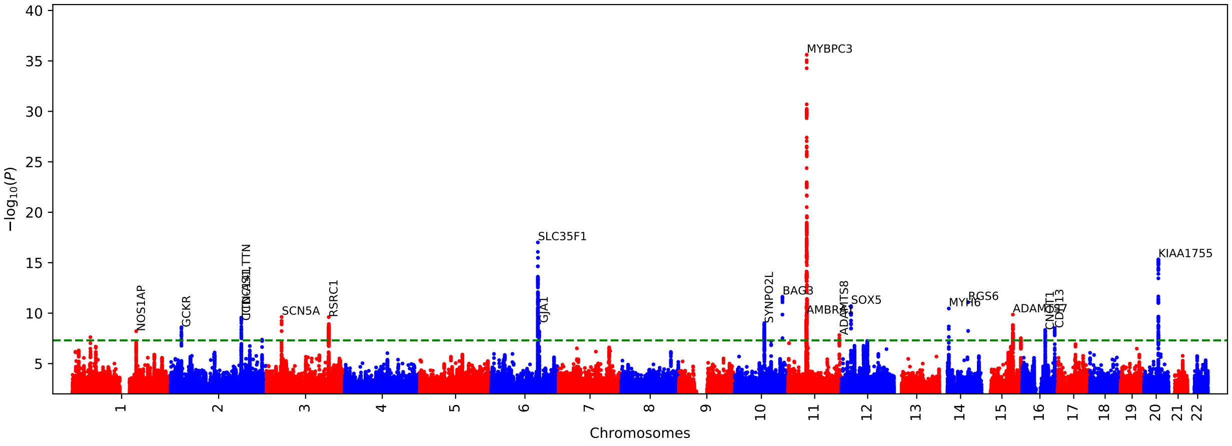


Supplementary Figure 23: **$PLENC_5$ GWAS Manhattan plot.**

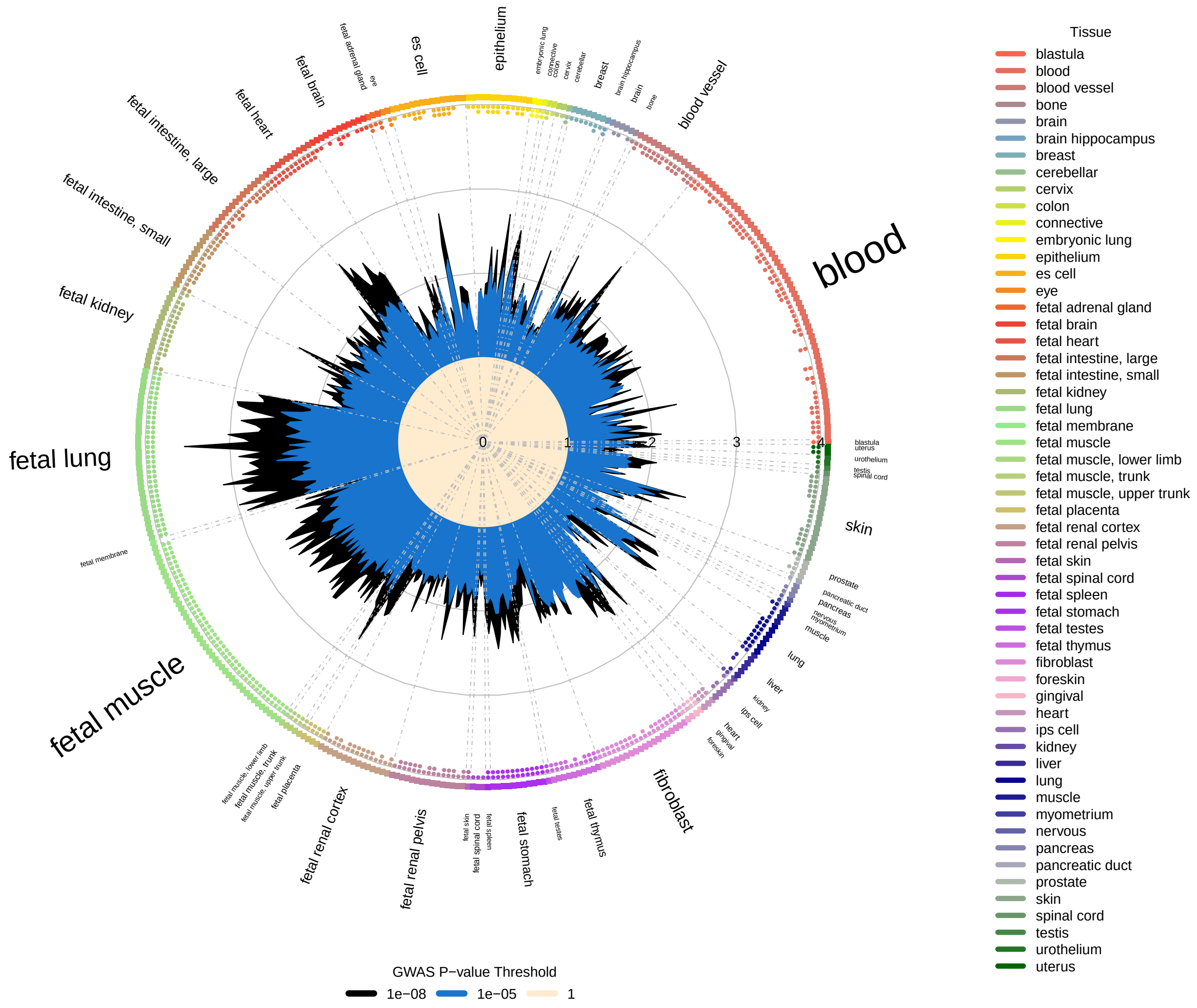


Supplementary Figure 24: **Enrichment overlap of SPINC$_1$ GWAS with DNase I hotspots computed using GARFIELD.** Radial plot illustrates the enrichment (OR) in each cell type for different GWAS p-value thresholds ($P < 10^{-8}$ and $10^{-5}$). In addition, the small dots on the outer side of the plot indicates enrichment significant level computed by GARFIELD for different significant level of $10^{-5}$, $10^{-6}$, $10^{-7}$, and $10^{-8}$ in direction of outside to insider of plot.

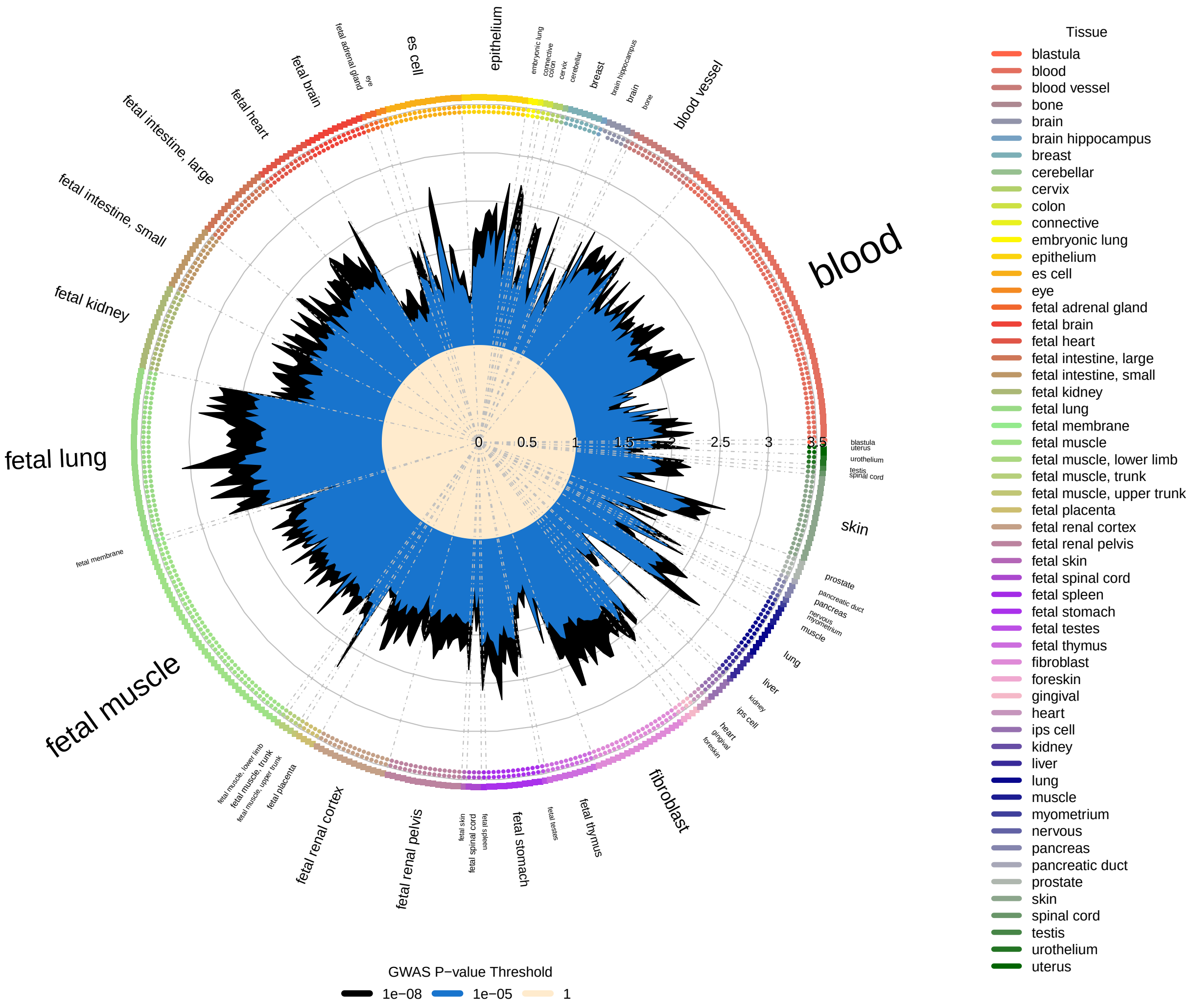


Supplementary Figure 25: **Enrichment overlap of $SPINC_2$ GWAS with DNase I hotspots computed using GARFIELD.** Radial plot illustrates the enrichment (OR) in each cell type for different GWAS p-value thresholds ($P < 10^{-8}$ and $10^{-5}$). In addition, the small dots on the outer side of the plot indicates enrichment significant level computed by GARFIELD for different significant level of $10^{-5}$, $10^{-6}$, $10^{-7}$, and $10^{-8}$ in direction of outside to insider of plot.

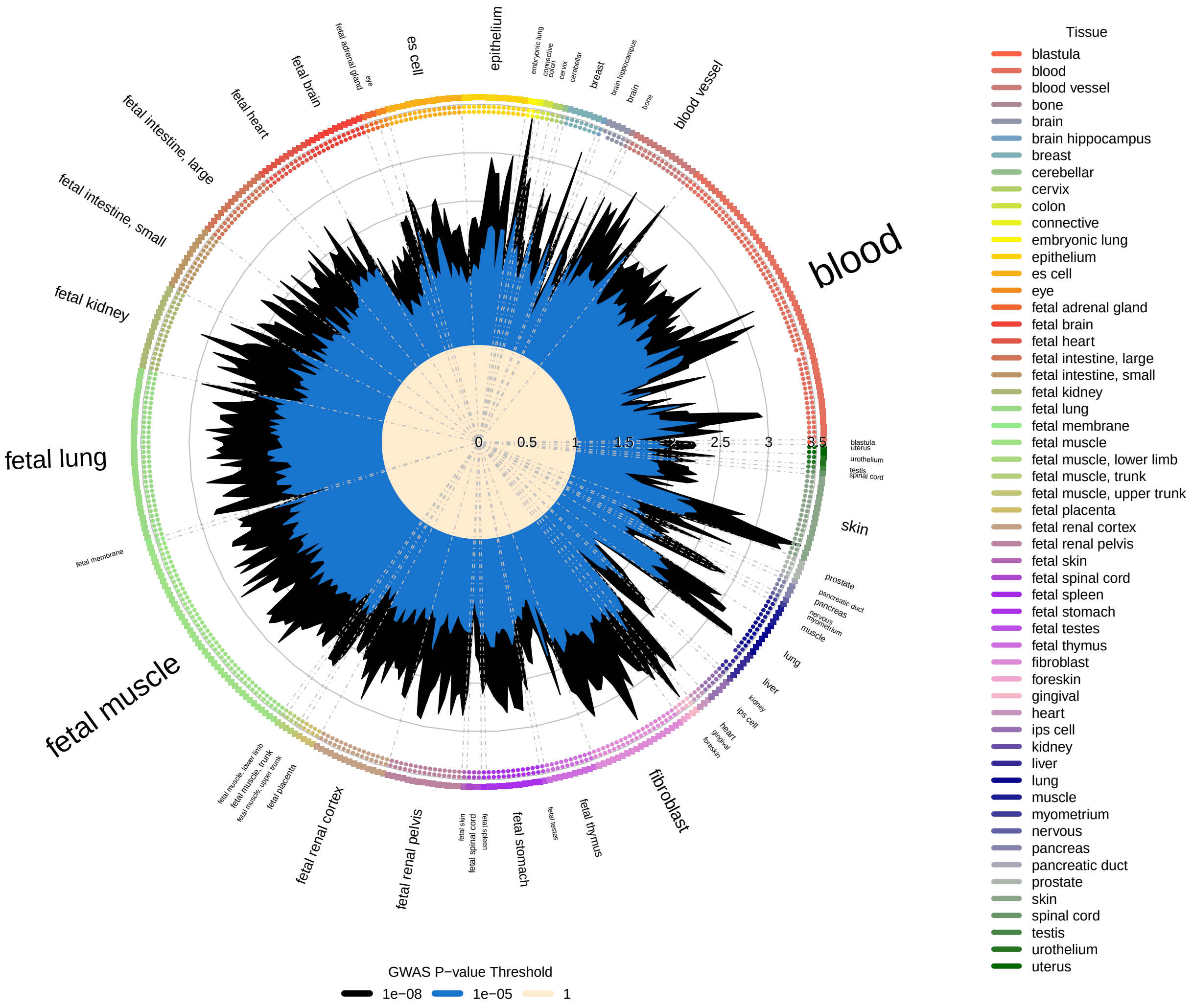


Supplementary Figure 26: **Enrichment overlap of SPINC$_3$ GWAS with DNase I hotspots computed using GARFIELD.** Radial plot illustrates the enrichment (OR) in each cell type for different GWAS p-value thresholds ($P < 10^{-8}$ and $10^{-5}$). In addition, the small dots on the outer side of the plot indicates enrichment significant level computed by GARFIELD for different significant level of $10^{-5}$, $10^{-6}$, $10^{-7}$, and $10^{-8}$ in direction of outside to insider of plot.

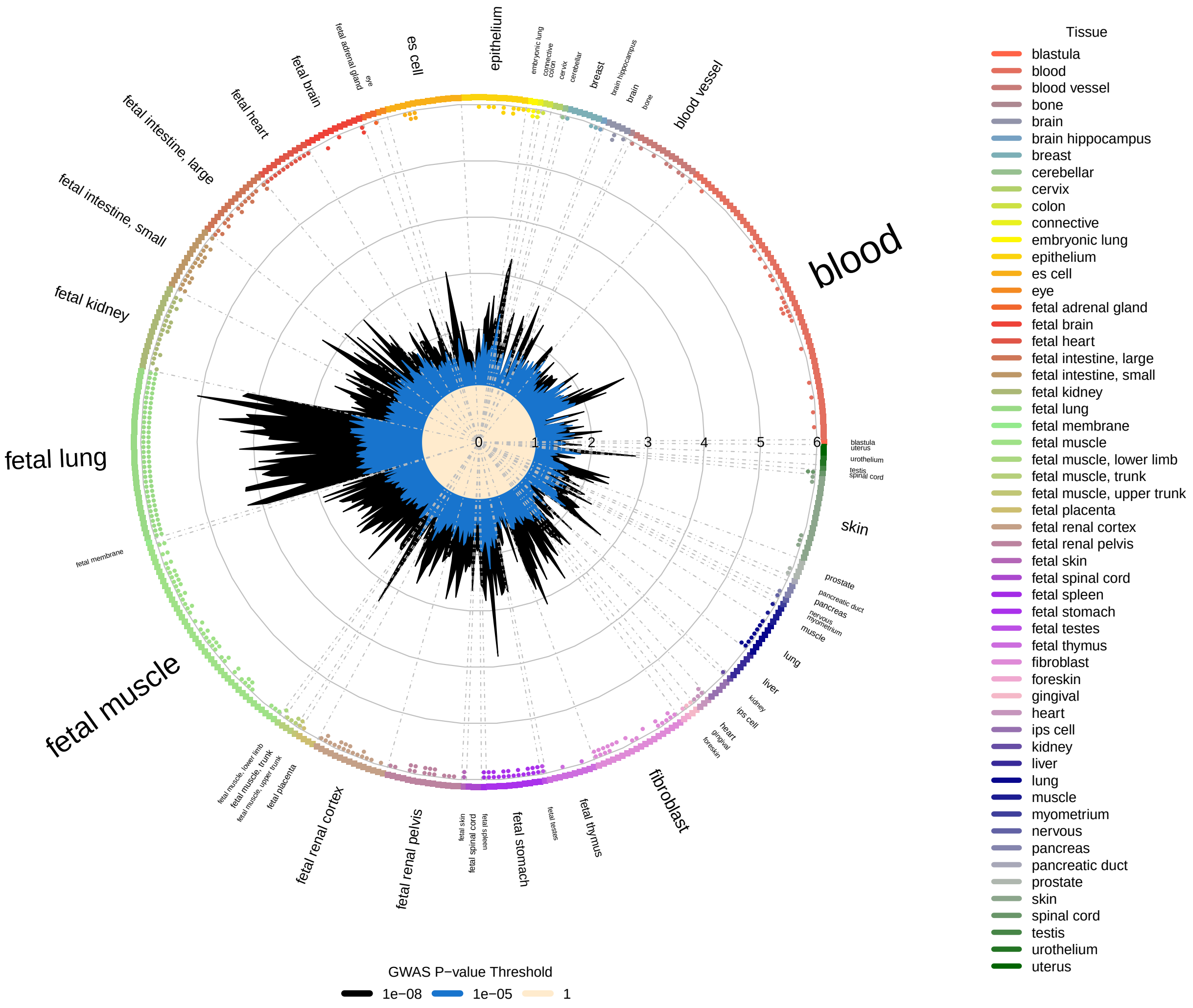


Supplementary Figure 27: **Enrichment overlap of SPINC$_4$ GWAS with DNase I hotspots computed using GARFIELD.** Radial plot illustrates the enrichment (OR) in each cell type for different GWAS p-value thresholds ($P < 10^{-8}$ and $10^{-5}$). In addition, the small dots on the outer side of the plot indicates enrichment significant level computed by GARFIELD for different significant level of $10^{-5}$, $10^{-6}$, $10^{-7}$, and $10^{-8}$ in direction of outside to insider of plot.

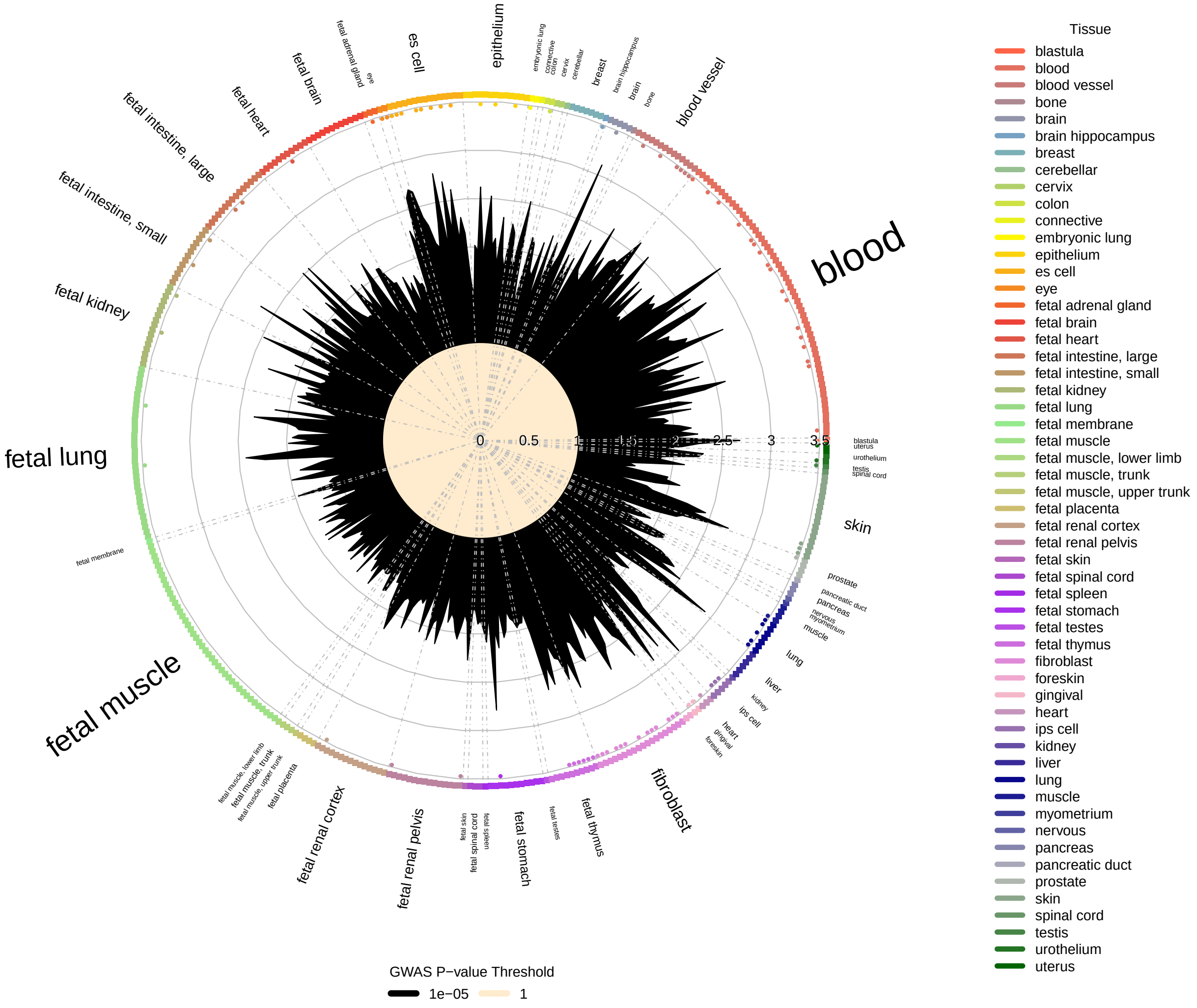


Supplementary Figure 28: **Enrichment overlap of SPINC$_5$ GWAS with DNase I hotspots computed using GARFIELD.** Radial plot illustrates the enrichment (OR) in each cell type for different GWAS p-value thresholds ($P < 10^{-8}$ and $10^{-5}$). In addition, the small dots on the outer side of the plot indicates enrichment significant level computed by GARFIELD for different significant level of $10^{-5}$, $10^{-6}$, $10^{-7}$, and $10^{-8}$ in direction of outside to insider of plot.

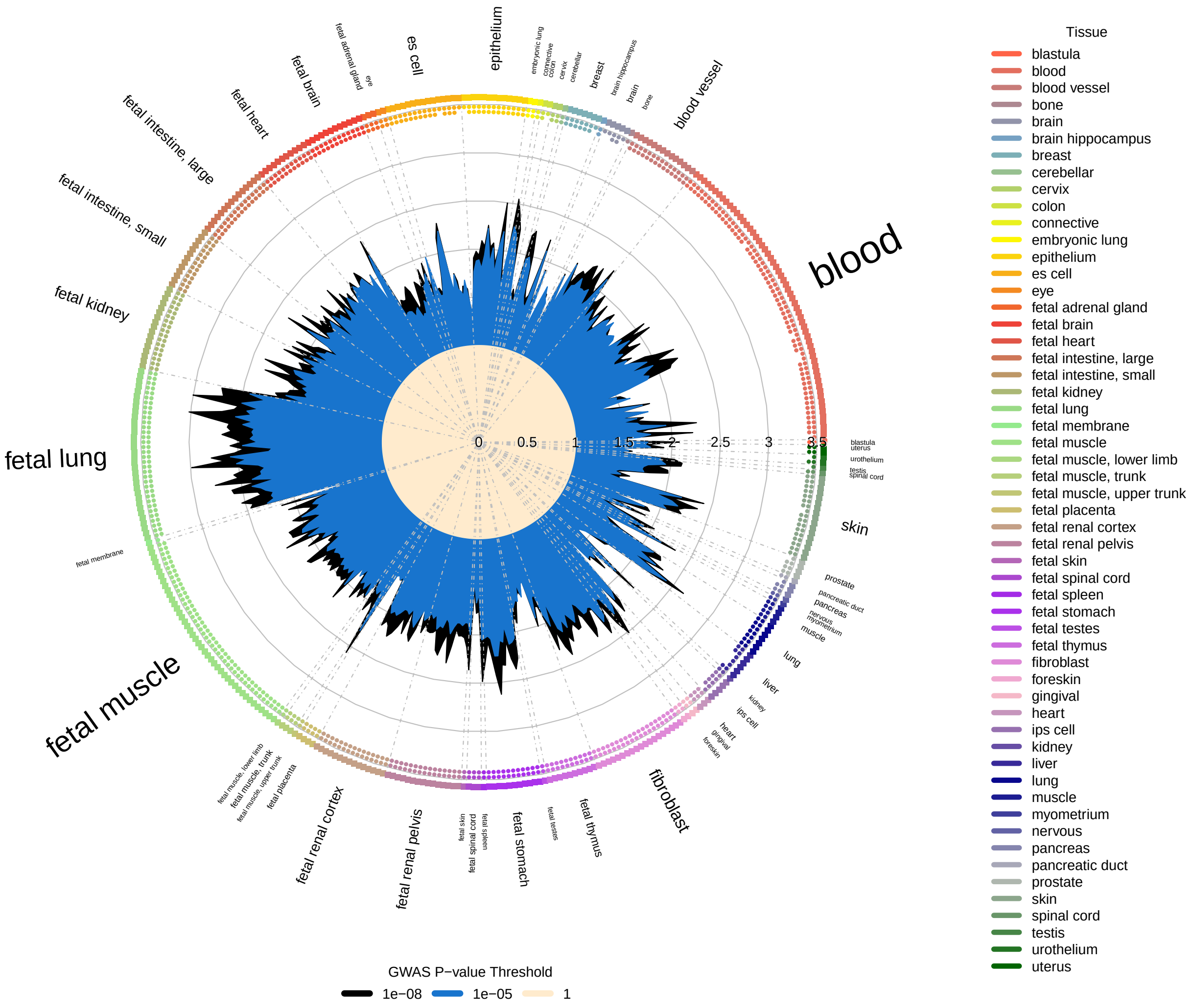


Supplementary Figure 29: **Enrichment overlap of RSPINC$_1$ GWAS with DNase I hotspots computed using GARFIELD.** Radial plot illustrates the enrichment (OR) in each cell type for different GWAS p-value thresholds ($P < 10^{-8}$ and $10^{-5}$). In addition, the small dots on the outer side of the plot indicates enrichment significant level computed by GARFIELD for different significant level of $10^{-5}$, $10^{-6}$, $10^{-7}$, and $10^{-8}$ in direction of outside to insider of plot.

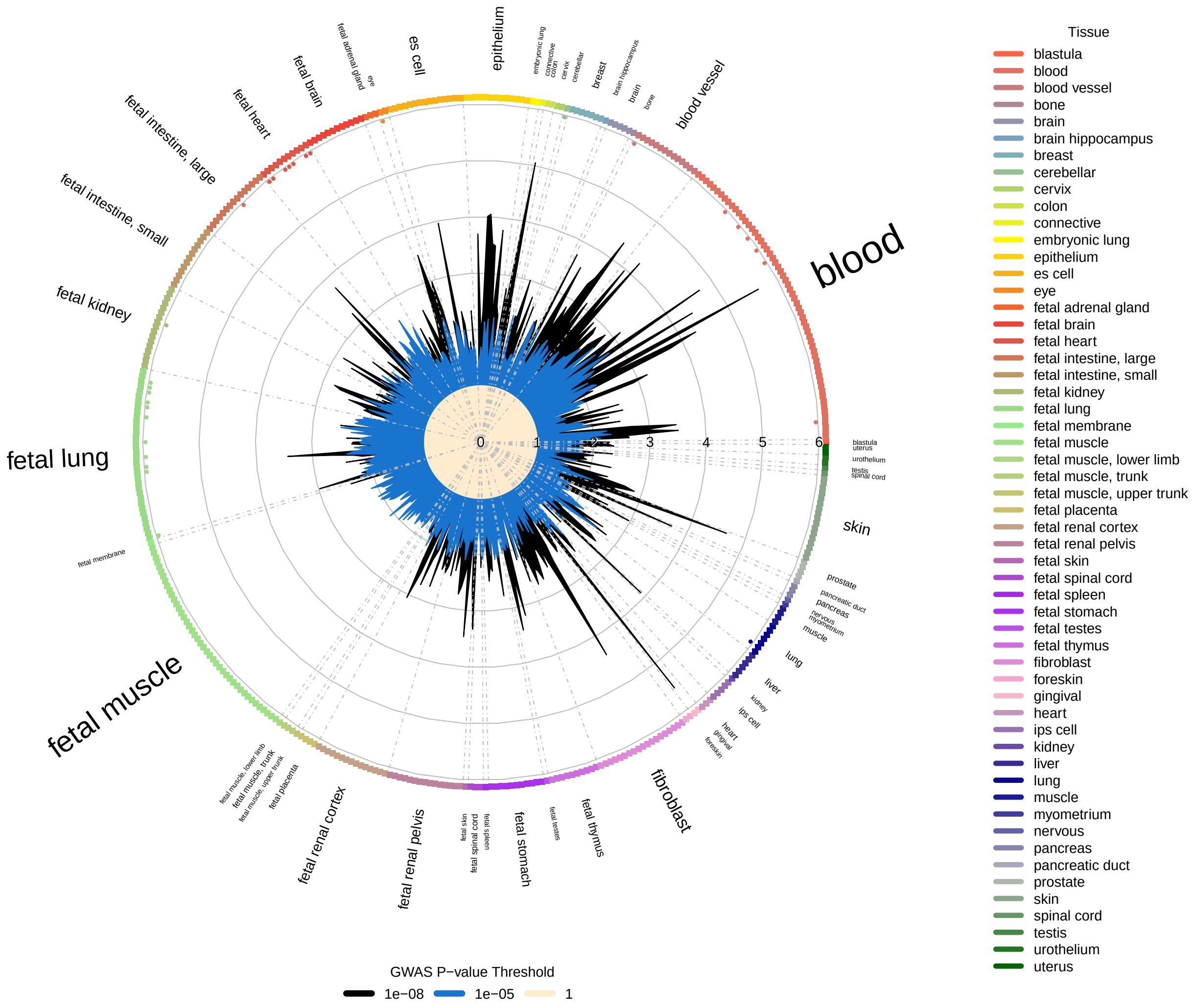


Supplementary Figure 30: **Enrichment overlap of RSPINC$_2$ GWAS with DNase I hotspots computed using GARFIELD.** Radial plot illustrates the enrichment (OR) in each cell type for different GWAS p-value thresholds ($P < 10^{-8}$ and $10^{-5}$). In addition, the small dots on the outer side of the plot indicates enrichment significant level computed by GARFIELD for different significant level of $10^{-5}$, $10^{-6}$, $10^{-7}$, and $10^{-8}$ in direction of outside to insider of plot.

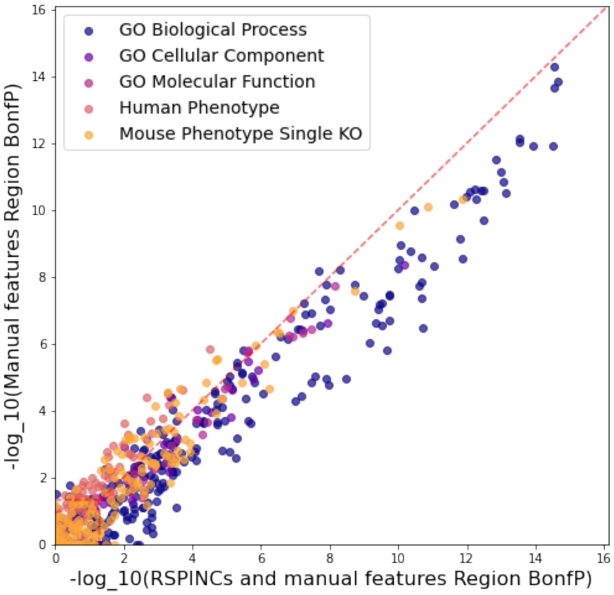


Supplementary Figure 31: **GREAT region-based enrichments for traditional measurements and RSPINCs.** The set of loci discovered through the union of traditional measurements and RSPINCs produces enrichments with lower P-values than the loci from traditional measurements alone.

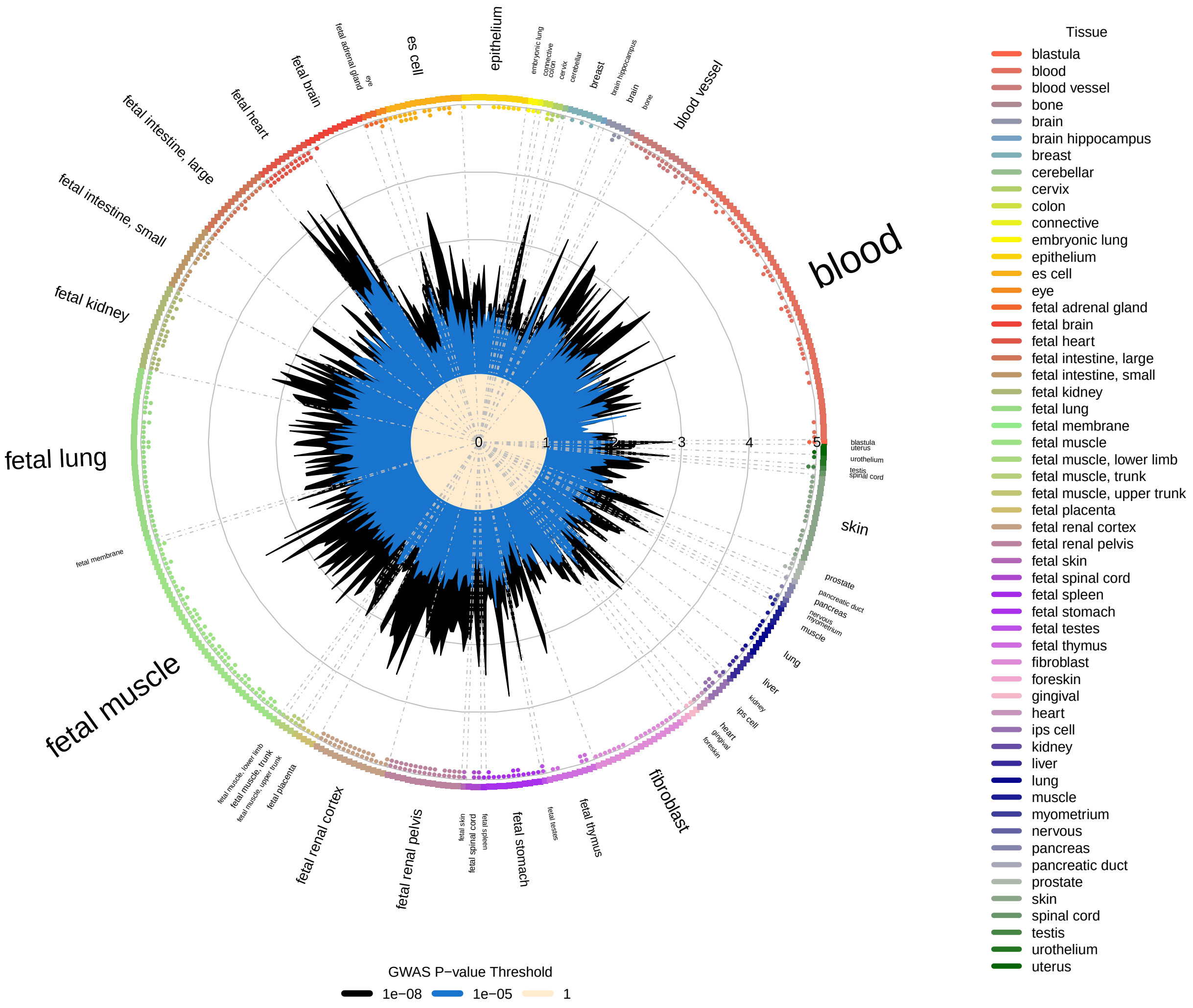


Supplementary Figure 32: **Enrichment overlap of PLENC$_1$ GWAS with DNase I hotspots computed using GARFIELD.** Radial plot illustrates the enrichment (OR) in each cell type for different GWAS p-value thresholds ($P < 10^{-8}$ and $10^{-5}$). In addition, the small dots on the outer side of the plot indicates enrichment significant level computed by GARFIELD for different significant level of $10^{-5}$, $10^{-6}$, $10^{-7}$, and $10^{-8}$ in direction of outside to insider of plot.

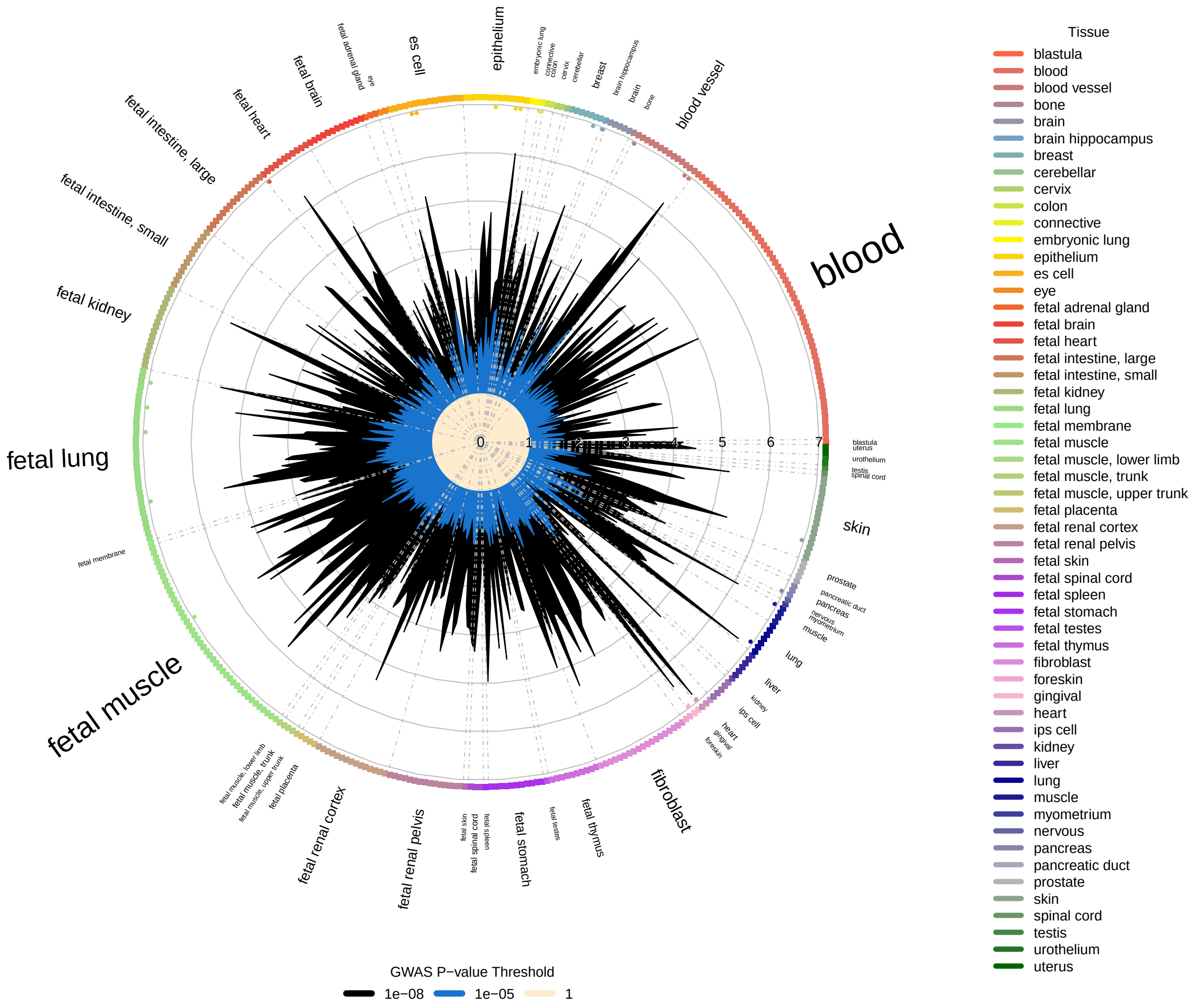


Supplementary Figure 33: **Enrichment overlap of PLENC$_2$ GWAS with DNase I hotspots computed using GARFIELD.** Radial plot illustrates the enrichment (OR) in each cell type for different GWAS p-value thresholds ($P < 10^{-8}$ and $10^{-5}$). In addition, the small dots on the outer side of the plot indicates enrichment significant level computed by GARFIELD for different significant level of $10^{-5}$, $10^{-6}$, $10^{-7}$, and $10^{-8}$ in direction of outside to insider of plot.

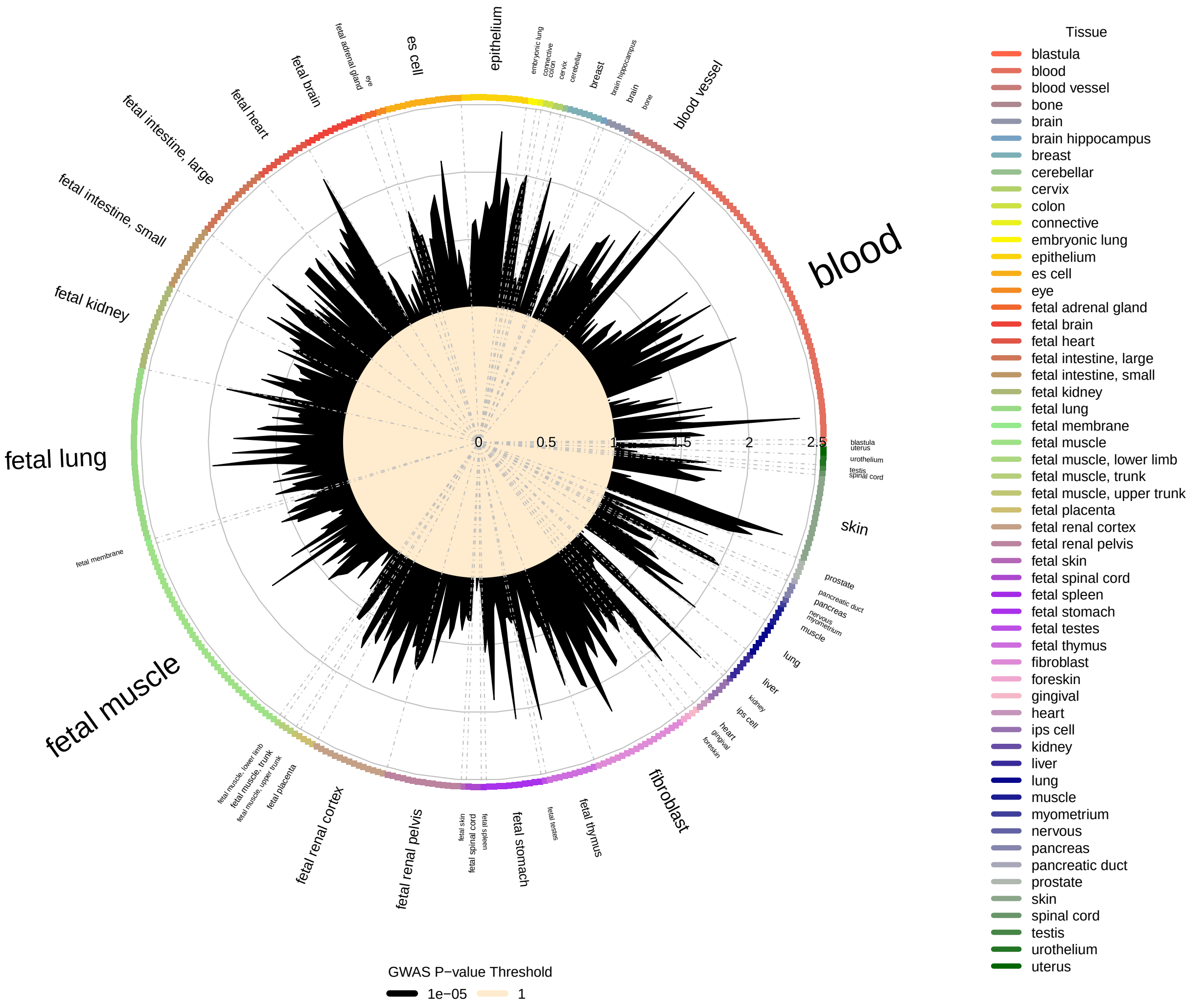


Supplementary Figure 34: **Enrichment overlap of PLENC$_3$ GWAS with DNase I hotspots computed using GARFIELD.** Radial plot illustrates the enrichment (OR) in each cell type for different GWAS p-value thresholds ($P < 10^{-8}$ and $10^{-5}$). In addition, the small dots on the outer side of the plot indicates enrichment significant level computed by GARFIELD for different significant level of $10^{-5}$, $10^{-6}$, $10^{-7}$, and $10^{-8}$ in direction of outside to insider of plot.

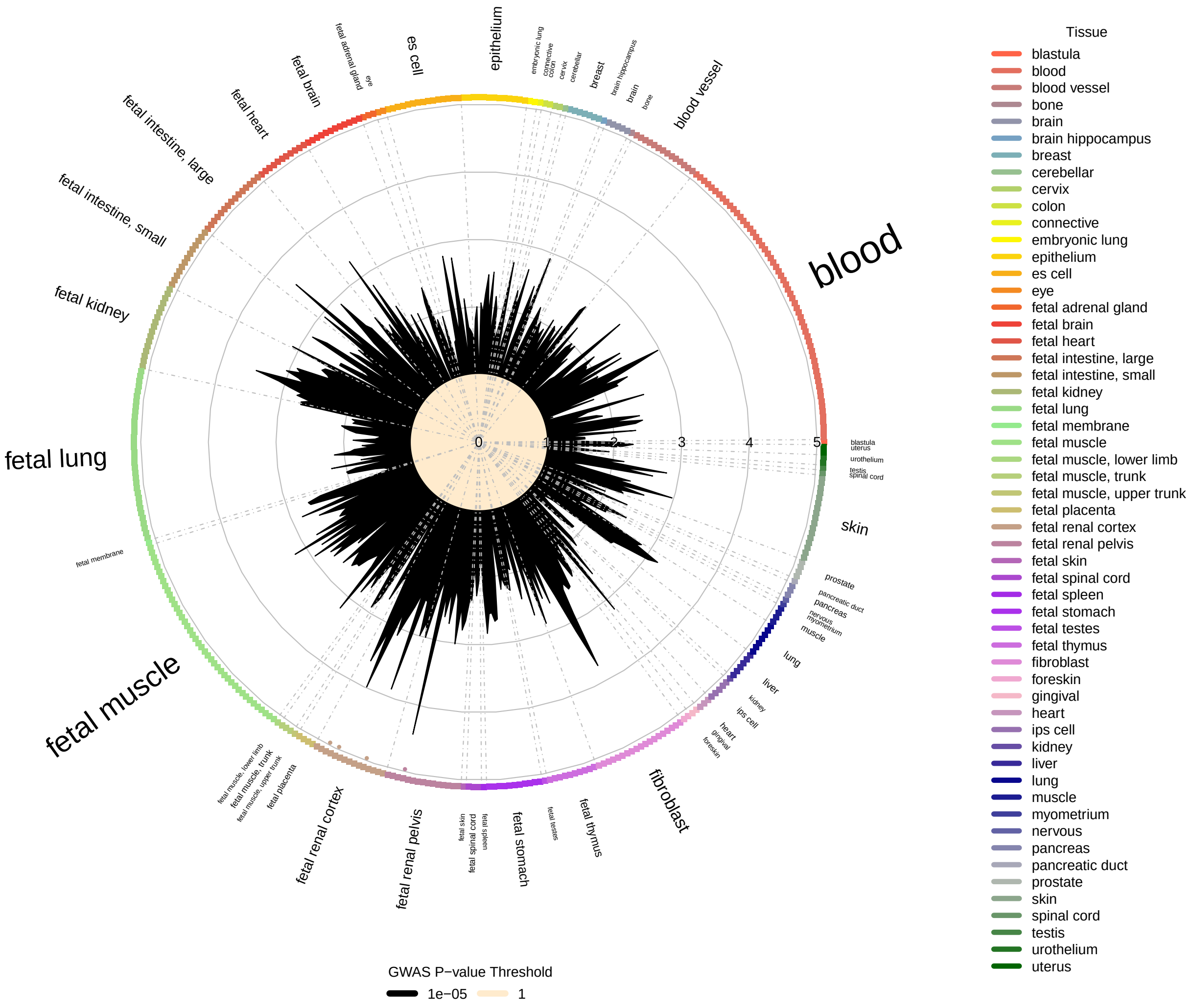


Supplementary Figure 35: **Enrichment overlap of PLENC$_4$ GWAS with DNase I hotspots computed using GARFIELD.** Radial plot illustrates the enrichment (OR) in each cell type for different GWAS p-value thresholds ($P < 10^{-8}$ and $10^{-5}$). In addition, the small dots on the outer side of the plot indicates enrichment significant level computed by GARFIELD for different significant level of $10^{-5}$, $10^{-6}$, $10^{-7}$, and $10^{-8}$ in direction of outside to insider of plot.

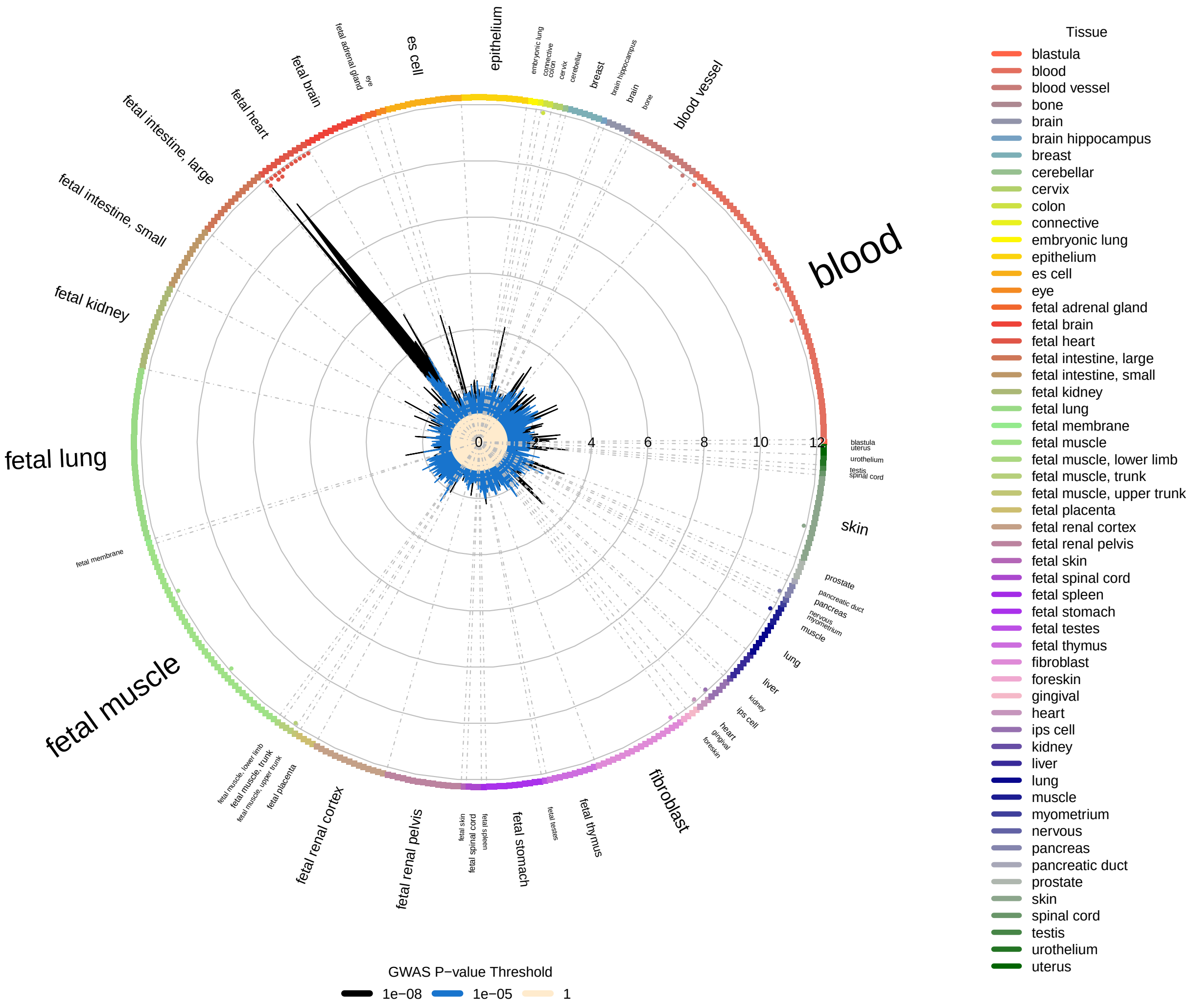


Supplementary Figure 36: **Enrichment overlap of PLENC$_5$ GWAS with DNase I hotspots computed using GARFIELD.** Radial plot illustrates the enrichment (OR) in each cell type for different GWAS p-value thresholds ($P < 10^{-8}$ and $10^{-5}$). In addition, the small dots on the outer side of the plot indicates enrichment significant level computed by GARFIELD for different significant level of $10^{-5}$, $10^{-6}$, $10^{-7}$, and $10^{-8}$ in direction of outside to insider of plot.

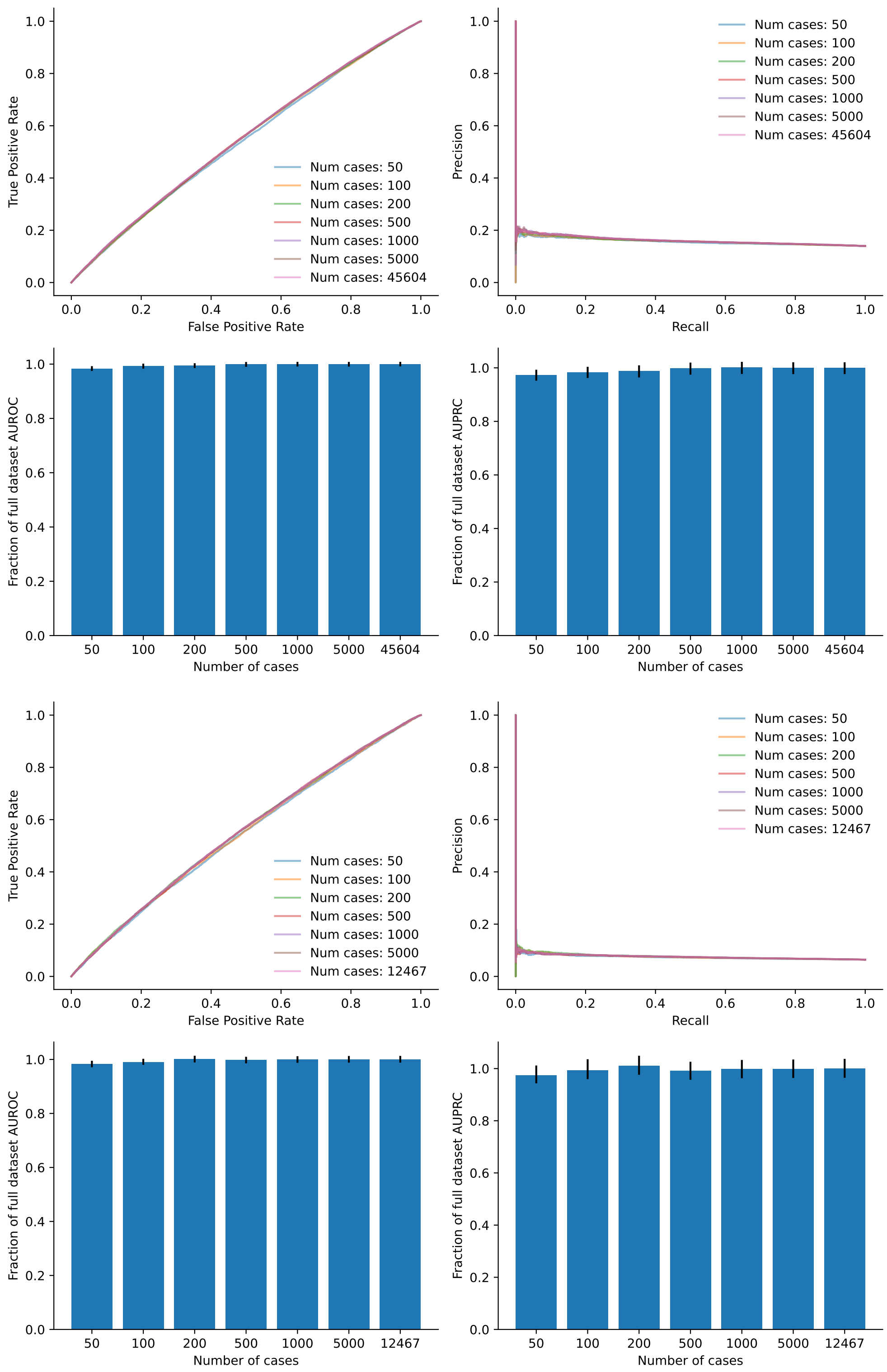


Supplementary Figure 37: **PRS performance under labeled training data ablation**. Datasets with balanced numbers of cases and controls were used to train PRS in Europeans for asthma (top four panels) and COPD (bottom four panels). Bar plots indicate the relative performance for AUROC (left) and AUPRC (right) using the largest number of cases as the reference performance level.

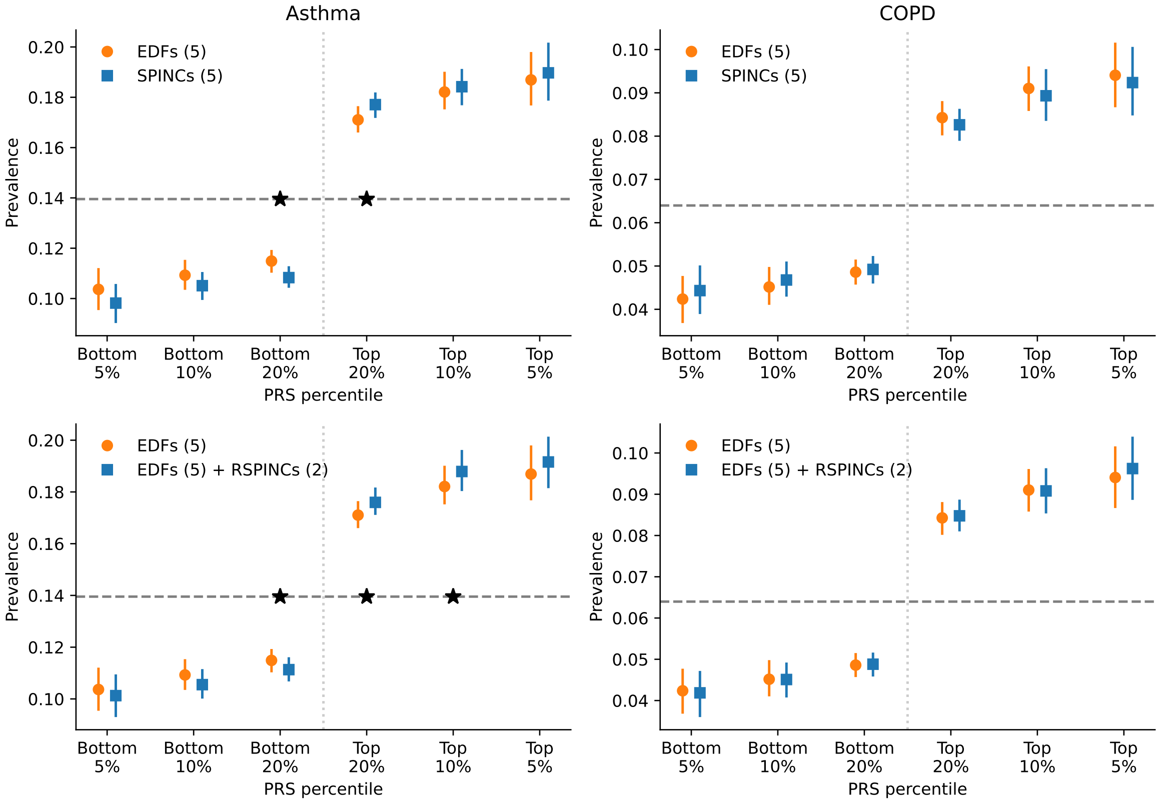


Supplementary Figure 38: **PRS using SPINCs and RSPINCs in UK Biobank with inverse normal transformation.** Combined PRS for asthma and COPD using three sets of intermediate PRS, five EDFs, five SPINCs, and five EDFs + two RSPINCs, after applying inverse-normal transformation on all. Each set of PRS is combined by a linear model trained using the target phenotype labels and the prevalence of the phenotypes in the top and bottom 5%, 10%, and 20% PRS individuals is evaluated in a separate evaluation set. Vertical line segments indicate 95% confidence interval generated by bootstrapping (300 samples). The horizontal dashed line shows the total prevalence. Star (*) signs indicate a statistically significant difference between the two methods using *paired* bootstrapping (300 samples) with $p < 0.05$. Lower is better for the bottom percentiles; higher is better for the top percentiles.

### Supplementary Tables

| Method | Optimizer | Learning rate | Batch size |
|---|---|---|---|
| SPINCs | Adam | 1E-4 | 16 |
| RSPINCs | Adam | 1E-4 | 16 |
| PLENCs | Adam | 1E-4 | 16 |

Supplementary Table 1: **Overview of the final hyperparameters used for each method.** Hyperparameter search was run over the learning rates of {1E-5, 1E-4, 1E-3} and the batch sizes of {16, 32, 64}. See also “SPINCs model architecture”, “RSPINCs model architecture”, and “PLENCs model architecture” in Supplementary Notes.

| Latent Dimension | SPINCs | PCA | RSPINCs |
|---|---|---|---|
| 1 | 0.063391 | 0.102179 | 0.038280 |
| 2 | 0.015881 | 0.051901 | 0.027203 |
| 3 | 0.008463 | 0.027312 | 0.019421 |
| 4 | 0.006265 | 0.017987 | 0.014800 |
| 5 | 0.005251 | 0.012641 | 0.012816 |
| 6 | 0.005263 | 0.009538 | 0.011725 |

Supplementary Table 2: **Reconstruction error of Spirograms based on MSE.**

| Latent Dimension | PLENCs | PCA |
|---|---|---|
| 1 | 0.161438 | 0.304929 |
| 2 | 0.085320 | 0.167717 |
| 3 | 0.055498 | 0.091606 |
| 4 | 0.044633 | 0.053183 |
| 5 | 0.039191 | 0.037153 |

Supplementary Table 3: **Reconstruction error of PPGs based on MSE.**

| | age | sex | height | BMI | smoker |
|---|---|---|---|---|---|
| $SPINC_1$-residual | -0.00 | -0.00 | 0.00 | 0.03 | 0.01 |
| $SPINC_2$-residual | -0.17 | -0.01 | 0.01 | -0.02 | -0.05 |
| $SPINC_3$-residual | -0.08 | -0.06 | -0.02 | -0.05 | -0.02 |
| $SPINC_4$-residual | 0.11 | -0.01 | -0.01 | -0.02 | 0.03 |
| $SPINC_5$-residual | 0.13 | 0.04 | 0.01 | 0.04 | 0.04 |
| $RSPINC_1$-residual | 0.18 | -0.02 | -0.02 | -0.01 | 0.04 |
| $RSPINC_2$-residual | 0.06 | 0.06 | 0.04 | 0.05 | 0.03 |

Supplementary Table 4: **Pearson correlation of (R)SPINCs residuals with lung-function-related covariates.** After residualizing the EDFs from the (R)SPINCS, they retain some information about the covariates.

| Fields group | $P$-value |
|---|---|
| Asthma (e.g. medical conditions self-report) | 1.18E-133 |
| Quality of spirograms collected (e.g. the number of spirometry measurements made) | 5.75E-62 |
| Breathing issues (e.g. wheeze or whistling in the chest in last year) | 3.83E-54 |
| Cognitive function (e.g. reaction time) | 1.43E-22 |
| Hayfever, allergic rhinitis or eczema (e.g. medical conditions self-report) | 6.99E-20 |
| Location (e.g. assessment center in Leeds) | 1.22E-17 |

Supplementary Table 5: **Selected groups of fields significantly correlated with (R)SPINCs after residualizing EDFs and covariates.** We note that the high correlation with "location" could be due to technical issues in particular assessment centers or ascertainment bias.

See the attached Excel table.

Supplementary Table 6: **Pearson correlation of (R)SPINCs residuals with UKB tabular fields.** After residualizing the EDFs and covariates from the (R)SPINCS, they still retain information about respiratory diseases such as asthma and allergic rhinitis, breathing issues, quality of spirograms, and cognitive function.

| Fields group | $P$-value |
|---|---|
| Pulse rate | $<$5.00e-300 |
| Pulse rate, automated reading | 5.65E-284 |
| Systolic blood pressure, automated reading | 1.17E-153 |
| Pulse wave reflection index | 1.12E-112 |
| Pulse rate, automated reading | 2.03E-77 |
| Diastolic blood pressure, automated reading | 6.74E-67 |
| Position of the shoulder on the pulse waveform | 3.98E-50 |
| Position of the pulse wave peak | 4.96E-42 |
| ECG, heart rate | 1.98E-35 |

Supplementary Table 7: **Selected groups of fields significantly correlated with PLENCs after residualizing EDFs and covariates.**

See the attached Excel table.

Supplementary Table 8: **Pearson correlation of PLENCs residuals with UKB tabular fields.** After residualizing the EDFs and covariates from the PLENCs, they still retain information about cardiovascular system such as pulse rate, systolic and diastolic blood pressure, ECG heart rate.

| Risk | Hazard Ratio | Lower 95% CI | Upper 95% CI | P |
|---|---|---|---|---|
| $SPINC_1$ | 1.08565 | 1.0516 | 1.1208 | 4.16E-07 |
| $SPINC_2$ | 1.13457 | 1.1010 | 1.1692 | 1.74E-16 |
| $SPINC_3$ | 0.67943 | 0.6534 | 0.7065 | 1.57E-83 |
| $SPINC_4$ | 1.07763 | 1.0449 | 1.1114 | 2.00E-06 |
| $SPINC_5$ | 1.00962 | 0.9794 | 1.0408 | 5.37E-01 |
| $RSPINC_1$ | 0.98222 | 0.9527 | 1.0127 | 2.49E-01 |
| $RSPINC_2$ | 1.06364 | 1.0316 | 1.0967 | 7.81E-05 |
| $FEV_1$ | 0.63971 | 0.6148 | 0.6656 | 8.10E-108 |
| FVC | 0.68493 | 0.6558 | 0.7153 | 2.15E-65 |
| PEF | 0.75200 | 0.7275 | 0.7773 | 7.72E-64 |
| $FEV_1$/FVC | 0.80562 | 0.7859 | 0.8258 | 1.07E-65 |
| $FEF_{25-75\%}$ | 0.68917 | 0.6630 | 0.7164 | 2.23E-79 |
| $PLENC_1$ | 0.87865 | 0.8339 | 0.9258 | 1.24E-06 |
| $PLENC_2$ | 0.76070 | 0.7208 | 0.8029 | 2.83E-23 |
| $PLENC_3$ | 1.07235 | 1.0177 | 1.1299 | 8.82E-03 |
| $PLENC_4$ | 1.01382 | 0.9615 | 1.0690 | 0.611623 |
| $PLENC_5$ | 0.99256 | 0.9419 | 1.0459 | 0.779784 |
| Absence of Notch | 1.07081 | 1.0245 | 1.1192 | 2.43E-03 |
| Position of Notch | 1.12913 | 1.0766 | 1.1842 | 5.89E-07 |
| Position of Peak | 1.11444 | 1.0536 | 1.1788 | 1.54E-04 |
| Position of Shoulder | 1.14546 | 1.0847 | 1.2096 | 1.04E-06 |
| Peak to Peak Time | 0.96973 | 0.9202 | 1.0219 | 2.50E-01 |

Supplementary Table 9: **Survival analysis hazard ratios per 1 standard deviation for SPINC, RSPINC, PLENC, and EDF risk scores.** Note that the $SPINC_1$ ($p = 0.0129$), $PLENC_1$ ($p = 0.0033$), $PLENC_2$ ($p = 0.0264$), and "Position of Notch" ($p = 0.0162$) models fail the proportional-hazards (PH) assumption and thus should be interpreted as hazard over time. The PH assumption holds for all other models.

| Phenotype | S-LDSC Intercept | S-LDSC Attenuation Ratio | S-LDSC SNP-heritability |
|---|---|---|---|
| $SPINC_1$ | 1.0224 (0.0181) | 0.0278 (0.0225) | 0.1302 (0.0071) |
| $SPINC_2$ | 1.0466 (0.0294) | 0.0294 (0.0186) | 0.2481 (0.0132) |
| $SPINC_3$ | 1.0431 (0.0161) | 0.0401 (0.0150) | 0.1604 (0.0066) |
| $SPINC_4$ | 1.0289 (0.0147) | 0.0591 (0.0300) | 0.0746 (0.0055) |
| $SPINC_5$ | 1.0019 (0.0103) | 0.0073 (0.0390) | 0.0428 (0.0029) |
| $RSPINC_1$ | 1.0231 (0.0237) | 0.0219 (0.0225) | 0.1615 (0.0113) |
| $RSPINC_2$ | 1.0113 (0.0114) | 0.0404 (0.0408) | 0.0446 (0.0034) |
| $PLENC_1$ | 1.0130 (0.0115) | 0.0360 (0.0318) | 0.1288 (0.0087) |
| $PLENC_2$ | 1.0177 (0.0109) | 0.0856 (0.0525) | 0.0682 (0.0072) |
| $PLENC_3$ | 1.0086 (0.0104) | 0.0526 (0.0637) | 0.0604 (0.0067) |
| $PLENC_4$ | 1.0000 (0.0105) | $< 0$ | 0.0366 (0.0067) |
| $PLENC_5$ | 1.0155 (0.0111) | 0.0745 (0.0534) | 0.0704 (0.0074) |

Supplementary Table 10: **S-LDSC results on SPINCs, RSPINCs, and PLENCs GWAS.** We computed the S-LDSC intercept, attenuation ratio and SNP-heritability. Values in parentheses are the standard error of the mean (s.e.m) obtained from S-LDSC.

Supplementary Table 11: **Potentially novel significant GWAS loci from SPINCs.** Using a stricter $P$-value of $1 \times 10^{-8}$, only displaying loci not found in Shrine et al 2023, GWAS catalog lung function search, or our own GWAS on EDFs. Conditional $P$-values ("cond. $P$") are obtained by conditional analysis on previously known variants. The closest genes are assigned to each variant. Variant IDs are in the form "chromosome:position_reference_alternate" using GRCh37 reference. In the "Source" column, "G" implies genotyped variants and "I" implies imputed variants.

| Chrom | Gene | $P$ | cond. $P$ | Variant ID | Range | Source |
|---|---|---|---|---|---|---|
| 1 | DNM3 | 2.30E-11 | 2.28E-10 | 1:172312769_G_A | 171809235-172463995 | G |
| 2 | ATP6V1E2 | 3.00E-09 | 3.60E-09 | 2:46737464_G_C | 46631429-47022818 | I |
| 2 | COL4A3 | 6.90E-09 | 9.39E-09 | 2:228161170_T_C | 228118156-228345413 | I |
| 3 | PLXND1 | 2.50E-11 | 2.25E-10 | 3:129274936_A_G | 129056595-129956119 | I |
| 3 | RBMS3 | 1.00E-08 | 2.23E-08 | 3:30304919_GT_G | 30270159-30484211 | I |
| 4 | AC097375.2 | 2.60E-09 | 9.07E-08 | 4:152949327_T_TA | 152782878-153100189 | I |
| 4 | BMP3 | 1.30E-16 | 3.44E-16 | 4:81952637_T_A | 81208992-82124698 | I |
| 4 | UNC5C | 2.00E-09 | 3.97E-09 | 4:96573857_C_T | 96451871-96666203 | I |
| 5 | AC079465.1 | 1.80E-11 | 9.27E-11 | 5:112739130_G_A | 112532935-113118026 | G |
| 5 | MCC | 1.80E-11 | 9.27E-11 | 5:112739130_G_A | 112532935-113118026 | G |
| 5 | CAMK2A | 2.20E-14 | 3.54E-13 | 5:149625611_A_T | 149565508-149665683 | I |
| 7 | FOXK1 | 4.00E-09 | 1.16E-08 | 7:4692566_A_G | 4676340-4746488 | I |
| 8 | HNF4G | 3.00E-09 | 2.50E-09 | 8:76362337_G_C | 76295984-77021947 | I |
| 8 | NRG1 | 9.80E-09 | 1.28E-07 | 8:32963969_G_C | 32642704-33263514 | I |
| 10 | AKR1C1 | 8.80E-09 | 2.21E-08 | 10:4961278_T_C | 4829609-5108821 | I |
| 10 | BICC1 | 1.20E-09 | 4.13E-10 | 10:60343348_G_C | 60169970-60396362 | I |
| 10 | VTI1A | 4.10E-09 | 7.83E-09 | 10:114606290_C_T | 114523887-114731845 | I |
| 11 | CHRDL2 | 4.60E-22 | 2.68E-22 | 11:74427921_C_T | 74376844-74838572 | I |
| 11 | CYB561A3 | 2.10E-14 | 1.03E-14 | 11:61126858_C_T | 60838260-61282934 | G |
| 11 | MRPL23 | 7.40E-09 | 4.22E-08 | 11:2019174_C_T | 1874072-2041831 | I |
| 15 | EMC7 | 6.00E-09 | 3.60E-09 | 15:34379605_C_T | 34166481-34424891 | G |
| 15 | SEMA6D | 1.50E-10 | 6.63E-11 | 15:47741212_T_G | 47649593-47991515 | G |
| 17 | KCNJ16 | 1.10E-17 | 3.24E-17 | 17:67962340_C_G | 67544154-68024377 | I |
| 19 | FCHO1 | 8.00E-09 | 2.56E-08 | 19:17862267_TC_T | 17818037-17895874 | I |
| 22 | TRIOBP | 7.70E-13 | 2.47E-11 | 22:38176979_T_G | 37977713-38449820 | I |

Supplementary Table 12: **Potentially novel significant GWAS loci from RSPINCs.** Using a stricter $P$-value of $1 \times 10^{-8}$, only displaying loci not found in Shrine et al 2023, GWAS catalog lung function search, or our own GWAS on EDFs. Conditional $P$-values ("cond. $P$") are obtained by conditional analysis on previously known variants. The closest genes are assigned to each variant. Variant IDs are in the form "chromosome:position_reference_alternate" using GRCh37 reference. In the "Source" column, "G" implies genotyped variants and "I" implies imputed variants.

| Chrom | Gene | $P$ | cond. $P$ | Variant ID | Range | Source |
|---|---|---|---|---|---|---|
| 1 | HHIPL2 | 1.40E-10 | 1.05E-09 | 1:222548602_T_C | 222236497-222560502 | I |
| 2 | LPIN1 | 2.20E-09 | 8.13E-09 | 2:12065180_A_G | 12044820-12145188 | I |
| 2 | PRKCE | 1.10E-09 | 7.43E-10 | 2:46218502_A_ATT | 46165972-46285524 | I |
| 2 | TMEM247 | 7.70E-10 | 4.06E-10 | 2:46692974_C_CT | 46583593-46870757 | I |
| 3 | H1-8 | 9.00E-11 | 6.84E-10 | 3:129263140_A_G | 129056595-129956119 | G |
| 4 | BMP3 | 2.20E-28 | 7.16E-28 | 4:81952637_T_A | 81208992-82124698 | I |
| 4 | OCIAD1 | 1.70E-09 | 5.92E-08 | 4:48810179_G_A | 48342682-53065669 | I |
| 5 | AC010451.3 | 1.50E-09 | 5.27E-08 | 5:4962498_C_T | 4940255-5067870 | I |
| 5 | AC027343.2 | 1.00E-08 | 5.23E-08 | 5:7158442_G_A | 7143293-7371420 | I |
| 5 | MIR4458HG | 2.70E-09 | 1.98E-09 | 5:8531288_C_G | 8495149-8584403 | I |
| 6 | ALDH8A1 | 7.50E-10 | 9.39E-10 | 6:135117710_TA_T | 135022253-135165945 | I |
| 6 | TBX18 | 3.00E-12 | 1.83E-12 | 6:85211448_T_C | 85134017-85581296 | I |
| 7 | AC019117.4 | 1.70E-09 | 2.02E-08 | 7:17441082_C_G | 17169922-17569101 | I |
| 7 | FERD3L | 1.00E-11 | 1.58E-11 | 7:19446881_GT_G | 19223257-19630474 | I |
| 8 | LINC02855 | 1.30E-12 | 5.45E-14 | 8:122668595_T_G | 122625186-122792872 | I |
| 8 | MRPS28 | 8.10E-09 | 3.19E-08 | 8:80756803_A_G | 80605017-81070612 | I |
| 8 | ZNF703 | 9.70E-09 | 2.63E-08 | 8:37532984_A_G | 37408632-37658001 | G |
| 10 | AKR1C1 | 6.70E-10 | 1.14E-08 | 10:4985193_T_C | 4829609-5108821 | I |
| 10 | SLC16A9 | 2.30E-09 | 5.46E-10 | 10:61320597_G_A | 61320597-61380392 | I |
| 11 | GRM5 | 1.00E-11 | 3.33E-12 | 11:88486055_A_G | 88329190-88952464 | I |
| 11 | NAV2 | 1.40E-18 | 2.37E-17 | 11:19973306_C_G | 19965487-20019667 | I |
| 11 | QSER1 | 6.00E-13 | 8.42E-13 | 11:32956492_C_T | 32385925-33241651 | G |
| 11 | XRRA1 | 2.50E-28 | 4.13E-27 | 11:74628743_C_T | 74413843-74838572 | I |
| 11 | YAP1 | 1.70E-09 | 2.24E-09 | 11:102002913_C_T | 101761385-102157900 | I |
| 13 | LINC01069 | 5.50E-09 | 1.06E-08 | 13:78651299_G_A | 78125293-78807836 | I |
| 14 | FLRT2 | 6.60E-10 | 1.23E-08 | 14:86646016_T_G | 86643782-86646282 | I |
| 15 | MCTP2 | 7.80E-10 | 4.86E-09 | 15:94357066_C_T | 94273406-94508663 | I |
| 15 | SEMA6D | 1.70E-17 | 2.27E-17 | 15:47734845_A_G | 47649593-47991515 | I |
| 20 | PTPN1 | 5.00E-09 | 4.24E-10 | 20:49096493_A_T | 48986299-49238073 | I |
| 22 | TRIOBP | 4.10E-18 | 1.99E-16 | 22:38176979_T_G | 37977713-39285885 | I |

| Chrom | Gene | $P$ | Variant ID | Range | Source |
|---|---|---|---|---|---|
| 1 | AKR1A1 | 1.50E-12 | 1:46026397_CT_C | 44973546-46891925 | I |
| 1 | NOS1AP | 6.20E-09 | 1:162161339_C_G | 162014632-162265976 | I |
| 2 | EFEMP1 | 4.30E-09 | 2:56095994_C_G | 55666224-56257941 | I |
| 3 | GLYCTK | 1.10E-11 | 3:52333671_C_G | 52214640-53553745 | I |
| 3 | ITGA9 | 1.20E-09 | 3:37596805_C_G | 37520793-37663628 | I |
| 3 | LINC02029 | 1.10E-10 | 3:156795414_G_T | 156791268-156848024 | I |
| 3 | RSRC1 | 2.50E-10 | 3:158173507_G_A | 157582078-158540961 | I |
| 4 | PPARGC1A | 1.40E-09 | 4:23951018_C_G | 23926728-24114572 | I |
| 7 | MKLN1 | 3.60E-09 | 7:130973495_C_T | 130946690-131213850 | I |
| 10 | NOC3L | 6.20E-09 | 10:96122543_A_G | 95971321-96993955 | G |
| 11 | FADS1 | 2.60E-17 | 11:61569830_C_T | 61523300-61678754 | G |
| 11 | FADS2 | 2.60E-17 | 11:61569830_C_T | 61523300-61678754 | G |
| 13 | FGF9 | 3.60E-11 | 13:22861921_A_G | 22853646-22911560 | I |
| 16 | CDH13 | 3.00E-09 | 16:82750051_A_G | 82679422-82924972 | I |
| 16 | CNOT1 | 4.60E-09 | 16:58566304_G_A | 58525312-58866367 | G |
| 16 | HNRNPA1P48 | 8.40E-09 | 16:51578359_C_G | 51451739-52103288 | I |
| 16 | TEKT5 | 1.60E-10 | 16:10740982_G_C | 10695121-10810459 | I |
| 18 | FHOD3 | 1.60E-13 | 18:34289285_G_T | 34185526-34942005 | I |
| 19 | CILP2 | 1.90E-10 | 19:19649748_G_C | 19087498-19865077 | I |
| 20 | GNAS | 3.40E-09 | 20:57466093_G_T | 57221133-57544177 | I |

Supplementary Table 13: **Potentially novel significant GWAS loci from PLENCs.** All loci not found in GWAS Catalog cardiovascular disease search. The closest genes are assigned to each variant. Variant IDs are in the form "chromosome:position_reference_alternate" using GRCh37 reference. In the "Source" column, "G" implies genotyped variants and "I" implies imputed variants.

| Method | AUC-ROC | AUC-PR | Top decile prevalence | Pearson R |
|---|---|---|---|---|
| Ratio (1) | 0.534 | 0.152 | 0.161 | 0.039 |
| EDFs (5) | 0.539 | 0.157 | 0.172 | 0.048 |
| Raw PC (5) | 0.524 | 0.149 | 0.153 | 0.030 |
| EDFs + RSPINCs (7) | 0.548* | 0.161* | 0.182* | 0.060* |
| SPINCs (5) | **0.553**** | **0.163**** | **0.182**** | **0.065**** |

Supplementary Table 14: **Asthma PRS performance in UK Biobank**. "Ratio" = $FEV_1/FVC$, "Manual" = {FVC, $FEV_1$, PEF, $FEF_{25\text{-}75\%}$, $FEV_1/FVC$}. * statistically significant improvement over "EDFs (5)" with paired bootstrapping with 95% confidence.

| Method | AUC-ROC | AUC-PR | Top decile prevalence | Pearson R |
|---|---|---|---|---|
| Ratio (1) | 0.543 | 0.073 | 0.080 | 0.037 |
| EDFs (5) | 0.547 | 0.075 | 0.083 | 0.041 |
| Raw PC (5) | 0.525 | 0.069 | 0.074 | 0.022 |
| EDFs + RSPINCs (7) | **0.550*** | **0.076*** | 0.084 | **0.044*** |
| SPINCs (5) | 0.549 | **0.076** | **0.086** | **0.044** |

Supplementary Table 15: **COPD PRS performance in UK Biobank**. "Ratio" = $FEV_1/FVC$, "EDFs" = {FVC, $FEV_1$, PEF, $FEF_{25\text{-}75\%}$, $FEV_1/FVC$}. * indicates statistically significant improvement over "EDFs (5)" with paired bootstrapping with 95% confidence.

| PRS pheno | Asthma PRS weight | COPD PRS weight |
|---|---|---|
| $SPINC_1$ | 0.0217 | 0.0057 |
| $SPINC_2$ | 0.0581 | 0.0213 |
| $SPINC_3$ | -0.0402 | -0.0285 |
| $SPINC_4$ | -0.0655 | 0.0073 |
| $SPINC_5$ | -0.1650 | -0.0188 |
| $FEV_1$ | 0.0049 | 0.0188 |
| FVC | -0.0227 | -0.0374 |
| PEF | 0.0022 | 0.0001 |
| $FEV_1/FVC$ | -0.4720 | -0.3541 |
| $FEF_{25\text{-}75\%}$ | -0.0471 | -0.0090 |

Supplementary Table 16: **SPINCs and EDFs PRS weights for pulmonary traits.**

| | Method | AUC-ROC | AUC-PR | Top decile prevalence | Pearson R |
|---|---|---|---|---|---|
| Non-Hispanic White | EDFs | 0.586 | 0.604 | 0.650 | 0.154 |
| | EDFs + RSPINCs | 0.589* | 0.605 | 0.656 | 0.158* |
| | SPINCs | **0.622*** | **0.635*** | **0.715*** | **0.212*** |
| African American | EDFs | 0.538 | 0.358 | 0.360 | 0.064 |
| | EDFs + RSPINCs | 0.536 | 0.356 | 0.358 | 0.062 |
| | SPINCs | **0.559*** | **0.372** | **0.374** | **0.096*** |

Supplementary Table 17: **COPDGene COPD PRS performance**. Bold numbers are the highest in the same category. * indicates statistically significant improvement over EDFs with paired bootstrapping $p < 0.05$. EDFs: $FEV_1$, FVC, $FEV_1/FVC$, PEF, and $FEF_{25\text{-}75\%}$.

| PRS pheno | HTN PRS weight | SBP PRS weight |
|---|---|---|
| $PLENC_1$ | 0.1326 | 15.2648 |
| $PLENC_2$ | -0.0059 | -0.5420 |
| $PLENC_3$ | 0.0895 | 2.8647 |
| $PLENC_4$ | 0.1949 | 21.6961 |
| $PLENC_5$ | -0.0411 | -14.0404 |
| Absence of notch | 0.3176 | 18.6592 |
| Position of notch | 0.0004 | -0.1606 |
| Position of peak | 0.0370 | 4.7233 |
| Position of shoulder | -0.0197 | -3.4335 |
| Peak-to-peak time | -0.0012 | 0.0220 |

Supplementary Table 18: **PLENCs and EDFs PRS weights for cardiovascular traits.**

| SPINC | ID | Description | Beta | R | P-value | SE |
|---|---|---|---|---|---|---|
| $SPINC_3$ | `continuous-3063-both_sexes-irnt` | FEV1 | 3.014 | 0.452 | < 5.00e-300 | 0.009 |
| $SPINC_2$ | `phecode-593-both_sexes` | Hematuria | -0.736 | -0.444 | <5.00e-300 | 0.002 |
| $SPINC_2$ | `phecode-695.4-both_sexes` | Lupus | 0.714 | 0.431 | <5.00e-300 | 0.002 |
| $SPINC_2$ | `categorical-20004-both_sexes-1228` | Thyroid radioablation therapy | 0.716 | 0.432 | <5.00e-300 | 0.002 |
| $SPINC_3$ | `continuous-20150-both_sexes-irnt` | FEV1, best measure | 3.051 | 0.458 | <5.00e-300 | 0.009 |
| $SPINC_3$ | `continuous-20154-both_sexes-irnt` | FEV1 % predicted | 3.409 | 0.511 | <5.00e-300 | 0.009 |
| $SPINC_2$ | `categorical-20086-both_sexes-8` | Gluten-free diet | 0.819 | 0.494 | <5.00e-300 | 0.002 |
| $SPINC_2$ | `categorical-41245-both_sexes-1860` | Urology consultant | -0.686 | -0.414 | <5.00e-300 | 0.002 |
| $SPINC_2$ | `categorical-6144-both_sexes-3` | Never eat wheat | 0.879 | 0.530 | <5.00e-300 | 0.002 |
| $SPINC_2$ | `phecode-242.1-both_sexes` | Graves' disease | 0.672 | 0.405 | <5.00e-300 | 0.002 |
| $SPINC_2$ | `categorical-1448-both_sexes-4` | "Other" bread type | 0.826 | 0.498 | <5.00e-300 | 0.002 |
| $SPINC_2$ | `continuous-FEV1FVC-both_sexes-irnt` | FEV1/FVC ratio | -1.329 | -0.802 | <5.00e-300 | 0.002 |
| $SPINC_2$ | `categorical-20002-both_sexes-1371` | Sarcoidosis | 0.774 | 0.467 | <5.00e-300 | 0.002 |
| $SPINC_2$ | `continuous-3064-both_sexes-irnt` | PEF | -0.830 | -0.500 | <5.00e-300 | 0.002 |
| $SPINC_2$ | `icd10-E05-both_sexes` | Thyrotoxicosis | 0.676 | 0.408 | <5.00e-300 | 0.002 |

Supplementary Table 19: **Top associations of SPINCs PRSs with UK Biobank phenotype PRSs.** The PRS of each SPINCs coordinate was compared to phenotype PRSs generated from GWAS summary statistics from the Pan-UKBB consortium. Results shown are limited to those with $|R| \geq 0.4$. Full results are available in Supplementary Table 20. R, Pearson R; SE, standard error; FEV1, forced expiratory volume in 1 second; FVC, forced vital capacity; PEF, peak expiratory flow.

See the attached Excel table.

Supplementary Table 20: **All associations of SPINCs PRSs with UK Biobank phenotype PRSs.** Same as Supplementary Table 19, full results.

| RSPINC | ID | Description | Beta | R | P-value | SE |
|---|---|---|---|---|---|---|
| $RSPINC_1$ | phecode-593-both_sexes | Hematuria | 1.007 | 0.439 | <5.00e-300 | 0.003 |
| $RSPINC_1$ | phecode-695.4-both_sexes | Lupus | -1.019 | -0.444 | <5.00e-300 | 0.003 |
| $RSPINC_1$ | phecode-695.42-both_sexes | Systemic lupus erythematosus | -0.975 | -0.425 | <5.00e-300 | 0.003 |
| $RSPINC_1$ | icd10-M32-both_sexes | Systemic lupus erythematosus | -0.975 | -0.425 | <5.00e-300 | 0.003 |
| $RSPINC_1$ | icd10-E05-both_sexes | Thyrotoxicosis | -0.962 | -0.419 | <5.00e-300 | 0.003 |
| $RSPINC_1$ | phecode-242-both_sexes | Thyrotoxicosis | -0.927 | -0.404 | <5.00e-300 | 0.003 |
| $RSPINC_1$ | categorical-41245-both_sexes-1860 | Urology consultant | 0.987 | 0.430 | <5.00e-300 | 0.003 |
| $RSPINC_1$ | categorical-6144-both_sexes-3 | Never eat wheat | -1.154 | -0.503 | <5.00e-300 | 0.003 |
| $RSPINC_2$ | categorical-20533-both_sexes-20533 | Trouble falling asleep | 25.894 | 0.560 | <5.00e-300 | 0.058 |
| $RSPINC_1$ | categorical-20004-both_sexes-1228 | Thyroid radioablation therapy | -0.955 | -0.416 | <5.00e-300 | 0.003 |
| $RSPINC_1$ | categorical-41200-both_sexes-M459 | Unspecified diagnostic endoscopic bladder exam | 0.946 | 0.412 | <5.00e-300 | 0.003 |
| $RSPINC_1$ | categorical-41200-both_sexes-W365 | Diagnostic extraction of bone marrow NEC | -0.980 | -0.427 | <5.00e-300 | 0.003 |
| $RSPINC_1$ | categorical-20086-both_sexes-8 | Gluten-free diet | -1.089 | -0.475 | <5.00e-300 | 0.003 |
| $RSPINC_1$ | categorical-1448-both_sexes-4 | "Other" bread type | -1.116 | -0.486 | <5.00e-300 | 0.003 |
| $RSPINC_1$ | categorical-20002-both_sexes-1371 | Sarcoidosis | -1.054 | -0.459 | <5.00e-300 | 0.003 |
| $RSPINC_1$ | continuous-3064-both_sexes-irnt | PEF | 1.166 | 0.508 | <5.00e-300 | 0.003 |
| $RSPINC_1$ | continuous-FEV1FVC-both_sexes-irnt | FEV1/FVC ratio | 1.218 | 0.530 | <5.00e-300 | 0.003 |

Supplementary Table 21: **Top associations of RSPINCs PRSs with UK Biobank phenotype PRSs.** The PRS of each RSPINCs coordinate was compared to phenotype PRSs generated from GWAS summary statistics from the Pan-UKBB consortium. Results shown are limited to those with $|R| \geq 0.4$. Full results are available in Supplementary Table 20. R, Pearson R; SE, standard error; NEC, necrosis; PEF, peak expiratory flow; FEV1, forced expiratory volume in 1 second; FVC, forced vital capacity.

See the attached Excel table.

Supplementary Table 22: **All associations of RSPINCs PRSs with UK Biobank phenotype PRSs.** Same as Supplementary Table 21, full results.

| PLENCs | ID | Description | Beta | R | P-value | SE |
|---|---|---|---|---|---|---|
| $RSPINC_1$ | continuous-12336-both_sexes-irnt | Ventricular rate | -2.909 | -0.438 | <5.00e-300 | 0.009 |
| $RSPINC_1$ | continuous-95-both_sexes-irnt | Pulse rate (during blood-pressure measurement) | -3.348 | -0.504 | <5.00e-300 | 0.009 |
| $RSPINC_1$ | continuous-4194-both_sexes-irnt | Pulse rate | -4.504 | -0.678 | <5.00e-300 | 0.007 |
| $RSPINC_1$ | continuous-102-both_sexes-irnt | Pulse rate, automated reading | -3.667 | -0.552 | <5.00e-300 | 0.008 |
| $RSPINC_1$ | continuous-5983-both_sexes-irnt | ECG, heart rate | -3.486 | -0.525 | <5.00e-300 | 0.009 |

Supplementary Table 23: **Top associations of PLENCs PRSs with UK Biobank phenotype PRSs.** The PRS of each PLENCs coordinate was compared to phenotype PRSs generated from GWAS summary statistics from the Pan-UKBB consortium. Results shown are limited to those with $|R| \geq 0.4$. Full results are available in Supplementary Table 24. R, Pearson R; SE, standard error.

See the attached Excel table.

Supplementary Table 24: **All associations of PLENCs PRSs with UK Biobank phenotype PRSs.** Same as Supplementary Table 23, full results.

| Trait 1 | Trait 2 | $\widehat{\text{GCP}}$ (SE) | $\log_{10} p_{\text{LCV}}$ | $\widehat{\rho_g}$ (SE) |
|---|---|---|---|---|
| $\text{SPINC}_5$ | Asthma | -0.71(0.12) | -42.1 | -0.20(0.06) |
| $\text{SPINC}_3$ | Sarcoidosis | -0.83(0.12) | -21.4 | -0.24(0.10) |
| $\text{SPINC}_3$ | Thyrotoxicosis | -0.78(0.15) | -13.9 | -0.24(0.10) |
| FVC | Sarcoidosis | -0.77(0.15) | -13.8 | -0.22(0.09) |
| $\text{FEV}_1$ | Sarcoidosis | -0.79(0.14) | -13.2 | -0.28(0.14) |
| $\text{RSPINC}_1$ | Lupus | -0.83(0.12) | -12.8 | -0.33(0.26) |
| FVC | Thyrotoxicosis | -0.75(0.17) | -12.6 | -0.20(0.11) |
| $\text{FEF}_{25\text{-}75\%}$ | Lupus | -0.84(0.12) | -12.4 | -0.33(0.19) |
| $\text{FEV}_1$/FVC | Lupus | -0.79(0.15) | -9.8 | -0.24(0.17) |
| $\text{FEV}_1$ | Thyrotoxicosis | -0.79(0.15) | -7.6 | -0.26(0.15) |
| $\text{FEV}_1$ | Gluten-free-diet | -0.72(0.20) | -7.4 | -0.28(0.17) |
| $\text{SPINC}_2$ | Lupus | -0.75(0.17) | -7.2 | 0.33(0.23) |
| $\text{FEV}_1$ | Asthma | -0.43(0.10) | -7.2 | -0.33(0.05) |
| $\text{FEV}_1$/FVC | COPD | 0.84(0.13) | -6.7 | -0.58(0.09) |
| $\text{SPINC}_2$ | COPD | 0.82(0.14) | -6.7 | 0.52(0.08) |
| $\text{FEF}_{25\text{-}75\%}$ | COPD | 0.82(0.14) | -5.8 | -0.55(0.09) |
| $\text{SPINC}_3$ | Gluten-free-diet | -0.68(0.22) | -4.7 | -0.18(0.14) |
| $\text{PLENC}_1$ | Hypertension | -0.35(0.10) | -4.3 | 0.23(0.07) |
| $\text{FEF}_{25\text{-}75\%}$ | Sarcoidosis | -0.53(0.19) | -4.1 | -0.25(0.19) |
| $\text{SPINC}_3$ | Asthma | -0.29(0.09) | -3.7 | -0.22(0.05) |

Supplementary Table 25: **Significant trait pairs based on LCV.** $\widehat{\text{GCP}}$: the estimated genetic causality proportion. SE: the standard error. $p_{\text{LCV}}$: the $p$-value from the latent causal variable model testing the null hypothesis that GCP = 0. $\widehat{\rho_g}$: the estimated genetic correlation.

See the attached Excel table.

Supplementary Table 26: **Latent causal variable (LCV) analysis of five SPINCs, two RSPINCs, and five EDFs on asthma, COPD, Lupus, Thyrotoxicosis, Gluten-free-diet, and Sarcoidosis.** $\widehat{\text{GCP}}$: the estimated genetic causality proportion. SE: the standard error. $p_{\text{LCV}}$: the $p$-value from the latent causal variable model testing the null hypothesis that GCP = 0. $\widehat{\rho_g}$: the estimated genetic correlation.

| Term | Description | Region P | Gene P | Num regions |
|---|---|---|---|---|
| GO:0048598 | embryonic morphogenesis | 2.05e-08 | 1.02e-18 | 106 |
| GO:0001501 | skeletal system development | 6.91e-08 | 2.66e-15 | 92 |
| GO:0002009 | morphogenesis of an epithelium | 5.16e-06 | 1.78e-13 | 82 |
| GO:0048568 | embryonic organ development | 5.20e-06 | 3.22e-13 | 81 |
| GO:0060562 | epithelial tube morphogenesis | 2.01e-05 | 8.68e-13 | 67 |
| GO:0035239 | tube morphogenesis | 2.21e-05 | 2.67e-14 | 73 |
| GO:0061138 | morphogenesis of a branching epithelium | 4.12e-05 | 2.96e-12 | 48 |
| GO:0001655 | urogenital system development | 7.85e-05 | 7.01e-14 | 71 |

Supplementary Table 27: **Strongest term enrichments of the RSPINCs plus EDFs loci.** Enrichments were computed using GREAT with default parameters. The 122 total terms significant at Bonferroni-corrected $P \leq 10^{-4}$ by both the region-based binomial and gene-based hypergeometric tests were filtered to those with region fold enrichment $\geq 2$.

| Method (# traits) | Sample size | Total | Known | Novel |
|---|---|---|---|---|
| Shrine 2023 + GWAS Catalog | $> 581\text{K}^*$ | 1104 | – | – |
| Shrine 2023 | $581\text{K}$ | 754 | – | – |
| EDFs (5) | 325K | 628 | 596 | 32 |
| PCA (5) | 325K | 485 | 464 | 21 |
| **SPINCs** (5) | 325K | 584 | 517 | 67 |
| **EDFs + RSPINCs** (7) | 325K | 671 | 609 | 62 |

Supplementary Table 28: **Comparison of (R)SPINCs loci with previous GWAS using inverse-normal transformation.** Expert-defined features (EDFs) are $FEV_1$, FVC, $FEV_1$/FVC, PEF, and $FEF_{25\text{-}75\%}$. "Known" and "novel" is in reference to lung function loci in Shrine et al. Nat. Genet. 2023 and GWAS catalog. Inverse-normal transformation is performed on all phenotypes.
* GWAS in Shrine et al. Nat. Genet. 2023 has 580,869 individuals and other previous GWAS in the GWAS catalog may have more individuals.
